# The decrease in childhood vaccination coverage in the Netherlands from birth cohort 2008 to 2020 and its sociodemographic determinants

**DOI:** 10.1101/2025.04.24.25326341

**Authors:** Joyce Pijpers, Annika van Roon, Maarten Schipper, Marijn Stok, Susan van den Hof, Ruben van Gaalen, Susan Hahné, Hester de Melker

## Abstract

**Introduction:** To understand differences in vaccination coverage between population subgroups in the Netherlands over time, we studied sociodemographic factors associated with measles-mumps-rubella (MMR) and diphtheria-tetanus-pertussis-poliomyelitis (DTaP-IPV) vaccination.

**Methods:** We conducted a national retrospective database study including children born in 2008-2020. Individual-level data-linkage allowed examination of associations of sociodemographic variables with MMR and DTaP-IPV vaccination status at 2 years of age. Coverage for each variable, stratified by birth cohort, was calculated. Multivariable Poisson regression assessed independent associations and changes in coverage over time.

**Results:** MMR coverage decreased in all population subgroups (overall 95% to 89% in 2008 and 2020 cohorts), with more substantial declines in some groups. The multivariable analysis showed that Dutch children of non-Dutch origin, particularly those of Moroccan and Turkish origin showed more pronounced declines (−25% and -12% compared to children of Dutch origin in cohort 2020, respectively). Among children not attending day care and children living in larger families (≥4 children), a faster decline in coverage was observed compared to those attending day care and living in smaller families (both -12% in cohort 2020). Among children of self-employed mothers and children in the lowest income households, lower coverage was observed compared to children of mothers in employment and children in the highest income households (−8% and -7% in cohort 2020, respectively). Nearly identical trends were observed for DTaP-IPV vaccination.

**Conclusion:** Our study reveals a significant decline in childhood vaccination coverage in the Netherlands, with increasing disparities between sociodemographic groups. This is crucial for prioritising vaccination efforts to protect public health equitably.

## Introduction

Childhood vaccination programmes are the single most effective intervention to protect children’s health. The Expanded Programme on Immunisation is estimated to have averted 154 million deaths globally, the majority in children below five years of age, between 1974 and 2024 [1]. Since 1957, the Netherlands’ National Immunisation Program (NIP) has provided free, voluntary vaccinations against 13 severe infectious diseases and has significantly reduced child and young adult mortality [2]. Despite its success and historically high coverage, childhood vaccination coverage has declined in recent years. Coverage for diphtheria, tetanus, pertussis, poliomyelitis vaccination (DTaP-IPV) and measles, mumps and rubella vaccination (MMR) in newborns dropped from 95% in the 2008-2010 birth cohort to 93% in 2015-2016 cohorts and further to 88% and 89%, respectively, in the 2020 cohort[3]. Historically, NIP vaccine coverage has been relatively low in regions where socio-demographically clustered orthodox reformed individuals live (Bible Belt region) [4, 5]. However, in recent years a decline was observed in coverage of all childhood vaccinations outside of the Bible Belt region, specifically in large cities [3].

(Inter)national research has summarized sociodemographic factors associated with childhood vaccination coverage. Lower (maternal) education level and lower socioeconomic status were associated with lower routine childhood vaccination coverage [6, 7]. Similarly, lower parental income, lower parental education level and not being the first-born child were independently associated with lower MMR vaccination uptake [8]. A Dutch study among infants born in 2005 found that parental country of birth and religious objections to vaccination were significant determinants of vaccination uptake [9]. Children with at least one parent born abroad, particularly those with both parents born in Turkey or Morocco, had 20-30% lower odds of being fully vaccinated. [9]. This lower uptake aligns with more recent Dutch studies on HPV and meningococcal (ACWY) vaccination [10-12]. In these studies, data on country of origin were available at individual level, while socioeconomic status (SES) variables and proxy variables for religion (voting proportions for the Reformed Political Party) were available at an aggregated level.

The aim of this study is to identify socio-demographic factors associated with changes in vaccination coverage over time (aligning with the WHO Tailoring Immunization Programmes (TIP) approach’s ‘situational analysis’) [13-15], employing novel, detailed individual-level data. In turn, social scientific research can inform targeted interventions to improve MMR and DTaP-IPV vaccination coverage, reduce disparities, and promote equitable access to vaccination and information [13-15].

## Methods

### Study design and population

For this retrospective database study, multiple data sources were linked using a unique identifier to assess determinants of MMR and DTaP-IPV vaccination coverage in the Netherlands of children born in 2008-2020. In this period sociodemographic registries maintained consistent variables and data collection methods. Data linkage was done in the secured remote access environment of Statistics Netherlands (CBS).

Children with birth year 2008–2020 were selected from the personal records database (BRP). Vaccination status for both MMR and DTaP-IPV was determined at 24 months of age. For this purpose, we created separate databases for each birth cohort. National registries regarding death and migration were used to determine whether children had been eligible for MMR1 vaccination at 14 months of age and DTaP-IPV booster vaccination at 11 months. Children were excluded in case they deceased or moved abroad ≤11 and ≤14 months of age or had moved to the Netherlands >11 and >14 months of age, respectively. Lastly, we linked children to their parents based on a CBS register containing child-parent relations. Children who could not be linked to a parent were excluded.

### Outcome measure

The binary outcome measures in this study were having received 1-dose MMR vaccination before the second birthday and having received at least three DTaP-IPV vaccinations (scheduled at 2 and/or 3 months of age, at 5 months and 11 months) before the second birthday (corresponding with full vaccination uptake). Age at time of vaccination was calculated based on birth date (month-year) and date of vaccine administration. Vaccination data was available from Praeventis, the register of the Dutch NIP [16]. The vaccination dataset including MMR and DTaP-IPV vaccinations was linked to the study population based on the same unique identifier. Children that could not be linked to the vaccination database were included as unvaccinated.

### Covariates

The following individual-level variables provided by CBS [17] were included as potential determinants for MMR1 and DTaP-IPV booster vaccination: year of birth, mother’s education level (high vs not high (including intermediate, low and unknown (20%)), household disposable income (quartiles), mother’s income source (employed; self-employed; benefit recipient; pensioner; student), family size (1-3 children; ≥4 children; institutional household), urbanisation level (extremely; strongly; moderately; hardly; not), day care attendance (yes; no), migration status (Dutch origin: all (grand)parents born in the Netherlands; migrant or child of at least one first-generation migrant; child of second-generation migrants or of one second-generation migrant and one Dutch parent). Migrant children and children of first-generation migrants were grouped due to low numbers. Country of origin was categorized as the Netherlands; Europe (excluding the Netherlands); Morocco; Turkey; Suriname; Dutch Caribbean; Indonesia; Other, America/Oceania; Other, Africa; Other, Asia). Indonesia; Other (Africa, Asia, America/Oceania). Country of origin was primarily defined by the country of birth (CoB) of the child itself. If the child was born in the Netherlands, country of origin was first determined by the parents’ country of birth (CoB). If both parents were born in the Netherlands, the grandparents’ CoB was then considered. Within this definition, the mother’s CoB was prioritized over the father’s, and the grandmother’s CoB was prioritized over the grandfather’s. Foreign-born origins were given precedence over Dutch origin if present in the lineage.

### Statistical analysis

Datasets for the separate birth cohorts were merged into one dataset containing all children born in 2008–2020 who were eligible for MMR1 vaccination and one dataset for DTaP-IPV. Descriptive analysis summarized the sociodemographic characteristics of the vaccinated and unvaccinated children. Vaccination coverage was calculated as the number of children vaccinated before their second birthday divided by the total population eligible for each category of the sociodemographic variables.

We used a multivariable Poisson regression model to assess the independent association between vaccination and the sociodemographic variables. The results are reported as adjusted rate ratios (RRs) with 95% confidence interval levels and p-values. The level of significance was set at 5%. Since the proportion of missing categorical values was small (≤3%), they were excluded from the multivariable regression analysis. To account for potential non-linearity of the trend of vaccination coverage over time, the variable birth cohort was included in the model as a categorical variable. The variable migration status was excluded from the multivariable analysis, given its complete overlap with the variable country of origin. To assess changes in the associations over time, we included interaction terms between the variable birth cohort and each determinant in the multivariable model. We report the RRs for each category within the sociodemographic variables per birth cohort and subsequently, we calculated the adjusted relative change in vaccination coverage ((RR – 1)*100), based on the RR of the total effect (main effect birth year + main effect socio-demographic variable + interaction effect). We calculated the number of unvaccinated children in each category for the most recent birth cohort to pinpoint where interventions could have the greatest impact on vaccination coverage (Supplementary Table S6).

All analyses were performed in R version 4.4.0 [18].

## Results

### Descriptive analysis

The total population of the 13 included birth cohorts for MMR and DTaP-IPV included 2,323,838 and 2,331,199 children, respectively (Table 1). Overall, by the age of 2 years, 2,174,299 children (94%) were vaccinated for MMR, and 2,172,402 children (93%) were vaccinated for DTaP-IPV. The distribution of sociodemographic groups in both the MMR and DTaP-IPV population are described in Table 1. Of children eligible for both MMR and DTaP-IPV vaccination (n=2,319,001), 97.2% had the same vaccination status(either vaccinated or unvaccinated), 1.4% only received the MMR vaccination and 1.3% only received the DTaP-IVP vaccination. Supplementary Table S1 presents the distribution of the sociodemographic variables per birth cohort.

**Table 1.**
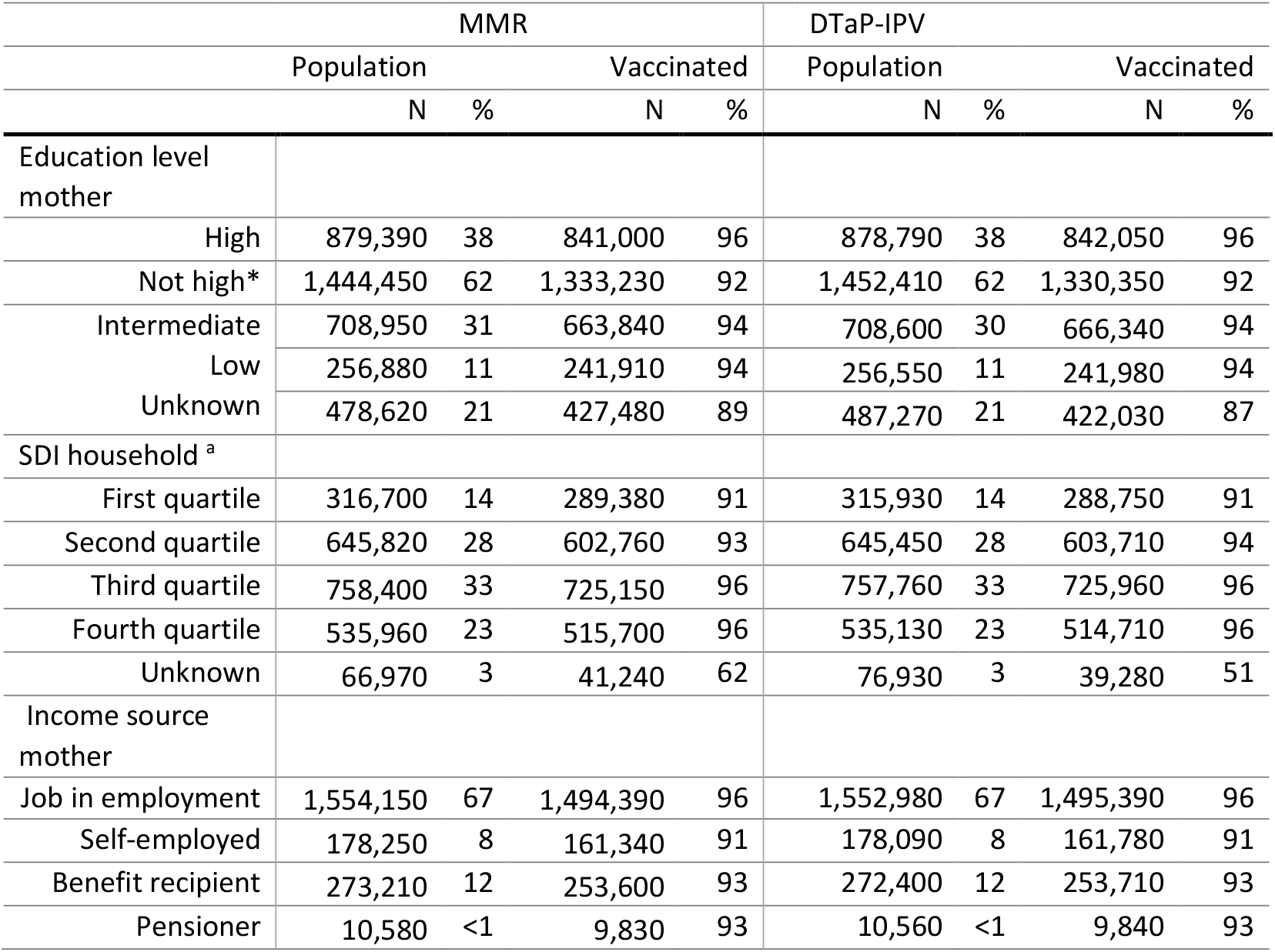
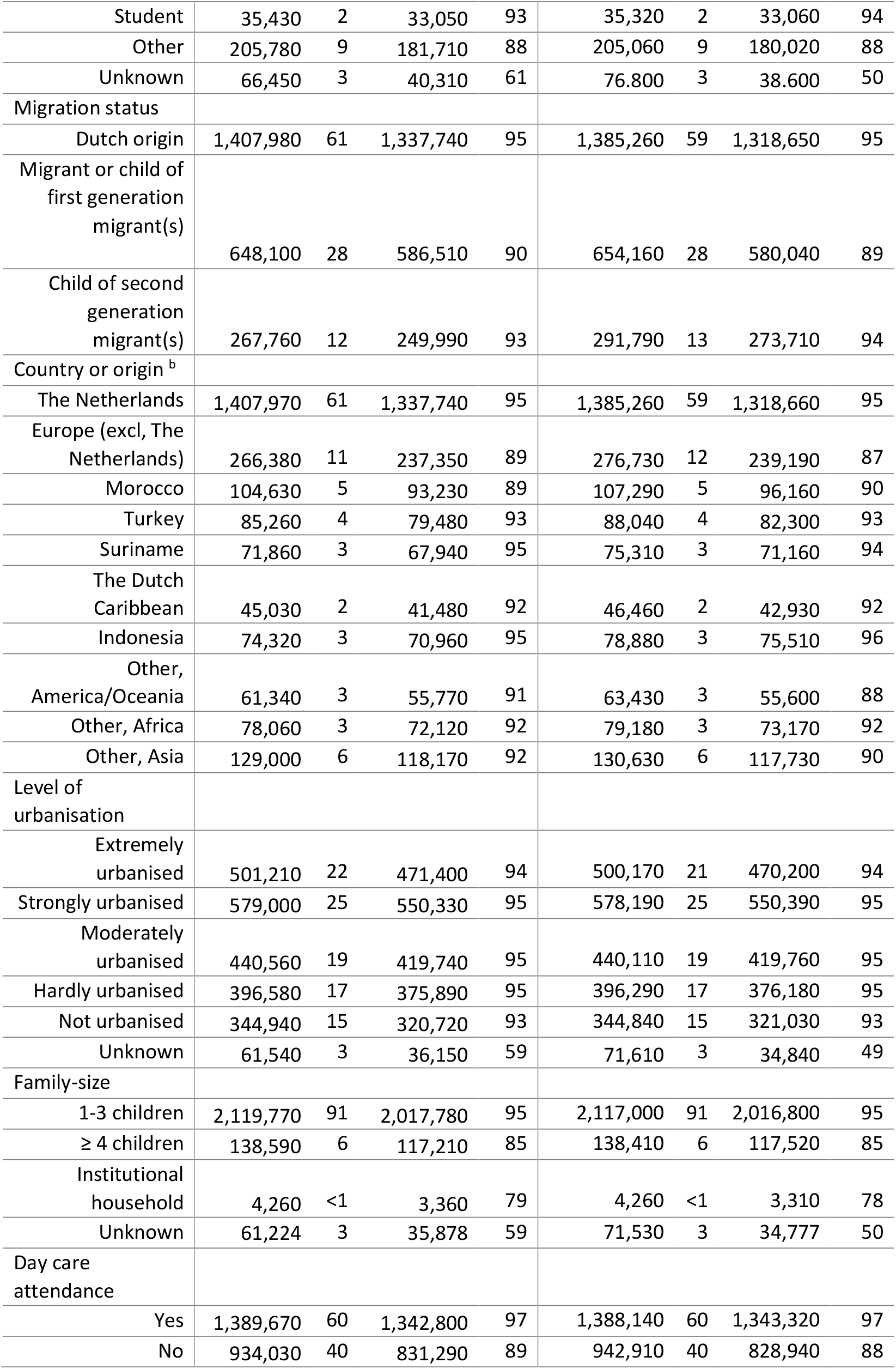
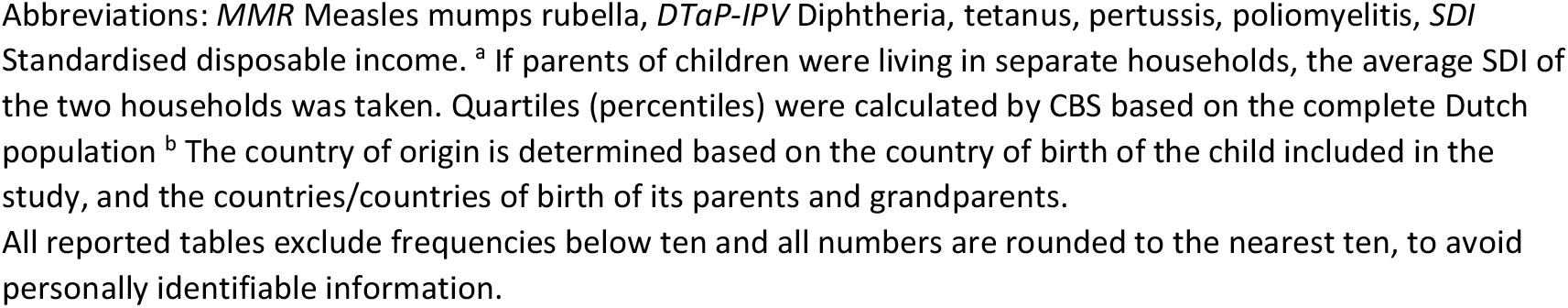
MMR and DTaP-IPV vaccination uptake at two years of age by sociodemographic characteristics for children in the Netherlands, born in 2008-2020 (n = 2,323,838 for MMR and n = 2,331,199 for DTaP-IPV).

### Sociodemographic factors associated with vaccination coverage over time

Overall, crude MMR vaccination coverage declined from 95% in cohort 2008 to 89% in cohort 2020, and a decrease was observed across all sociodemographic groups (Figure 1).

**Figure 1.**
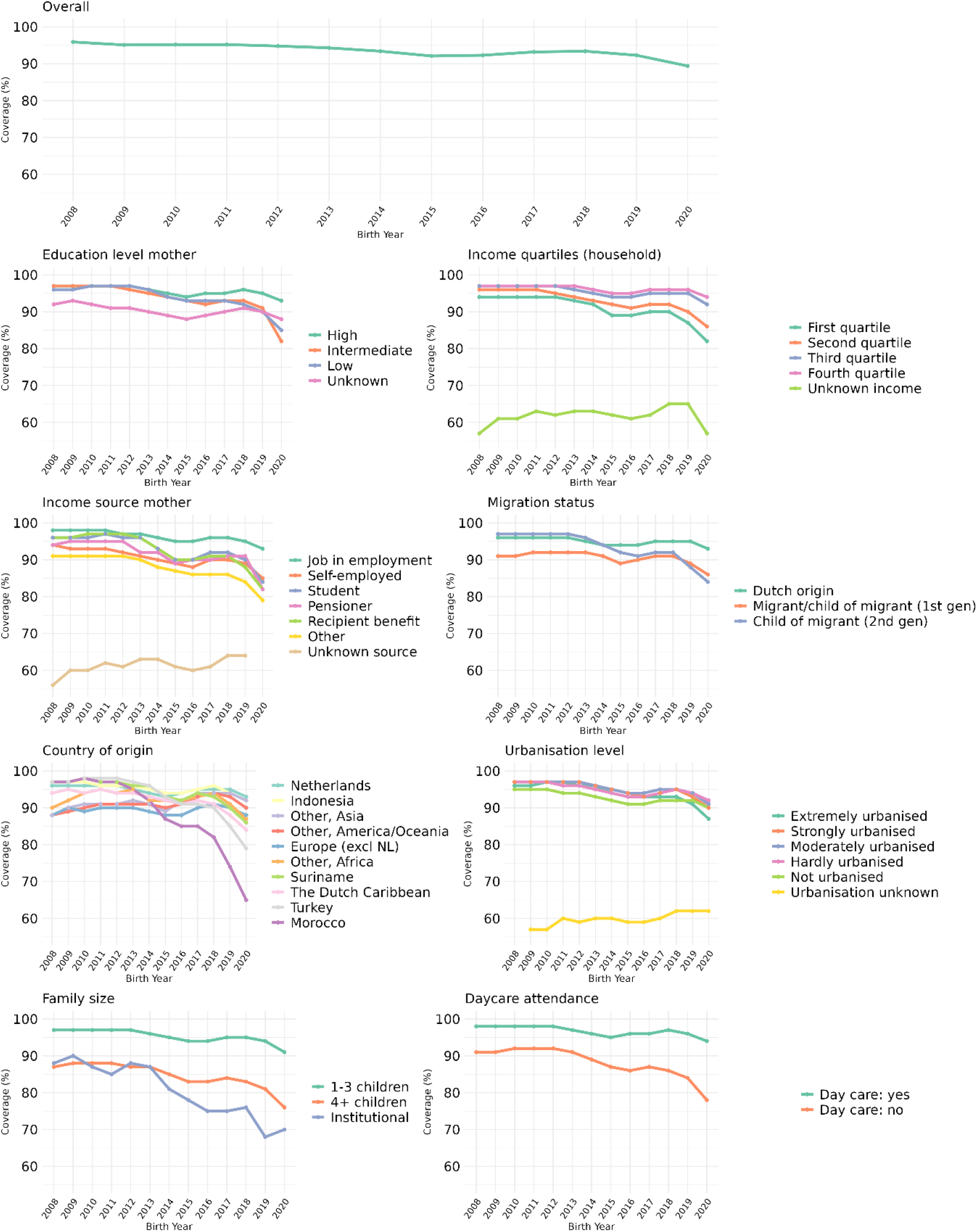
Unadjusted MMR vaccination coverage (%) at 2 years of age per birth cohort overall, and by maternal education level, household income, income source of the mother, migration status, country of origin, urbanisation level, family size and day-care attendance. The vaccination coverage for cohort 2020 may be underestimated due to informed consent.

Vaccination coverage was not associated with maternal educational level in 2008 (97%) but this changed over time: children with mothers who had higher education levels maintained relatively high coverage over time (94% in cohort 2020) while it decreased to 85% in cohort 2020 for children with mothers with low educational level. Lower vaccination coverage was consistently present among children in lowest income quartile households (94% in cohort 2008 and 82% in cohort 2020) and those with self-employed mothers (94% in cohort 2008 and 85% in cohort 2020). In the 2008 cohort, children of second-generation migrants (at least one grandparent born abroad) had higher coverage compared to the combined group of migrant children (born abroad themselves) and children of first-generation migrant (at least one parent born abroad) (97% and 81%, respectively). Coverage declined faster in children of second-generation migrants over time so that in cohort 2020, coverage (84%) was lower compared to the combined group of migrant children and children of first-generation migrants (86%). Children of Dutch origin had relatively high vaccination coverage (96% in cohort 2008-2012 and 93% in in cohort 2020) compared to Dutch children of non-Dutch origin. Specifically Dutch children of Moroccan and Turkish origin had historically higher coverage, compared to children of Dutch origin, but experienced a steeper decline in coverage from cohort 2014 onwards (97% in cohort 2008 to 65% in cohort 2020 and 97% in cohort 2008 to 79% in cohort 2020, for Dutch children of Moroccan and Turkish origin, respectively). Within the group of Dutch children of Moroccan origin, since cohort 2013, coverage was lower for children of second-generation migrants compared to the combined group of migrant children and children of first-generation migrants (58% and 71% in cohort 2020, respectively. Within the group of Dutch children of Turkish origin, only in cohort 2020, coverage was slightly lower for children of second-generation migrants compared to the combined group of migrant children and children of first-generation migrants (84% and 86%, respectively). Geographical factors also played a role, with children living in non-urbanized areas consistently showing lowest coverage until cohort 2019, after which the coverage of children in extremely urbanized areas was lowest (87% in cohort 2020). Additionally, lower coverage was consistently observed in children living in large families (76% in cohort 2020), children living in institutional households (70% in cohort 2020), and children not attending day care (78% in cohort 2020).

DTaP-IPV vaccination coverage decreased from 95% in cohort 2008 to 90% in cohort 2020. The DTaP-IPV coverage by country of origin shows some slight differences compared to MMR coverage, with Dutch children from European origin (excluding the Netherlands), ‘Other, America/Oceania’ and ‘Other, Asia’ having lower coverage in earlier cohorts (2008-2014). Aside from this, similar trends were observed in the crude DTaP-IPV vaccination coverage over time (Supplementary Figure S2).

### Sociodemographic factors associated with vaccination coverage

Figure 2 shows the results of the univariable and multivariable regression analysis for MMR vaccination and corresponding rate ratios (unadjusted and adjusted). The figure shows variations in the RR across different sociodemographic groups. Overall, the strongest associations with lower vaccination coverage in adjusted analyses were related to larger family size (≥4 children) (aRR: 0.91, living in an institutional household (aRR: 0.95) and not-attending day care (aRR: 0.96). Maternal education level overall showed a nuanced effect: while lower education was associated with lower vaccination coverage in the unadjusted model (RR: 0.97), the adjusted model indicates minimal difference in children from mothers with high education level and children from mothers without high education level (aRR: 1.00). With regard to the economic variables: lower standard disposable income (SDI) of the household was associated with lower vaccination coverage in the adjusted model, particularly in the first quartile (aRR: 0.98) as well as having a self-employed mother and a mother with ‘other’ income (aRR: 0.95 and aRR: 0.96, respectively). The association between country of origin and vaccine coverage varied: Dutch children of Moroccan origin had lower coverage compared to children of Dutch origin (aRR: 0.97), while Dutch children of Turkish, African, and Asian origin showed higher coverage compared to children of Dutch origin (aRR: 1.01 for Turkey; aRR: 1.02 for Africa and aRR: 1.03 for Asia). Living in hardly to extremely urbanised areas, compared to non-urbanised areas, overall was positively associated with MMR vaccination coverage (aRR: 1.02 for strongly urbanised areas).

**Figure 2.**
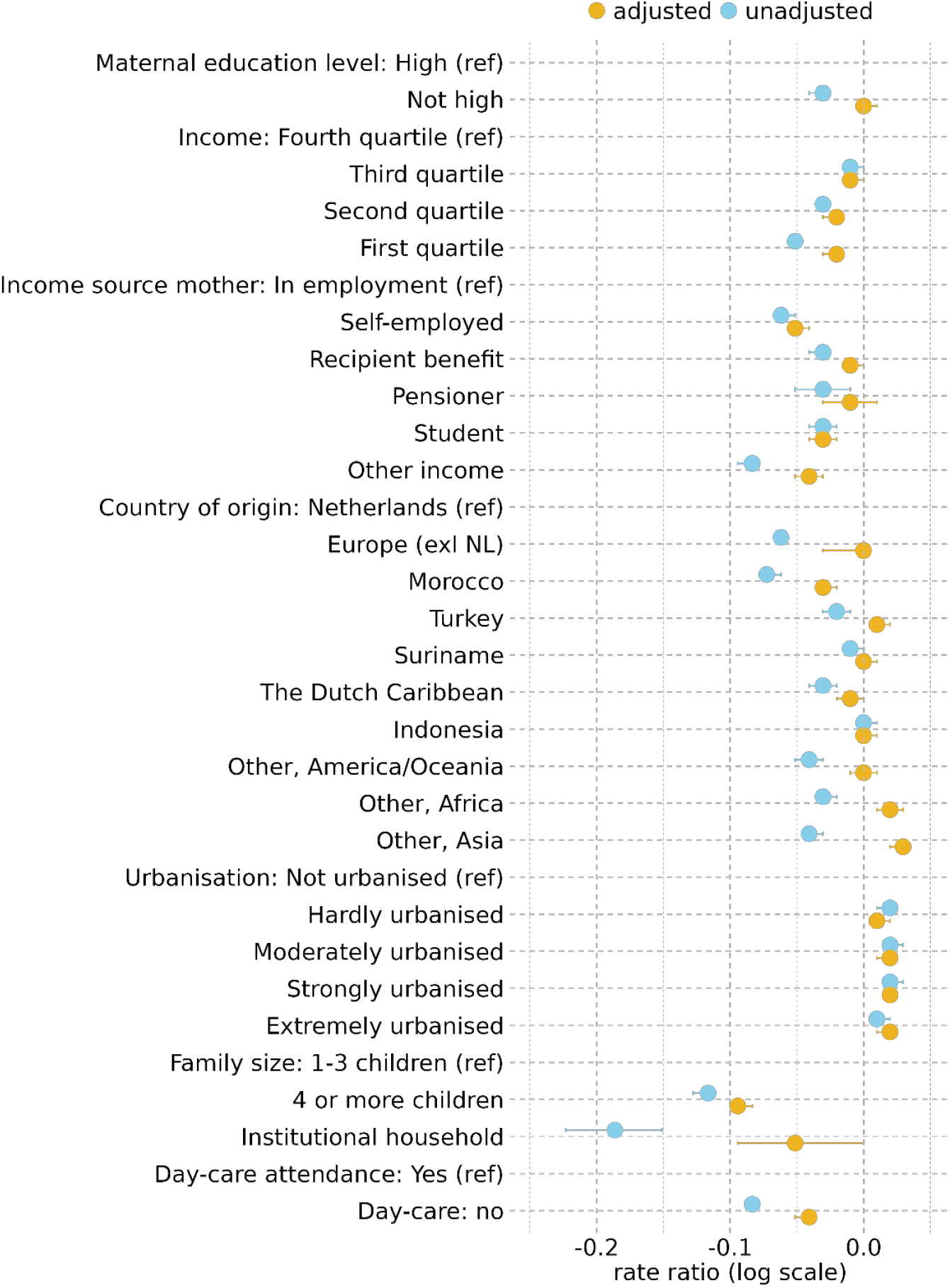
Unadjusted and adjusted rate ratios for MMR vaccination coverage by sociodemographic factors. In addition to all the variables listed, the RRs are also adjusted for birth cohort.

With regard to DTaP-IPV vaccination coverage, we found similar results (Supplementary Figure S3).

### Changes in sociodemographic factors associated with vaccination coverage over time

The multivariable regression model including interaction terms for birth cohort revealed significant variations in MMR vaccination coverage over time (the extensive model can be found in Supplementary Table S4). Table 2 presents the relative changes in vaccination coverage across sociodemographic groups relative to cohort 2008, calculated based on the estimates from the model and adjusted for baseline coverage and all sociodemographic variables. Country of origin played a significant role, with Dutch children of Moroccan origin having the most pronounced decline in coverage compared to Dutch children of Dutch origin, with 4% lower coverage in cohort 2009 and 25% lower coverage in cohort 2020. Dutch children of Turkish origin also showed a substantial decrease in coverage, with 4% lower coverage in cohort 2009 and 12% lower coverage in cohort 2020. Other notable declines were observed among Dutch children of Dutch Caribbean origin (−9% in cohort 2020) and Dutch children of Surinamese origin (−7% in cohort 2020). Day care attendance and family size was also significantly associated with differential changes in vaccination coverage. Children not attending day care had increasingly lower coverage compared to children attending day care (−3% in cohort 2009 to -12% in cohort 2020). No significant differential changes in vaccination coverage over time were found for maternal educational level, urbanisation level, and household size. Children living in families with four or more children however did have consistently lower vaccination coverage compared to children living in smaller families with the largest difference in cohort 2020 (−12%). However, these differences have remained stable over time. Socioeconomic disparities were observed and became slightly wider in cohort 2020, with consistently lower coverage among children of self-employed mothers compared to children of mothers with a job in employment (−8% in cohort 2020) and lower coverage among children in the lowest income households compared to the highest income households (−7% in cohort 2020).

**Table 2.** Adjusted relative change (%) in MMR vaccination coverage compared to the reference cohort 2008 for each stratum of the covariates, based on the rate ratios derived from the multivariable regression model with interaction terms for birth cohort

|  | Birth cohort |  |  |  |  |  |  |  |  |  |  |  |
| --- | --- | --- | --- | --- | --- | --- | --- | --- | --- | --- | --- | --- |
|  | 2009 | 2010 | 2011 | 2012 | 2013 | 2014 | 2015 | 2016 | 2017 | 2018 | 2019 | 2020 |
| Education level mother (ref: high) |  |  |  |  |  |  |  |  |  |  |  |  |
| Not high | 0,13 | 0,29 | 0,05 | -0,38 | -0,41 | -1,01 | -1,51 | -1,34 | -1,02 | -1,10 | -0,42 | -2,69 |
| SDI quartiles (ref: fourth quartile) |  |  |  |  |  |  |  |  |  |  |  |  |
| First | -1,64 | -1,80 | -2,33 | -2,42 | -3,20 | -4,02 | -5,32 | -5,57* | -4,65 | -4,30 | -5,11* | -7,20* |
| Second | -1,00 | -1,46 | -1,80 | -2,14 | -2,99 | -4,03 | -5,05 | -5,02* | -4,12* | -4,14* | -3,97* | -5,92* |
| Third | -0,33 | -0,68 | -0,88 | -1,27 | -1,76 | -2,61 | -3,67 | -3,15 | -2,36 | -2,16 | -1,61 | -2,87 |
| Income source mother (ref: job in employment) |  |  |  |  |  |  |  |  |  |  |  |  |
| Self-employed | -4,04 | -4,21 | -4,42 | -5,05 | -5,92 | -7,31 | -8,11 | -7,84* | -5,84 | -6,22 | -5,80 | -8,05* |
| Benefit recipient | -0,76 | -0,13 | -0,72 | -0,74 | -1,32 | -3,06 | -4,75 | -2,94 | -1,38 | -0,95 | -0,35 | -2,15 |
| Pensioner | -1,26 | -1,22 | -2,05 | -2,66 | -4,70 | -4,62 | -7,23 | -3,77 | -3,59 | -2,61 | 0,11 | -5,63 |
| Student | -2,31 | -1,82 | -2,30 | -2,83 | -2,81 | -5,07 | -7,95 | -6,78 | -4,40 | -4,39 | -4,52 | -7,95 |
| Other | -5,15 | -5,05 | -5,10 | -5,55 | -5,71 | -6,57 | -6,80 | -5,34 | -4,82 | -4,80 | -2,64 | -2,10* |
| Country of origin (ref: The Netherlands) |  |  |  |  |  |  |  |  |  |  |  |  |
| Europe excl. NL | -0,57 | -0,61 | -0,80 | -1,12 | -1,61 | -2,25 | -2,99 | -2,79 | -1,92 | -1,84 | -1,36 | -3,16 |
| Indonesia | 0,37 | 0,36 | -0,30 | -0,20 | -0,59 | -1,43 | -2,03 | -1,65 | -0,66 | -1,00 | -1,24 | -2,62 |
| Morocco | 4,05 | 4,78 | 3,73 | 3,33 | 1,90 | -0,58 | -5,11* | -7,04* | -7,25* | -10,05* | -17,74* | -25,41* |
| Other, Africa | 0,18 | 1,15 | 2,03 | 1,15 | 1,86 | -0,33 | 0,20 | 0,63 | 1,00 | 1,35 | 1,00 | -1,56 |
| Other, America/Oceania | -1,25 | -0,69 | -1,14 | -1,75 | -1,52 | -2,49 | -3,06 | -2,04 | -1,19 | -1,23 | -1,07 | -2,61 |
| Other, Asia | 1,32 | 2,42 | 2,03 | 1,55 | 1,42 | 0,14 | -1,87 | 0,46 | 2,23 | 2,28 | 3,30 | 2,03 |
| Suriname | 1,15 | 1,44 | 1,02 | 1,03 | 0,52 | 0,21 | -1,59 | -2,57 | -1,23 | -2,66 | -3,83* | -7,05* |
| The Dutch Caribbean | 0,84 | 0,51 | 0,56 | -0,07 | -0,08 | -0,77 | -1,87 | -3,35 | -2,11 | -3,82 | -5,02 | -8,70* |
| Turkey | 3,68 | 4,31 | 3,77 | 3,66 | 3,61 | 2,56 | 0,40 | -1,17 | -1,48 | -2,47* | -7,19* | -11,55* |
| Level of urbanization (ref: not urbanized) |  |  |  |  |  |  |  |  |  |  |  |  |
| Extremely | 0,74 | 0,65 | 0,67 | 0,52 | 0,58 | -0,22 | -0,18 | 0,51 | 0,71 | 1,18 | 1,28 | -0,51 |
| Strongly | 1,19 | 1,25 | 1,14 | 0,91 | 0,44 | 0,00 | -0,24 | 0,35 | 0,93 | 1,68 | 1,67 | 0,66 |
| Moderately | 1,06 | 1,23 | 1,01 | 0,91 | 0,51 | -0,04 | -0,36 | 0,44 | 0,65 | 0,90 | 1,31 | 0,24 |
| Hardly | 1,04 | 0,91 | 0,67 | 0,32 | -0,49 | -1,05 | -1,91 | -1,00 | -0,25 | 0,55 | 1,05 | -0,37 |
| Family size (ref: 1-3 children) |  |  |  |  |  |  |  |  |  |  |  |  |
| ≥ 4 children | -8,73 | -8,84 | -8,70 | -9,66 | -9,15 | -10,78 | -11,30 | -10,09 | -8,78 | -9,77 | -9,12 | -12,00 |
| Institutional | -0,73 | -4,21 | -5,76 | -2,33 | -3,31 | -8,66 | -8,66 | -7,51 | -6,31 | -3,97 | -9,21 | -3,99 |
| Day care attendance (ref: day care yes) |  |  |  |  |  |  |  |  |  |  |  |  |
| Day care no | -2,61 | -2,91 | -3,18 | -3,70 | -4,38 | -5,67 | -6,55* | -7,13* | -7,20* | -8,11* | -8,60* | -11,98* |

Abbreviations: *MMR* Measles mumps rubella, *SDI* Standardised disposable income. \* Significant values (p<0.05)

We performed a sensitivity analysis including missing values which showed minimal changes in the model estimates (Supplementary Table S4). Regarding DTaP-IPV vaccination, similar trends were observed. The multivariable model can be found in Supplementary Table 3 and the adjusted relative changes in DTaP-IPV vaccination over time can be found in Supplementary Table S4.

## Discussion

Our study revealed a significant decline in MMR and DTaP-IPV vaccination coverage in the Netherlands from birth cohort 2008 to 2020, with a more pronounced and faster decline observed in specific sociodemographic subgroups. This trend poses an increased risk for outbreaks of vaccine preventable diseases and indicates an increasing inequity in vaccination coverage. This study presents an application of the situation analysis phase of the WHO TIP framework [13-15]. Further research within the SocioVax program will explore barriers and drivers of vaccination in subgroups with lower coverage [19].

The general decline in vaccination coverage observed in the Netherlands is consistent with trends reported in other countries [20, 21]. The coverage for the first-dose measles containing vaccine in the EU/EEA region decreased from 95% in 2018 to 92% in 2022 [22]. General explanations for the decline in vaccination coverage include distrust in vaccines and governmental organisations, misinformation and the influence of social media[23-25]. Studies link scepticism towards governmental policies and lack of confidence in vaccines to reduced vaccination uptake [25]. The COVID-19 pandemic may have exacerbated these issues. Trust in governmental institutions among people in the Netherlands in 2022 was below the pre-pandemic level, especially among lower educated individuals, and was associated with lower COVID-19 vaccine uptake [26]. Parental perceptions about childhood vaccination also became slightly more negative post-pandemic [27], though coverage had already been declining earlier. Also, social media plays a role in the spread of misinformation. Research on Dutch Twitter activity in 2019, showed how anti-vaccine narratives gained attention and influenced public opinion [24]. Due to selective exposure, individuals with more sceptical attitudes towards vaccination are more likely to encounter negative messages about vaccination [24]. The anti-vaccination message on the internet is much less restrained than in other media, leading to a higher likelihood of the public making uninformed decisions about vaccination [23].

Crucially, our findings indicate that specific sociodemographic groups experience a more pronounced and faster decrease in vaccination coverage. The most substantial relative declines were observed among Dutch children of non-Dutch origin, children not attending day care, those with self-employed mothers and children in the lowest income households. More specifically, we found notable declines in vaccination coverage among Dutch children with a migration background, specifically children of Moroccan, Turkish, Dutch-Caribbean and Surinamese origin, also after adjusting for other variables. These findings are consistent with earlier studies in the Netherlands on NIP coverage at national and municipal levels [9, 12]. The potential social clustering of unvaccinated children within these populations poses public health risks by increasing outbreak potential [28]. Several qualitative studies have been conducted to provide further insights in reasons behind lower vaccination coverage among Dutch children with a migration background. The results of a focus group study indicated difficulty in understanding information about the NIP and distance to vaccination location (i.e. Child Welfare Centres) as important barriers to vaccination [29]. Other reasons may be equally or more important given that Morocco currently is experiencing a large measles epidemic, following declining vaccination rates after the COVID-19 pandemic [30]. Unlike objections in the Orthodox Reformed population [4], no clear Islamic religious objection to vaccination has been identified [29, 31, 32]. While some studies mention violation of dietary laws (e.g. non-halal-based vaccines), we found no difference in MMR (where one of the two widely used vaccine contains porcine gelatine and is therefore non-halal) and DTaP-IPV (halal vaccines) coverage.

Moreover, children not attending day care showed lower vaccination coverage. A Canadian systematic review found varying associations between day care attendance and infant vaccination uptake [33]. In the Dutch context, even though vaccination is not required for day care attendance, parents are often asked about their child’s vaccination status, which may influence their vaccination decision. As day care attendance is likely associated with other sociodemographic variables, there may be residual confounding after adjusting for available information. Due to close contacts among children, day care centres can be significant hotspots for infectious disease transmission, thus the higher vaccination coverage in these settings is advantageous for preventing outbreaks.

The socioeconomic disparities observed in our study are consistent with findings from previous research. Systematic reviews reported persisting lower routine childhood vaccination coverage among children with lower parental socioeconomic status [34, 35]. Contrary to low- and middle-income countries where (financial) access is often a barrier to vaccine uptake in lower socioeconomic groups, in high-income countries economic disparities may be more related to issues of perceived risks (both of the diseases against which vaccines protect as well as of the vaccines themselves), trust in the government and in health care professionals, and vaccine confidence [34, 35]. However, the effects of factors on vaccine uptake among people from lower socioeconomic positions need further research to investigate underlying causes to address these disparities.

While we described some findings about underlying factors from research in the general population it is unclear whether, and to what extent, these same factors play a role in the vaccination uptake of people from these specific subgroups. In co-creating with specific subgroups, further research is developed to explore barriers and drivers of vaccination in subgroups with lower coverage [19].

Low coverage becomes a significant problem especially when unvaccinated individuals cluster. For example, children with similar migration backgrounds may cluster within households, families, schools and geographically (e.g. larger cities). This social clustering potentially creates environments where infections spread more easily. Therefore, further research to investigate networks of unvaccinated and nonimmune individuals is needed to assess the risk of spread of vaccine-preventable diseases.

A strength of our study is the use of nationwide registry data and the possibility to link vaccination data at individual level, avoiding ecological fallacies that may result from aggregated data. The results provide a comprehensive understanding of factors influencing changes in vaccination coverage and help identify potential risk groups for disease outbreaks. A limitation is the requirement for informed consent for a subset of children born in 2020. Children who could not be linked to the vaccination database were categorised as unvaccinated. This may be differential between subgroups (no data on characteristics of parents not providing consent is available). However, the effect is limited. For MMR vaccination, only children born in November and December (approximately ⅙ of the 2020 cohort) were scheduled for vaccination after the introduction of the informed consent in January 2022. For the DTaP-IPV vaccination, all children born in 2020 were scheduled for vaccination before the introduction of the informed consent. The percentage of parents not providing informed consent for vaccination was 3.1% for DTaP-IPV and 4.5% for MMR in 2022 [36]. Finally, the 2019 and 2020 cohorts were the first to be vaccinated during the COVID-19 pandemic, which may have caused delays. Nonetheless, when vaccination status was assessed at age 3 instead of 2 years, overall coverage for these cohorts remained unchanged.

## Conclusion

In conclusion, we found a significant decline in MMR and DTaP-IPV childhood vaccination coverage in the Netherlands, with increasing disparities between sociodemographic strata. The potential social clustering of these unvaccinated children increases the risk of localised outbreaks. Future research should focus on understanding the underlying reasons for non-uptake of childhood vaccinations, focusing on populations with lower uptake, according to the WHO TIP framework. Results from social scientific research can inform interventions with the aim to increase childhood vaccination coverage.

## Data Availability

All data is available within CBS Microdata and can be made available under strict conditions.

