## Supplementary material for "The decrease in childhood vaccination coverage in the Netherlands from birth cohort 2008 to 2020 and its sociodemographic determinants"

### Supplementary files

**Supplementary Table S1. Distribution sociodemographic variables and MMR and DTaP-IPV vaccination coverage per birth cohort**

| Category | MMR population | Vaccinated | | DTaP-IPV population | Vaccinated | | Category | MMR population | Vaccinated | | DTaP-IPV population | Vaccinated | |
|---|---|---|---|---|---|---|---|---|---|---|---|---|---|
| | N | N | % | N | N | % | | N | N | % | N | N | % |
| Education level mother high | | | | | | | Country of origin: The Netherlands | | | | | | |
| Cohort 2008 | 55314 | 53742 | 97 | 55265 | 53619 | 97 | Cohort 2008 | 118975 | 114700 | 96 | 118968 | 114613 | 96 |
| Cohort 2009 | 58417 | 56784 | 97 | 58371 | 56674 | 97 | Cohort 2009 | 118260 | 114019 | 96 | 118263 | 113889 | 96 |
| Cohort 2010 | 60797 | 59028 | 97 | 60762 | 58905 | 97 | Cohort 2010 | 116353 | 112059 | 96 | 116365 | 111827 | 96 |
| Cohort 2011 | 67581 | 65501 | 97 | 67542 | 65440 | 97 | Cohort 2011 | 112850 | 108601 | 96 | 112844 | 108497 | 96 |
| Cohort 2012 | 67717 | 65440 | 97 | 67658 | 65372 | 97 | Cohort 2012 | 108811 | 104285 | 96 | 108804 | 104218 | 96 |
| Cohort 2013 | 67827 | 65115 | 96 | 67780 | 65335 | 96 | Cohort 2013 | 105450 | 100399 | 95 | 105445 | 100864 | 96 |
| Cohort 2014 | 70118 | 66677 | 95 | 70060 | 67062 | 96 | Cohort 2014 | 106602 | 100614 | 94 | 106617 | 101408 | 95 |
| Cohort 2015 | 69658 | 65574 | 94 | 69604 | 66144 | 95 | Cohort 2015 | 102457 | 95762 | 93 | 102444 | 96678 | 94 |
| Cohort 2016 | 71439 | 67530 | 95 | 71381 | 67673 | 95 | Cohort 2016 | 102144 | 95820 | 94 | 102136 | 96169 | 94 |
| Cohort 2017 | 71475 | 68213 | 95 | 71430 | 68078 | 95 | Cohort 2017 | 100069 | 94617 | 95 | 100063 | 94538 | 94 |
| Cohort 2018 | 72076 | 68993 | 96 | 72031 | 68821 | 96 | Cohort 2018 | 97802 | 92814 | 95 | 97783 | 92759 | 95 |
| Cohort 2019 | 73178 | 69616 | 95 | 73148 | 69522 | 95 | Cohort 2019 | 97499 | 92343 | 95 | 97475 | 92208 | 95 |
| Cohort 2020 | 73790 | 68789 | 93 | 73753 | 69406 | 94 | Cohort 2020 | 97023 | 89821 | 93 | 96990 | 90830 | 94 |
| Education level mother intermediate | | | | | | | Country of origin: Europe (excl. NL) | | | | | | |
| Cohort 2008 | 40155 | 38877 | 97 | 40120 | 38698 | 96 | Cohort 2008 | 20717 | 18316 | 88 | 21108 | 17658 | 84 |
| Cohort 2009 | 43837 | 42431 | 97 | 43810 | 42223 | 96 | Cohort 2009 | 21197 | 18990 | 90 | 21613 | 18304 | 85 |
| Cohort 2010 | 47775 | 46229 | 97 | 47745 | 46018 | 96 | Cohort 2010 | 21609 | 19320 | 89 | 21978 | 18815 | 86 |

| | | | | | | | | | | | | | |
|---|---|---|---|---|---|---|---|---|---|---|---|---|---|
| Cohort 2011 | 51776 | 50083 | 97 | 51752 | 49919 | 96 | Cohort 2011 | 20861 | 18709 | 90 | 21243 | 18206 | 86 |
| Cohort 2012 | 53284 | 51260 | 96 | 53251 | 51121 | 96 | Cohort 2012 | 20764 | 18593 | 90 | 21172 | 18151 | 86 |
| Cohort 2013 | 54219 | 51701 | 95 | 54197 | 51920 | 96 | Cohort 2013 | 20453 | 18306 | 90 | 20827 | 17994 | 86 |
| Cohort 2014 | 58034 | 54628 | 94 | 58004 | 55115 | 95 | Cohort 2014 | 21000 | 18696 | 89 | 21422 | 18495 | 86 |
| Cohort 2015 | 58154 | 53904 | 93 | 58121 | 54522 | 94 | Cohort 2015 | 20800 | 18378 | 88 | 21171 | 18306 | 86 |
| Cohort 2016 | 60073 | 55373 | 92 | 60042 | 55760 | 93 | Cohort 2016 | 21007 | 18574 | 88 | 21432 | 18570 | 87 |
| Cohort 2017 | 60144 | 55808 | 93 | 60118 | 55836 | 93 | Cohort 2017 | 20897 | 18703 | 90 | 21245 | 18540 | 87 |
| Cohort 2018 | 59755 | 55305 | 93 | 59730 | 55458 | 93 | Cohort 2018 | 20863 | 18915 | 91 | 21211 | 18764 | 88 |
| Cohort 2019 | 60875 | 55291 | 91 | 60863 | 55713 | 92 | Cohort 2019 | 20862 | 18804 | 90 | 21203 | 18859 | 89 |
| Cohort 2020 | 60872 | 52946 | 87 | 60851 | 54039 | 89 | Cohort 2020 | 20832 | 18267 | 88 | 21108 | 18531 | 88 |
| Education level mother low | | | | | | | Country of origin: Indonesia | | | | | | |
| Cohort 2008 | 19495 | 18777 | 96 | 19472 | 18591 | 95 | Cohort 2008 | 7741 | 7483 | 97 | 7738 | 7466 | 96 |
| Cohort 2009 | 20538 | 19802 | 96 | 20521 | 19545 | 95 | Cohort 2009 | 7664 | 7420 | 97 | 7665 | 7424 | 97 |
| Cohort 2010 | 20951 | 20266 | 97 | 20921 | 20042 | 96 | Cohort 2010 | 7316 | 7075 | 97 | 7321 | 7086 | 97 |
| Cohort 2011 | 23765 | 23033 | 97 | 23742 | 22866 | 96 | Cohort 2011 | 6953 | 6703 | 96 | 6952 | 6689 | 96 |
| Cohort 2012 | 23175 | 22427 | 97 | 23166 | 22281 | 96 | Cohort 2012 | 6601 | 6356 | 96 | 6610 | 6364 | 96 |
| Cohort 2013 | 21683 | 20822 | 96 | 21647 | 20815 | 96 | Cohort 2013 | 6248 | 5976 | 96 | 6250 | 6002 | 96 |
| Cohort 2014 | 21883 | 20686 | 95 | 21812 | 20838 | 96 | Cohort 2014 | 6094 | 5767 | 95 | 6100 | 5825 | 95 |
| Cohort 2015 | 20424 | 18911 | 93 | 20374 | 19123 | 94 | Cohort 2015 | 5842 | 5466 | 94 | 5849 | 5518 | 94 |
| Cohort 2016 | 19764 | 18302 | 93 | 19747 | 18456 | 93 | Cohort 2016 | 5420 | 5120 | 94 | 5423 | 5148 | 95 |
| Cohort 2017 | 17925 | 16641 | 93 | 17898 | 16648 | 93 | Cohort 2017 | 5170 | 4927 | 95 | 5178 | 4938 | 95 |
| Cohort 2018 | 17291 | 15999 | 93 | 17279 | 16040 | 93 | Cohort 2018 | 4905 | 4693 | 96 | 4907 | 4690 | 96 |
| Cohort 2019 | 15621 | 14069 | 90 | 15612 | 14239 | 91 | Cohort 2019 | 4608 | 4349 | 94 | 4617 | 4389 | 95 |
| Cohort 2020 | 14364 | 12173 | 85 | 14354 | 12500 | 87 | Cohort 2020 | 4260 | 3927 | 92 | 4267 | 3971 | 93 |
| Education level mother unknown | | | | | | | Country of origin: Morocco | | | | | | |
| Cohort 2008 | 75891 | 69672 | 92 | 76539 | 68438 | 89 | Cohort 2008 | 8591 | 8366 | 97 | 8587 | 8279 | 96 |
| Cohort 2009 | 67652 | 62153 | 92 | 68390 | 61023 | 89 | Cohort 2009 | 8519 | 8274 | 97 | 8518 | 8199 | 96 |
| Cohort 2010 | 60169 | 55002 | 91 | 60865 | 54063 | 89 | Cohort 2010 | 8487 | 8278 | 98 | 8484 | 8216 | 97 |
| Cohort 2011 | 41636 | 37316 | 90 | 42393 | 36575 | 86 | Cohort 2011 | 8438 | 8216 | 97 | 8451 | 8190 | 97 |

| | | | | | | | | | | | | | |
|---|---|---|---|---|---|---|---|---|---|---|---|---|---|
| Cohort 2012 | 36309 | 32047 | 88 | 37116 | 31496 | 85 | Cohort 2012 | 8366 | 8095 | 97 | 8380 | 8091 | 97 |
| Cohort 2013 | 31681 | 27854 | 88 | 32409 | 27446 | 85 | Cohort 2013 | 8142 | 7749 | 95 | 8147 | 7802 | 96 |
| Cohort 2014 | 28867 | 25043 | 87 | 29633 | 24780 | 84 | Cohort 2014 | 8330 | 7658 | 92 | 8342 | 7815 | 94 |
| Cohort 2015 | 25951 | 22096 | 85 | 26586 | 21947 | 83 | Cohort 2015 | 8201 | 7160 | 87 | 8197 | 7324 | 89 |
| Cohort 2016 | 24170 | 20749 | 86 | 24851 | 20624 | 83 | Cohort 2016 | 8255 | 7015 | 85 | 8261 | 7133 | 86 |
| Cohort 2017 | 22840 | 19960 | 87 | 23482 | 19801 | 84 | Cohort 2017 | 8014 | 6826 | 85 | 8015 | 6858 | 86 |
| Cohort 2018 | 21636 | 19147 | 88 | 22196 | 19036 | 86 | Cohort 2018 | 7830 | 6460 | 83 | 7830 | 6534 | 83 |
| Cohort 2019 | 21345 | 18841 | 88 | 21912 | 18891 | 86 | Cohort 2019 | 8074 | 5990 | 74 | 8071 | 6237 | 77 |
| Cohort 2020 | 20472 | 17603 | 86 | 20893 | 17905 | 86 | Cohort 2020 | 8007 | 5239 | 65 | 8006 | 5480 | 68 |
| Standardised disposable income household: first quartile | | | | | | | Country of origin: Turkey | | | | | | |
| Cohort 2008 | 29025 | 27353 | 94 | 28978 | 27030 | 93 | Cohort 2008 | 6806 | 6589 | 97 | 6821 | 6504 | 95 |
| Cohort 2009 | 28916 | 27197 | 94 | 28844 | 26831 | 93 | Cohort 2009 | 6552 | 6363 | 97 | 6559 | 6302 | 96 |
| Cohort 2010 | 28991 | 27382 | 94 | 28925 | 27075 | 94 | Cohort 2010 | 6789 | 6640 | 98 | 6793 | 6571 | 97 |
| Cohort 2011 | 27754 | 26167 | 94 | 27697 | 26002 | 94 | Cohort 2011 | 6668 | 6533 | 98 | 6682 | 6502 | 97 |
| Cohort 2012 | 26877 | 25271 | 94 | 26838 | 25112 | 94 | Cohort 2012 | 6750 | 6596 | 98 | 6765 | 6584 | 97 |
| Cohort 2013 | 25035 | 23304 | 93 | 24969 | 23263 | 93 | Cohort 2013 | 6501 | 6317 | 97 | 6505 | 6307 | 97 |
| Cohort 2014 | 24806 | 22720 | 92 | 24730 | 22820 | 92 | Cohort 2014 | 6994 | 6688 | 96 | 6997 | 6713 | 96 |
| Cohort 2015 | 22868 | 20455 | 89 | 22782 | 20626 | 91 | Cohort 2015 | 6803 | 6330 | 93 | 6784 | 6402 | 94 |
| Cohort 2016 | 22806 | 20298 | 89 | 22735 | 20382 | 90 | Cohort 2016 | 7112 | 6504 | 91 | 7097 | 6556 | 92 |
| Cohort 2017 | 21171 | 18963 | 90 | 21096 | 18817 | 89 | Cohort 2017 | 6895 | 6288 | 91 | 6883 | 6308 | 92 |
| Cohort 2018 | 20718 | 18549 | 90 | 20673 | 18540 | 90 | Cohort 2018 | 6839 | 6181 | 90 | 6828 | 6233 | 91 |
| Cohort 2019 | 19229 | 16641 | 87 | 19223 | 16834 | 88 | Cohort 2019 | 6812 | 5780 | 85 | 6813 | 5955 | 87 |
| Cohort 2020 | 18502 | 15082 | 82 | 18442 | 15414 | 84 | Cohort 2020 | 6527 | 5174 | 79 | 6512 | 5363 | 82 |
| Standardised disposable income household: second quartile | | | | | | | Country of origin: Suriname | | | | | | |

| | | | | | | | | | | | | | |
|---|---|---|---|---|---|---|---|---|---|---|---|---|---|
| Cohort 2008 | 58480 | 56286 | 96 | 58438 | 55989 | 96 | Cohort 2008 | 6074 | 5915 | 97 | 6077 | 5812 | 96 |
| Cohort 2009 | 57046 | 54864 | 96 | 57025 | 54642 | 96 | Cohort 2009 | 6187 | 6015 | 97 | 6188 | 5933 | 96 |
| Cohort 2010 | 54402 | 52197 | 96 | 54365 | 51910 | 95 | Cohort 2010 | 6236 | 6080 | 97 | 6237 | 6018 | 96 |
| Cohort 2011 | 51616 | 49462 | 96 | 51598 | 49257 | 95 | Cohort 2011 | 6029 | 5869 | 97 | 6038 | 5846 | 97 |
| Cohort 2012 | 49035 | 46768 | 95 | 49003 | 46585 | 95 | Cohort 2012 | 5846 | 5680 | 97 | 5857 | 5653 | 97 |
| Cohort 2013 | 46153 | 43581 | 94 | 46137 | 43705 | 95 | Cohort 2013 | 5758 | 5545 | 96 | 5760 | 5552 | 96 |
| Cohort 2014 | 47911 | 44643 | 93 | 47889 | 45000 | 94 | Cohort 2014 | 5678 | 5436 | 96 | 5681 | 5452 | 96 |
| Cohort 2015 | 47018 | 43119 | 92 | 46980 | 43578 | 93 | Cohort 2015 | 5708 | 5338 | 94 | 5711 | 5408 | 95 |
| Cohort 2016 | 48674 | 44501 | 91 | 48645 | 44747 | 92 | Cohort 2016 | 5721 | 5281 | 92 | 5730 | 5321 | 93 |
| Cohort 2017 | 47119 | 43384 | 92 | 47074 | 43362 | 92 | Cohort 2017 | 5557 | 5213 | 94 | 5564 | 5227 | 94 |
| Cohort 2018 | 46632 | 42836 | 92 | 46594 | 42773 | 92 | Cohort 2018 | 5468 | 5070 | 93 | 5472 | 5088 | 93 |
| Cohort 2019 | 46870 | 42352 | 90 | 46879 | 42575 | 91 | Cohort 2019 | 5485 | 4955 | 90 | 5492 | 5022 | 91 |
| Cohort 2020 | 44864 | 38765 | 86 | 44827 | 39585 | 88 | Cohort 2020 | 5490 | 4705 | 86 | 5498 | 4830 | 88 |
| Standardised disposable income household: third quartile | | | | | | | Country of origin: The Dutch Caribbean | | | | | | |
| Cohort 2008 | 56445 | 54740 | 97 | 56385 | 54556 | 97 | Cohort 2008 | 3597 | 3362 | 93 | 3604 | 3325 | 92 |
| Cohort 2009 | 57012 | 55494 | 97 | 56981 | 55253 | 97 | Cohort 2009 | 3498 | 3326 | 95 | 3508 | 3301 | 94 |
| Cohort 2010 | 57237 | 55666 | 97 | 57196 | 55467 | 97 | Cohort 2010 | 3591 | 3390 | 94 | 3596 | 3370 | 94 |
| Cohort 2011 | 57771 | 56244 | 97 | 57755 | 56122 | 97 | Cohort 2011 | 3457 | 3288 | 95 | 3482 | 3294 | 95 |
| Cohort 2012 | 57084 | 55284 | 97 | 57062 | 55153 | 97 | Cohort 2012 | 3460 | 3265 | 94 | 3483 | 3266 | 94 |
| Cohort 2013 | 56781 | 54708 | 96 | 56762 | 54872 | 97 | Cohort 2013 | 3439 | 3246 | 94 | 3459 | 3282 | 95 |
| Cohort 2014 | 58602 | 55909 | 95 | 58520 | 56206 | 96 | Cohort 2014 | 3642 | 3405 | 93 | 3652 | 3449 | 94 |
| Cohort 2015 | 58006 | 54571 | 94 | 57902 | 55017 | 95 | Cohort 2015 | 3527 | 3248 | 92 | 3526 | 3296 | 93 |
| Cohort 2016 | 58542 | 55297 | 94 | 58464 | 55447 | 95 | Cohort 2016 | 3667 | 3326 | 91 | 3681 | 3365 | 91 |
| Cohort 2017 | 59987 | 57072 | 95 | 59938 | 56987 | 95 | Cohort 2017 | 3505 | 3230 | 92 | 3514 | 3237 | 92 |
| Cohort 2018 | 59755 | 57031 | 95 | 59691 | 57015 | 96 | Cohort 2018 | 3629 | 3300 | 91 | 3634 | 3313 | 91 |
| Cohort 2019 | 60585 | 57331 | 95 | 60553 | 57396 | 95 | Cohort 2019 | 3576 | 3164 | 88 | 3585 | 3207 | 89 |
| Cohort 2020 | 60592 | 55798 | 92 | 60547 | 56468 | 93 | Cohort 2020 | 3738 | 3125 | 84 | 3733 | 3224 | 86 |

| | | | | | | | | | | | | | |
|---|---|---|---|---|---|---|---|---|---|---|---|---|---|
| Standardised disposable income household: fourth quartile | | | | | | | Country of origin: Other, Africa | | | | | | |
| Cohort 2008 | 39762 | 38652 | 97 | 39717 | 38368 | 97 | Cohort 2008 | 5522 | 4981 | 90 | 5532 | 4897 | 89 |
| Cohort 2009 | 40547 | 39409 | 97 | 40504 | 39149 | 97 | Cohort 2009 | 5615 | 5169 | 92 | 5630 | 5060 | 90 |
| Cohort 2010 | 42067 | 41001 | 97 | 42027 | 40773 | 97 | Cohort 2010 | 5860 | 5490 | 94 | 5881 | 5426 | 92 |
| Cohort 2011 | 41049 | 39943 | 97 | 41008 | 39755 | 97 | Cohort 2011 | 5864 | 5564 | 95 | 5887 | 5554 | 94 |
| Cohort 2012 | 41087 | 39862 | 97 | 41026 | 39689 | 97 | Cohort 2012 | 5941 | 5608 | 94 | 5964 | 5605 | 94 |
| Cohort 2013 | 41634 | 40244 | 97 | 41556 | 40214 | 97 | Cohort 2013 | 5794 | 5499 | 95 | 5821 | 5480 | 94 |
| Cohort 2014 | 42141 | 40325 | 96 | 42071 | 40450 | 96 | Cohort 2014 | 5954 | 5488 | 92 | 5953 | 5552 | 93 |
| Cohort 2015 | 41288 | 39221 | 95 | 41197 | 39410 | 96 | Cohort 2015 | 5789 | 5304 | 92 | 5797 | 5377 | 93 |
| Cohort 2016 | 41122 | 39226 | 95 | 41056 | 39220 | 96 | Cohort 2016 | 6031 | 5573 | 92 | 6060 | 5626 | 93 |
| Cohort 2017 | 40424 | 38902 | 96 | 40373 | 38764 | 96 | Cohort 2017 | 6163 | 5748 | 93 | 6148 | 5754 | 94 |
| Cohort 2018 | 40354 | 38880 | 96 | 40254 | 38722 | 96 | Cohort 2018 | 6721 | 6297 | 94 | 6711 | 6301 | 94 |
| Cohort 2019 | 41085 | 39390 | 96 | 41029 | 39255 | 96 | Cohort 2019 | 7015 | 6409 | 91 | 7023 | 6462 | 92 |
| Cohort 2020 | 43395 | 40645 | 94 | 43314 | 40942 | 95 | Cohort 2020 | 6784 | 5908 | 87 | 6774 | 6076 | 90 |
| Standardised disposable income household: unknown | | | | | | | Country of origin: Other, America/Oceania | | | | | | |
| Cohort 2008 | 7143 | 4037 | 57 | 7878 | 3403 | 43 | Cohort 2008 | 4524 | 4007 | 89 | 4559 | 3739 | 82 |
| Cohort 2009 | 6923 | 4206 | 61 | 7738 | 3590 | 46 | Cohort 2009 | 4566 | 4086 | 89 | 4613 | 3827 | 83 |
| Cohort 2010 | 6995 | 4279 | 61 | 7780 | 3803 | 49 | Cohort 2010 | 4636 | 4179 | 90 | 4670 | 3939 | 84 |
| Cohort 2011 | 6568 | 4117 | 63 | 7371 | 3664 | 50 | Cohort 2011 | 4724 | 4308 | 91 | 4782 | 4100 | 86 |
| Cohort 2012 | 6402 | 3989 | 62 | 7262 | 3731 | 51 | Cohort 2012 | 4642 | 4217 | 91 | 4703 | 4006 | 85 |
| Cohort 2013 | 5807 | 3655 | 63 | 6609 | 3462 | 52 | Cohort 2013 | 4641 | 4215 | 91 | 4704 | 4079 | 87 |
| Cohort 2014 | 5442 | 3437 | 63 | 6299 | 3319 | 53 | Cohort 2014 | 5002 | 4550 | 91 | 5037 | 4433 | 88 |
| Cohort 2015 | 5007 | 3119 | 62 | 5824 | 3105 | 53 | Cohort 2015 | 4944 | 4460 | 90 | 5002 | 4402 | 88 |
| Cohort 2016 | 4302 | 2632 | 61 | 5121 | 2717 | 53 | Cohort 2016 | 4892 | 4475 | 91 | 4929 | 4421 | 90 |

| | | | | | | | | | | | | | |
|---|---|---|---|---|---|---|---|---|---|---|---|---|---|
| Cohort 2017 | 3683 | 2301 | 62 | 4447 | 2433 | 55 | Cohort 2017 | 4988 | 4626 | 93 | 5036 | 4549 | 90 |
| Cohort 2018 | 3299 | 2148 | 65 | 4024 | 2305 | 57 | Cohort 2018 | 4977 | 4652 | 93 | 5012 | 4620 | 92 |
| Cohort 2019 | 3250 | 2103 | 65 | 3851 | 2305 | 60 | Cohort 2019 | 5195 | 4816 | 93 | 5220 | 4800 | 92 |
| Cohort 2020 | 2145 | 1221 | 57 | 2721 | 1441 | 53 | Cohort 2020 | 5147 | 4640 | 90 | 5164 | 4683 | 91 |
| Income source mother: Job in employment | | | | | | | Country of origin: Other, Asia | | | | | | |
| Cohort 2008 | 126454 | 123593 | 98 | 126369 | 123045 | 97 | Cohort 2008 | 8308 | 7349 | 88 | 8330 | 7046 | 85 |
| Cohort 2009 | 126154 | 123321 | 98 | 126070 | 122821 | 97 | Cohort 2009 | 8386 | 7508 | 90 | 8448 | 7215 | 85 |
| Cohort 2010 | 123131 | 120277 | 98 | 123051 | 119800 | 97 | Cohort 2010 | 8815 | 8014 | 91 | 8888 | 7753 | 87 |
| Cohort 2011 | 118906 | 116056 | 98 | 118860 | 115783 | 97 | Cohort 2011 | 8914 | 8142 | 91 | 9005 | 7909 | 88 |
| Cohort 2012 | 115778 | 112628 | 97 | 115696 | 112363 | 97 | Cohort 2012 | 9304 | 8479 | 91 | 9374 | 8319 | 89 |
| Cohort 2013 | 113800 | 110078 | 97 | 113734 | 110342 | 97 | Cohort 2013 | 8984 | 8240 | 92 | 9039 | 8141 | 90 |
| Cohort 2014 | 118000 | 113223 | 96 | 117904 | 113830 | 97 | Cohort 2014 | 9606 | 8732 | 91 | 9635 | 8645 | 90 |
| Cohort 2015 | 116902 | 110997 | 95 | 116815 | 111859 | 96 | Cohort 2015 | 10116 | 9039 | 89 | 10106 | 9008 | 89 |
| Cohort 2016 | 119594 | 113604 | 95 | 119476 | 113932 | 95 | Cohort 2016 | 11197 | 10266 | 92 | 11191 | 10191 | 91 |
| Cohort 2017 | 117271 | 112132 | 96 | 117171 | 112008 | 96 | Cohort 2017 | 11126 | 10444 | 94 | 11196 | 10403 | 93 |
| Cohort 2018 | 116973 | 112118 | 96 | 116832 | 111932 | 96 | Cohort 2018 | 11724 | 11062 | 94 | 11763 | 11044 | 94 |
| Cohort 2019 | 119930 | 114012 | 95 | 119855 | 114016 | 95 | Cohort 2019 | 11893 | 11207 | 94 | 11943 | 11213 | 94 |
| Cohort 2020 | 121259 | 112354 | 93 | 121142 | 113663 | 94 | Cohort 2020 | 11690 | 10705 | 92 | 11716 | 10842 | 93 |
| Income source mother: self-employed | | | | | | | Level of urbanisation: Not urbanised | | | | | | |
| Cohort 2008 | 13017 | 12196 | 94 | 12997 | 12202 | 94 | Cohort 2008 | 30407 | 28842 | 95 | 30380 | 28741 | 95 |
| Cohort 2009 | 13026 | 12134 | 93 | 13014 | 12126 | 93 | Cohort 2009 | 29830 | 28262 | 95 | 29809 | 28184 | 95 |
| Cohort 2010 | 13209 | 12289 | 93 | 13198 | 12289 | 93 | Cohort 2010 | 28743 | 27173 | 95 | 28717 | 27038 | 94 |
| Cohort 2011 | 13170 | 12257 | 93 | 13161 | 12250 | 93 | Cohort 2011 | 27401 | 25862 | 94 | 27385 | 25765 | 94 |
| Cohort 2012 | 13485 | 12462 | 92 | 13470 | 12434 | 92 | Cohort 2012 | 26170 | 24584 | 94 | 26157 | 24505 | 94 |
| Cohort 2013 | 13553 | 12389 | 91 | 13541 | 12430 | 92 | Cohort 2013 | 24684 | 23067 | 93 | 24672 | 23144 | 94 |
| Cohort 2014 | 14195 | 12769 | 90 | 14180 | 12880 | 91 | Cohort 2014 | 25185 | 23260 | 92 | 25166 | 23437 | 93 |

| | | | | | | | | | | | | | |
|---|---|---|---|---|---|---|---|---|---|---|---|---|---|
| Cohort 2015 | 14146 | 12555 | 89 | 14125 | 12695 | 90 | Cohort 2015 | 25053 | 22900 | 91 | 25028 | 23138 | 92 |
| Cohort 2016 | 14234 | 12583 | 88 | 14227 | 12642 | 89 | Cohort 2016 | 25256 | 23109 | 91 | 25235 | 23178 | 92 |
| Cohort 2017 | 13898 | 12553 | 90 | 13895 | 12515 | 90 | Cohort 2017 | 25079 | 23189 | 92 | 25070 | 23161 | 92 |
| Cohort 2018 | 14209 | 12776 | 90 | 14198 | 12742 | 90 | Cohort 2018 | 25503 | 23526 | 92 | 25487 | 23498 | 92 |
| Cohort 2019 | 14451 | 12807 | 89 | 14447 | 12835 | 89 | Cohort 2019 | 25696 | 23661 | 92 | 25691 | 23615 | 92 |
| Cohort 2020 | 13657 | 11567 | 85 | 13638 | 11739 | 86 | Cohort 2020 | 25935 | 23283 | 90 | 26043 | 23626 | 91 |
| Income source mother: Recipient benefit | | | | | | | Level of urbanisation: hardly | | | | | | |
| Cohort 2008 | 19690 | 18856 | 96 | 19658 | 18638 | 95 | Cohort 2008 | 35156 | 33954 | 97 | 35124 | 33772 | 96 |
| Cohort 2009 | 20548 | 19798 | 96 | 20529 | 19555 | 95 | Cohort 2009 | 33737 | 32645 | 97 | 33709 | 32541 | 97 |
| Cohort 2010 | 22681 | 22025 | 97 | 22645 | 21775 | 96 | Cohort 2010 | 32816 | 31682 | 97 | 32784 | 31570 | 96 |
| Cohort 2011 | 23139 | 22405 | 97 | 23109 | 22209 | 96 | Cohort 2011 | 32141 | 31009 | 96 | 32122 | 30927 | 96 |
| Cohort 2012 | 22606 | 21828 | 97 | 22596 | 21716 | 96 | Cohort 2012 | 31172 | 29899 | 96 | 31155 | 29847 | 96 |
| Cohort 2013 | 21541 | 20588 | 96 | 21476 | 20571 | 96 | Cohort 2013 | 28701 | 27229 | 95 | 28675 | 27270 | 95 |
| Cohort 2014 | 21549 | 20074 | 93 | 21428 | 20226 | 94 | Cohort 2014 | 30067 | 28274 | 94 | 30039 | 28437 | 95 |
| Cohort 2015 | 20182 | 18221 | 90 | 20012 | 18418 | 92 | Cohort 2015 | 29083 | 26919 | 93 | 29040 | 27195 | 94 |
| Cohort 2016 | 20391 | 18445 | 90 | 20277 | 18564 | 92 | Cohort 2016 | 29354 | 27287 | 93 | 29316 | 27382 | 93 |
| Cohort 2017 | 21840 | 19957 | 91 | 21768 | 19884 | 91 | Cohort 2017 | 29048 | 27290 | 94 | 29010 | 27237 | 94 |
| Cohort 2018 | 21259 | 19380 | 91 | 21196 | 19433 | 92 | Cohort 2018 | 28487 | 26938 | 95 | 28462 | 26900 | 95 |
| Cohort 2019 | 19270 | 16893 | 88 | 19245 | 17139 | 89 | Cohort 2019 | 28388 | 26730 | 94 | 28374 | 26720 | 94 |
| Cohort 2020 | 18512 | 15132 | 82 | 18462 | 15580 | 84 | Cohort 2020 | 28431 | 26032 | 92 | 28475 | 26384 | 93 |
| Income source mother: Pensioner | | | | | | | Level of urbanisation: moderately | | | | | | |
| Cohort 2008 | 1415 | 1334 | 94 | 1413 | 1334 | 94 | Cohort 2008 | 37879 | 36662 | 97 | 37859 | 36512 | 96 |
| Cohort 2009 | 1590 | 1505 | 95 | 1586 | 1511 | 95 | Cohort 2009 | 38302 | 37100 | 97 | 38265 | 36884 | 96 |
| Cohort 2010 | 1370 | 1304 | 95 | 1369 | 1302 | 95 | Cohort 2010 | 37074 | 35978 | 97 | 37041 | 35752 | 97 |
| Cohort 2011 | 1038 | 985 | 95 | 1037 | 978 | 94 | Cohort 2011 | 35596 | 34490 | 97 | 35577 | 34401 | 97 |
| Cohort 2012 | 780 | 738 | 95 | 779 | 730 | 94 | Cohort 2012 | 34224 | 33076 | 97 | 34180 | 32933 | 96 |

| | | | | | | | | | | | | | |
|---|---|---|---|---|---|---|---|---|---|---|---|---|---|
| Cohort 2013 | 897 | 827 | 92 | 897 | 841 | 94 | Cohort 2013 | 32598 | 31297 | 96 | 32554 | 31358 | 96 |
| Cohort 2014 | 846 | 777 | 92 | 844 | 782 | 93 | Cohort 2014 | 33089 | 31443 | 95 | 33038 | 31643 | 96 |
| Cohort 2015 | 604 | 536 | 89 | 601 | 544 | 91 | Cohort 2015 | 32237 | 30252 | 94 | 32174 | 30429 | 95 |
| Cohort 2016 | 533 | 481 | 90 | 531 | 478 | 90 | Cohort 2016 | 32520 | 30608 | 94 | 32477 | 30698 | 95 |
| Cohort 2017 | 473 | 428 | 90 | 470 | 425 | 90 | Cohort 2017 | 31883 | 30169 | 95 | 31848 | 30099 | 95 |
| Cohort 2018 | 456 | 417 | 91 | 456 | 416 | 91 | Cohort 2018 | 31504 | 29796 | 95 | 31444 | 29763 | 95 |
| Cohort 2019 | 306 | 277 | 91 | 306 | 278 | 91 | Cohort 2019 | 31718 | 29740 | 94 | 31690 | 29728 | 94 |
| Cohort 2020 | 267 | 218 | 82 | 267 | 222 | 83 | Cohort 2020 | 31940 | 29132 | 91 | 31960 | 29559 | 92 |
| Income source mother: Student | | | | | | | Level of urbanisation: strongly | | | | | | |
| Cohort 2008 | 3125 | 2990 | 96 | 3116 | 2964 | 95 | Cohort 2008 | 45514 | 44160 | 97 | 45452 | 43932 | 97 |
| Cohort 2009 | 3337 | 3200 | 96 | 3328 | 3146 | 95 | Cohort 2009 | 45542 | 44147 | 97 | 45486 | 43851 | 96 |
| Cohort 2010 | 3222 | 3109 | 96 | 3211 | 3072 | 96 | Cohort 2010 | 46811 | 45433 | 97 | 46754 | 45143 | 97 |
| Cohort 2011 | 3228 | 3119 | 97 | 3220 | 3091 | 96 | Cohort 2011 | 44261 | 42967 | 97 | 44212 | 42779 | 97 |
| Cohort 2012 | 3046 | 2926 | 96 | 3040 | 2895 | 95 | Cohort 2012 | 44469 | 43021 | 97 | 44429 | 42861 | 96 |
| Cohort 2013 | 2808 | 2690 | 96 | 2802 | 2701 | 96 | Cohort 2013 | 43881 | 42094 | 96 | 43826 | 42195 | 96 |
| Cohort 2014 | 2619 | 2446 | 93 | 2610 | 2471 | 95 | Cohort 2014 | 45047 | 42750 | 95 | 44962 | 42956 | 96 |
| Cohort 2015 | 2352 | 2111 | 90 | 2342 | 2147 | 92 | Cohort 2015 | 44263 | 41417 | 94 | 44158 | 41721 | 94 |
| Cohort 2016 | 2328 | 2087 | 90 | 2320 | 2110 | 91 | Cohort 2016 | 44851 | 41931 | 93 | 44747 | 42040 | 94 |
| Cohort 2017 | 2361 | 2177 | 92 | 2353 | 2185 | 93 | Cohort 2017 | 44638 | 42143 | 94 | 44557 | 42042 | 94 |
| Cohort 2018 | 2365 | 2182 | 92 | 2358 | 2175 | 92 | Cohort 2018 | 43655 | 41331 | 95 | 43560 | 41244 | 95 |
| Cohort 2019 | 2187 | 1958 | 90 | 2185 | 1980 | 91 | Cohort 2019 | 43668 | 40646 | 93 | 43620 | 40773 | 93 |
| Cohort 2020 | 2452 | 2058 | 84 | 2437 | 2118 | 87 | Cohort 2020 | 42402 | 38288 | 90 | 42430 | 38855 | 92 |
| Income source mother: Other | | | | | | | Level of urbanisation: Extremely | | | | | | |
| Cohort 2008 | 19919 | 18048 | 91 | 19857 | 17742 | 89 | Cohort 2008 | 35217 | 33860 | 96 | 35137 | 33365 | 95 |
| Cohort 2009 | 18845 | 17077 | 91 | 18794 | 16771 | 89 | Cohort 2009 | 36689 | 35369 | 96 | 36632 | 34899 | 95 |
| Cohort 2010 | 19074 | 17316 | 91 | 19017 | 17030 | 90 | Cohort 2010 | 37904 | 36619 | 97 | 37836 | 36279 | 96 |
| Cohort 2011 | 18545 | 16928 | 91 | 18489 | 16738 | 91 | Cohort 2011 | 39261 | 37956 | 97 | 39192 | 37661 | 96 |

| | | | | | | | | | | | | | |
|---|---|---|---|---|---|---|---|---|---|---|---|---|---|
| Cohort 2012 | 18261 | 16592 | 91 | 18197 | 16344 | 90 | Cohort 2012 | 38490 | 37083 | 96 | 38412 | 36789 | 96 |
| Cohort 2013 | 16905 | 15217 | 90 | 16848 | 15120 | 90 | Cohort 2013 | 40222 | 38618 | 96 | 40142 | 38505 | 96 |
| Cohort 2014 | 16239 | 14331 | 88 | 16199 | 14307 | 88 | Cohort 2014 | 40615 | 38378 | 94 | 40506 | 38470 | 95 |
| Cohort 2015 | 15036 | 13044 | 87 | 14972 | 13024 | 87 | Cohort 2015 | 39041 | 36325 | 93 | 38904 | 36535 | 94 |
| Cohort 2016 | 14089 | 12174 | 86 | 14052 | 12085 | 86 | Cohort 2016 | 39588 | 36742 | 93 | 39504 | 36792 | 93 |
| Cohort 2017 | 12925 | 11156 | 86 | 12864 | 10981 | 85 | Cohort 2017 | 38442 | 35864 | 93 | 38333 | 35687 | 93 |
| Cohort 2018 | 12311 | 10540 | 86 | 12239 | 10445 | 85 | Cohort 2018 | 38773 | 36102 | 93 | 38662 | 35978 | 93 |
| Cohort 2019 | 11882 | 9963 | 84 | 11861 | 9975 | 84 | Cohort 2019 | 38869 | 35368 | 91 | 38843 | 35617 | 92 |
| Cohort 2020 | 11745 | 9322 | 79 | 11669 | 9460 | 81 | Cohort 2020 | 38102 | 33116 | 87 | 38062 | 33624 | 88 |
| Income source mother: unknown | | | | | | | Level of urbanisation: Unknown | | | | | | |
| Cohort 2008 | 7235 | 4051 | 56 | 7986 | 3421 | 43 | Cohort 2008 | 6682 | 3590 | 54 | 7444 | 3024 | 41 |
| Cohort 2009 | 6944 | 4135 | 60 | 7771 | 3535 | 45 | Cohort 2009 | 6344 | 3647 | 57 | 7191 | 3106 | 43 |
| Cohort 2010 | 7005 | 4205 | 60 | 7802 | 3760 | 48 | Cohort 2010 | 6344 | 3640 | 57 | 7161 | 3246 | 45 |
| Cohort 2011 | 6732 | 4183 | 62 | 7553 | 3751 | 50 | Cohort 2011 | 6098 | 3649 | 60 | 6941 | 3267 | 47 |
| Cohort 2012 | 6529 | 4000 | 61 | 7413 | 3788 | 51 | Cohort 2012 | 5960 | 3511 | 59 | 6858 | 3335 | 49 |
| Cohort 2013 | 5906 | 3703 | 63 | 6735 | 3511 | 52 | Cohort 2013 | 5324 | 3187 | 60 | 6164 | 3044 | 49 |
| Cohort 2014 | 5454 | 3414 | 63 | 6344 | 3299 | 52 | Cohort 2014 | 4899 | 2929 | 60 | 5798 | 2852 | 49 |
| Cohort 2015 | 4965 | 3021 | 61 | 5818 | 3049 | 52 | Cohort 2015 | 4510 | 2672 | 59 | 5381 | 2718 | 51 |
| Cohort 2016 | 4277 | 2580 | 60 | 5138 | 2702 | 53 | Cohort 2016 | 3877 | 2277 | 59 | 4742 | 2423 | 51 |
| Cohort 2017 | 3616 | 2219 | 61 | 4407 | 2365 | 54 | Cohort 2017 | 3294 | 1967 | 60 | 4110 | 2137 | 52 |
| Cohort 2018 | 3185 | 2031 | 64 | 3957 | 2212 | 56 | Cohort 2018 | 2836 | 1751 | 62 | 3621 | 1972 | 54 |
| Cohort 2019 | 2993 | 1907 | 64 | 3636 | 2142 | 59 | Cohort 2019 | 2680 | 1672 | 62 | 3317 | 1912 | 58 |
| Cohort 2020 | 1606 | 860 | 54 | 2236 | 1068 | 48 | Cohort 2020 | 2688 | 1660 | 62 | 2881 | 1802 | 63 |
| Family size: 1-3 children | | | | | | | Generation: Dutch origin | | | | | | |
| Cohort 2008 | 172545 | 167376 | 97 | 172331 | 166275 | 96 | Cohort 2008 | 118975 | 114700 | 96 | 118970 | 114615 | 96 |
| Cohort 2009 | 172826 | 167650 | 97 | 172634 | 166559 | 96 | Cohort 2009 | 118260 | 114019 | 96 | 118265 | 113891 | 96 |
| Cohort 2010 | 172303 | 167205 | 97 | 172089 | 166132 | 97 | Cohort 2010 | 116353 | 112059 | 96 | 116368 | 111830 | 96 |
| Cohort 2011 | 167861 | 162776 | 97 | 167683 | 162026 | 97 | Cohort 2011 | 112850 | 108601 | 96 | 112850 | 108502 | 96 |
| Cohort 2012 | 164117 | 158579 | 97 | 163922 | 157936 | 96 | Cohort 2012 | 108811 | 104285 | 96 | 108810 | 104224 | 96 |

| | | | | | | | | | | | | | |
|---|---|---|---|---|---|---|---|---|---|---|---|---|---|
| Cohort 2013 | 159857 | 153400 | 96 | 159633 | 153537 | 96 | Cohort 2013 | 105450 | 100399 | 95 | 105451 | 100870 | 96 |
| Cohort 2014 | 163176 | 154938 | 95 | 162898 | 155677 | 96 | Cohort 2014 | 106602 | 100614 | 94 | 106619 | 101410 | 95 |
| Cohort 2015 | 159037 | 148994 | 94 | 158693 | 150077 | 95 | Cohort 2015 | 102457 | 95762 | 93 | 102447 | 96681 | 94 |
| Cohort 2016 | 160491 | 150498 | 94 | 160217 | 150847 | 94 | Cohort 2016 | 102144 | 95820 | 94 | 102140 | 96173 | 94 |
| Cohort 2017 | 158065 | 149396 | 95 | 157800 | 148990 | 94 | Cohort 2017 | 100069 | 94617 | 95 | 100064 | 94539 | 94 |
| Cohort 2018 | 156588 | 148292 | 95 | 156281 | 147957 | 95 | Cohort 2018 | 97802 | 92814 | 95 | 97785 | 92759 | 95 |
| Cohort 2019 | 157085 | 147094 | 94 | 156945 | 147294 | 94 | Cohort 2019 | 97499 | 92343 | 95 | 97476 | 92209 | 95 |
| Cohort 2020 | 155815 | 141583 | 91 | 155870 | 143489 | 92 | Cohort 2020 | 97023 | 89821 | 93 | 96991 | 90831 | 94 |
| Family size: 4 or more children | | | | | | | Generation: Migrant or child of first generation migrant(s) | | | | | | |
| Cohort 2008 | 11358 | 9893 | 87 | 11350 | 9853 | 87 | Cohort 2008 | 51981 | 47029 | 90 | 52448 | 45470 | 87 |
| Cohort 2009 | 11032 | 9655 | 88 | 11026 | 9594 | 87 | Cohort 2009 | 51511 | 47036 | 91 | 52052 | 45526 | 87 |
| Cohort 2010 | 10722 | 9397 | 88 | 10713 | 9359 | 87 | Cohort 2010 | 52194 | 47890 | 92 | 52684 | 46676 | 89 |
| Cohort 2011 | 10492 | 9249 | 88 | 10491 | 9246 | 88 | Cohort 2011 | 51078 | 47107 | 92 | 51667 | 46090 | 89 |
| Cohort 2012 | 10109 | 8820 | 87 | 10106 | 8744 | 87 | Cohort 2012 | 50467 | 46355 | 92 | 51082 | 45506 | 89 |
| Cohort 2013 | 9949 | 8662 | 87 | 9941 | 8680 | 87 | Cohort 2013 | 48420 | 44413 | 92 | 48963 | 43835 | 90 |
| Cohort 2014 | 10496 | 8902 | 85 | 10469 | 8992 | 86 | Cohort 2014 | 49846 | 45238 | 91 | 50338 | 44933 | 89 |
| Cohort 2015 | 10356 | 8599 | 83 | 10318 | 8706 | 84 | Cohort 2015 | 48906 | 43659 | 89 | 49296 | 43634 | 89 |
| Cohort 2016 | 10776 | 8960 | 83 | 10739 | 9003 | 84 | Cohort 2016 | 49557 | 44413 | 90 | 50030 | 44389 | 89 |
| Cohort 2017 | 10708 | 9025 | 84 | 10683 | 8982 | 84 | Cohort 2017 | 48639 | 44122 | 91 | 49069 | 43869 | 89 |
| Cohort 2018 | 10975 | 9128 | 83 | 10961 | 9147 | 83 | Cohort 2018 | 48802 | 44463 | 91 | 49168 | 44279 | 90 |
| Cohort 2019 | 10880 | 8810 | 81 | 10872 | 8896 | 82 | Cohort 2019 | 48973 | 43782 | 89 | 49387 | 44108 | 89 |
| Cohort 2020 | 10736 | 8108 | 76 | 10743 | 8315 | 77 | Cohort 2020 | 47729 | 40998 | 86 | 47969 | 41723 | 87 |
| Family size: Institutional | | | | | | | Generation: Child of second generation migrant(s) | | | | | | |
| Cohort 2008 | 265 | 202 | 76 | 261 | 182 | 70 | Cohort 2008 | 19899 | 19339 | 97 | 19908 | 19256 | 97 |
| Cohort 2009 | 241 | 218 | 90 | 238 | 204 | 86 | Cohort 2009 | 20673 | 20115 | 97 | 20690 | 20039 | 97 |
| Cohort 2010 | 330 | 288 | 87 | 330 | 288 | 87 | Cohort 2010 | 21145 | 20576 | 97 | 21164 | 20518 | 97 |

| | | | | | | | | | | | | | |
|---|---|---|---|---|---|---|---|---|---|---|---|---|---|
| Cohort 2011 | 317 | 269 | 85 | 317 | 264 | 83 | Cohort 2011 | 20830 | 20225 | 97 | 20855 | 20200 | 97 |
| Cohort 2012 | 312 | 275 | 88 | 311 | 261 | 84 | Cohort 2012 | 21207 | 20534 | 97 | 21226 | 20533 | 97 |
| Cohort 2013 | 292 | 255 | 87 | 297 | 257 | 87 | Cohort 2013 | 21540 | 20680 | 96 | 21549 | 20804 | 97 |
| Cohort 2014 | 347 | 280 | 81 | 347 | 277 | 80 | Cohort 2014 | 22454 | 21182 | 94 | 22482 | 21446 | 95 |
| Cohort 2015 | 298 | 231 | 78 | 295 | 236 | 80 | Cohort 2015 | 22824 | 21064 | 92 | 22847 | 21407 | 94 |
| Cohort 2016 | 334 | 250 | 75 | 340 | 256 | 75 | Cohort 2016 | 23745 | 21721 | 91 | 23774 | 21942 | 92 |
| Cohort 2017 | 337 | 253 | 75 | 342 | 260 | 76 | Cohort 2017 | 23676 | 21883 | 92 | 23710 | 21945 | 93 |
| Cohort 2018 | 383 | 291 | 76 | 383 | 286 | 75 | Cohort 2018 | 24154 | 22167 | 92 | 24200 | 22308 | 92 |
| Cohort 2019 | 425 | 287 | 68 | 424 | 284 | 67 | Cohort 2019 | 24547 | 21692 | 88 | 24580 | 22036 | 90 |
| Cohort 2020 | 378 | 263 | 70 | 376 | 257 | 68 | Cohort 2020 | 24746 | 20692 | 84 | 24809 | 21277 | 86 |
| Family size: unknown | | | | | | | Day care attendance: No | | | | | | |
| Cohort 2008 | 6687 | 3597 | 54 | 7454 | 3036 | 41 | Cohort 2008 | 88086 | 80259 | 91 | 88723 | 79035 | 89 |
| Cohort 2009 | 6345 | 3647 | 57 | 7194 | 3108 | 43 | Cohort 2009 | 85088 | 77790 | 91 | 85826 | 76598 | 89 |
| Cohort 2010 | 6337 | 3635 | 57 | 7161 | 3249 | 45 | Cohort 2010 | 86964 | 79657 | 92 | 87645 | 78557 | 90 |
| Cohort 2011 | 6088 | 3639 | 60 | 6938 | 3264 | 47 | Cohort 2011 | 90229 | 83143 | 92 | 90960 | 82231 | 90 |
| Cohort 2012 | 5947 | 3500 | 59 | 6852 | 3329 | 49 | Cohort 2012 | 88910 | 81565 | 92 | 89693 | 80855 | 90 |
| Cohort 2013 | 5312 | 3175 | 60 | 6162 | 3042 | 49 | Cohort 2013 | 84040 | 76559 | 91 | 84742 | 76419 | 90 |
| Cohort 2014 | 4883 | 2914 | 60 | 5795 | 2849 | 49 | Cohort 2014 | 77555 | 69284 | 89 | 78325 | 69565 | 89 |
| Cohort 2015 | 4496 | 2661 | 59 | 5379 | 2717 | 51 | Cohort 2015 | 68283 | 59536 | 87 | 68965 | 60024 | 87 |
| Cohort 2016 | 3845 | 2246 | 58 | 4725 | 2407 | 51 | Cohort 2016 | 60695 | 52341 | 86 | 61433 | 52632 | 86 |
| Cohort 2017 | 3274 | 1948 | 59 | 4103 | 2131 | 52 | Cohort 2017 | 54559 | 47194 | 87 | 55237 | 47104 | 85 |
| Cohort 2018 | 2812 | 1733 | 62 | 3611 | 1965 | 54 | Cohort 2018 | 53172 | 45831 | 86 | 53798 | 46007 | 86 |
| Cohort 2019 | 2629 | 1626 | 62 | 3294 | 1891 | 57 | Cohort 2019 | 50469 | 42142 | 84 | 51076 | 42741 | 84 |
| Cohort 2020 | 2569 | 1557 | 61 | 2862 | 1789 | 63 | Cohort 2020 | 45981 | 35993 | 78 | 46490 | 37171 | 80 |
| Day care attendance: yes | | | | | | | | | | | | | |
| Cohort 2008 | 102713 | 100753 | 98 | 102617 | 100258 | 98 | | | | | | | |
| Cohort 2009 | 105331 | 103355 | 98 | 105241 | 102842 | 98 | | | | | | | |
| Cohort 2010 | 102713 | 100853 | 98 | 102632 | 100456 | 98 | | | | | | | |
| Cohort 2011 | 94513 | 92776 | 98 | 94453 | 92555 | 98 | | | | | | | |
| Cohort 2012 | 91568 | 89602 | 98 | 91491 | 89408 | 98 | | | | | | | |

| | | | | | | |
|---|---|---|---|---|---|---|
| Cohort 2013 | 91365 | 88928 | 97 | 91286 | 89092 | 98 |
| Cohort 2014 | 101346 | 97749 | 96 | 101183 | 98229 | 97 |
| Cohort 2015 | 105899 | 100944 | 95 | 105715 | 101707 | 96 |
| Cohort 2016 | 114749 | 109611 | 96 | 114586 | 109879 | 96 |
| Cohort 2017 | 117822 | 113425 | 96 | 117688 | 113256 | 96 |
| Cohort 2018 | 117585 | 113612 | 97 | 117437 | 113348 | 97 |
| Cohort 2019 | 120547 | 115672 | 96 | 120456 | 115621 | 96 |
| Cohort 2020 | 123517 | 115518 | 94 | 123353 | 116671 | 95 |

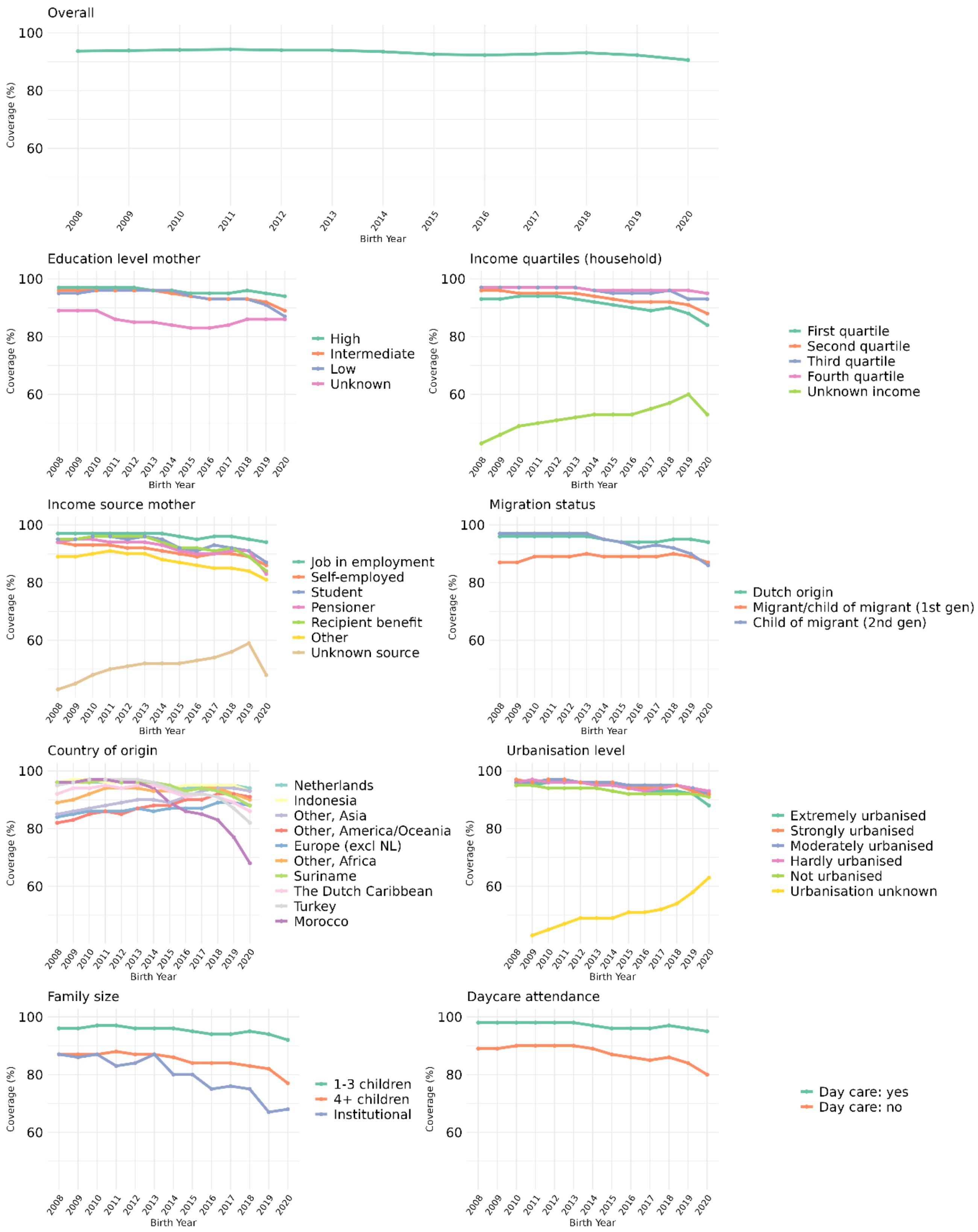


**Supplementary Figure S2. Crude DTaP-IPV vaccination coverage by sociodemographic determinant**

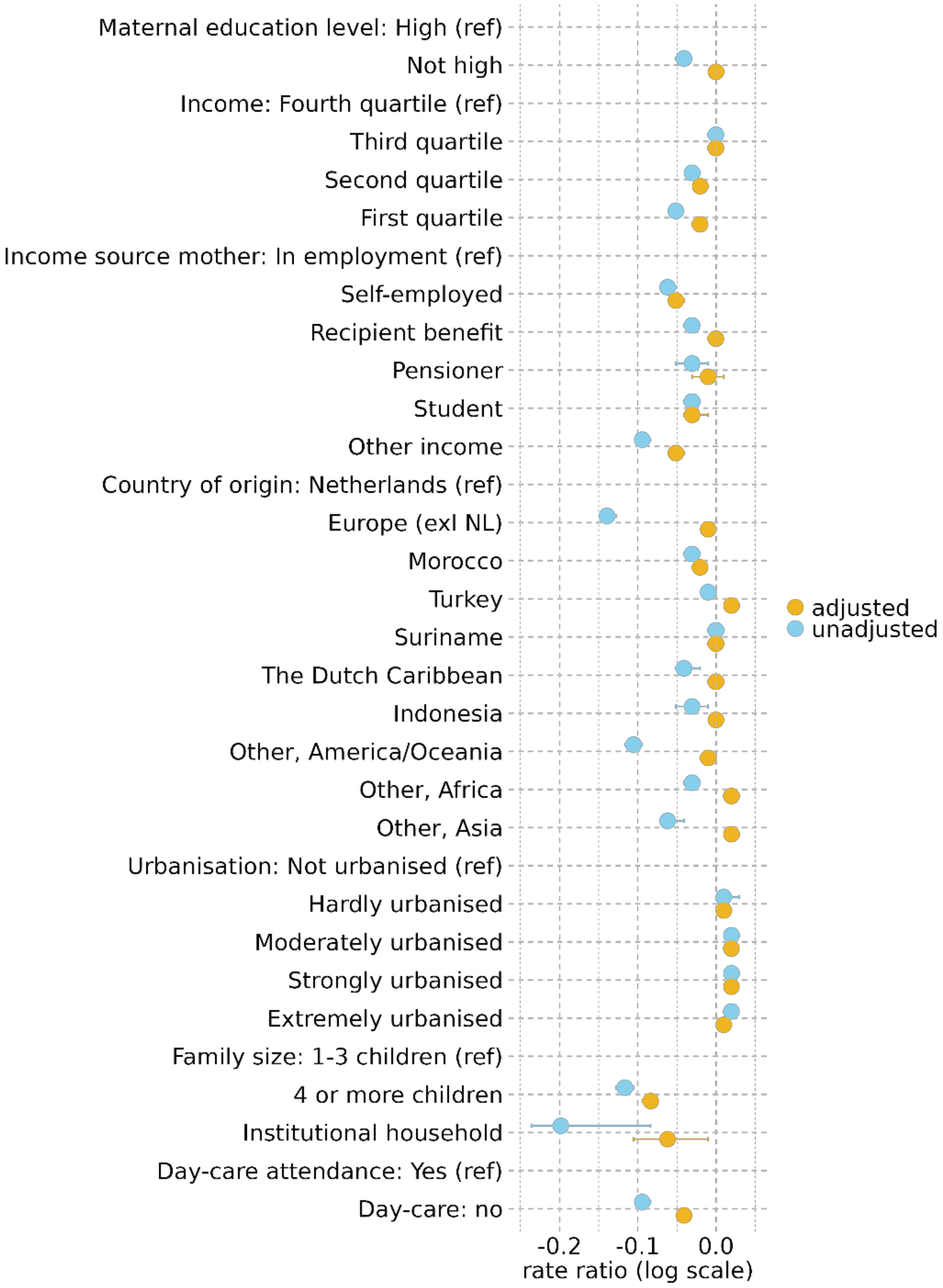


**Supplementary Figure S3. DTaP-IPV univariable and multivariable Poisson regression model – unadjusted and adjusted RRs**

In addition to all the variables listed, the RRs are also adjusted for birth cohort.

**Supplementary Table S4. Multivariable Poisson regression model with interaction terms for birth cohort * sociodemographic variable for both MMR and DTaP-IPV vaccination**

| | MMR vaccination model | Sensitivity analysis*: | | DTaP-IPV vaccination model | Sensitivity analysis*: | |
|---|---|---|---|---|---|---|
| Category | aRR (95% CI) | p-value | aRR (95% CI) | aRR (95% CI) | p-value | aRR (95% CI) |
| Birth_year2009 | 1.00 (0.97 - 1.02) | 0.807 | 1.00 (0.97 - 1.02) | 1.00 (0.98 - 1.02) | 0.980 | 1.00 (0.98 -1.02) |
| Birth_year2010 | 1.00 (0.97 - 1.02) | 0.767 | 1.00 (0.97 - 1.02) | 1.00 (0.97 - 1.02) | 0.742 | 1.00 (0.97 -1.02) |
| Birth_year2011 | 0.99 (0.97 - 1.02) | 0.542 | 0.99 (0.97 - 1.02) | 0.99 (0.97 - 1.02) | 0.598 | 0.99 (0.97 -1.02) |
| Birth_year2012 | 0.99 (0.97 - 1.01) | 0.374 | 0.99 (0.97 - 1.01) | 0.99 (0.97 - 1.01) | 0.483 | 0.99 (0.97 -1.01) |
| Birth_year2013 | 0.99 (0.96 - 1.01) | 0.256 | 0.99 (0.96 - 1.01) | 0.99 (0.97 - 1.02) | 0.517 | 0.99 (0.97 -1.01) |
| Birth_year2014 | 0.98 (0.96 - 1.00) | 0.074 | 0.98 (0.95 - 1.00) | 0.99 (0.97 - 1.01) | 0.293 | 0.99 (0.96 -1.01) |
| Birth_year2015 | 0.97 (0.95 - 1.00) | 0.027 | 0.97 (0.95 - 1.00) | 0.98 (0.96 - 1.01) | 0.186 | 0.98 (0.96 -1.01) |
| Birth_year2016 | 0.98 (0.96 - 1.00) | 0.083 | 0.98 (0.96 - 1.00) | 0.98 (0.96 - 1.01) | 0.141 | 0.98 (0.96 -1.00) |
| Birth_year2017 | 0.99 (0.96 - 1.01) | 0.237 | 0.99 (0.96 - 1.01) | 0.99 (0.96 - 1.01) | 0.272 | 0.98 (0.96 -1.01) |
| Birth_year2018 | 0.99 (0.96 - 1.01) | 0.228 | 0.98 (0.96 - 1.01) | 0.99 (0.96 - 1.01) | 0.219 | 0.98 (0.96 -1.01) |
| Birth_year2019 | 0.99 (0.97 - 1.02) | 0.598 | 0.99 (0.97 - 1.01) | 0.99 (0.97 - 1.01) | 0.324 | 0.99 (0.96 -1.01) |
| Birth_year2020 | 0.98 (0.96 - 1.00) | 0.094 | 0.98 (0.96 - 1.00) | 0.99 (0.96 - 1.01) | 0.253 | 0.98 (0.96 -1.01) |
| High_education_motherNo | 1.00 (0.99 - 1.02) | 0.475 | 1.00 (0.99 - 1.02) | 1.00 (0.99 - 1.01) | 0.977 | 1.00 (0.99 -1.01) |
| SDIhh_quartileFirst quartile | 0.99 (0.97 - 1.01) | 0.222 | 0.99 (0.97 - 1.01) | 0.99 (0.97 - 1.01) | 0.264 | 0.99 (0.97 -1.01) |
| SDIhh_quartileSecond quartile | 0.99 (0.98 - 1.01) | 0.335 | 0.99 (0.98 - 1.01) | 1.00 (0.98 - 1.01) | 0.645 | 1.00 (0.98 -1.01) |
| SDIhh_quartileThird quartile | 1.00 (0.98 - 1.01) | 0.525 | 1.00 (0.98 - 1.01) | 1.00 (0.99 - 1.01) | 0.996 | 1.00 (0.99 -1.01) |
| SDIhh_quartileUnknown | NA | NA | 0.87 (0.81 - 0.93) | NA | NA | 0.79 (0.72 -0.85) |
| Country_origin_thirdgenEurope (excl NL) | 1.00 (0.98 - 1.01) | 0.593 | 0.99 (0.97 - 1.00) | 0.99 (0.97 - 1.00) | 0.157 | 0.97 (0.95 -0.98) |
| Country_origin_thirdgenIndonesia | 1.00 (0.98 - 1.03) | 0.694 | 1.00 (0.98 - 1.03) | 1.00 (0.98 - 1.03) | 0.830 | 1.00 (0.98 -1.03) |
| Country_origin_thirdgenMorocco | 1.05 (1.02 - 1.07) | <0.001 | 1.05 (1.02 - 1.07) | 1.04 (1.02 - 1.07) | 0.001 | 1.04 (1.02 -1.07) |
| Country_origin_thirdgenOther, Africa | 1.00 (0.97 - 1.03) | 0.787 | 0.98 (0.95 - 1.01) | 0.98 (0.95 - 1.01) | 0.226 | 0.98 (0.95 -1.01) |
| Country_origin_thirdgenOther, America/Oceania | 0.99 (0.96 - 1.03) | 0.699 | 1.01 (0.98 - 1.04) | 0.98 (0.95 - 1.02) | 0.387 | 0.97 (0.93 -1.00) |
| Country_origin_thirdgenOther, Asia | 1.01 (0.99 - 1.04) | 0.308 | 1.00 (0.98 - 1.03) | 1.00 (0.97 - 1.02) | 0.781 | 0.99 (0.96 -1.01) |
| Country_origin_thirdgenSuriname | 1.01 (0.99 - 1.04) | 0.295 | 1.02 (0.99 - 1.04) | 1.00 (0.98 - 1.03) | 0.848 | 1.00 (0.98 -1.03) |
| Country_origin_thirdgenThe Dutch Caribbean | 1.00 (0.97 - 1.04) | 0.846 | 1.01 (0.97 - 1.04) | 0.99 (0.96 - 1.03) | 0.772 | 1.01 (0.97 -1.05) |
| Country_origin_thirdgenTurkey | 1.03 (1.01 - 1.06) | 0.011 | 1.03 (1.01 - 1.06) | 1.03 (1.00 - 1.05) | 0.057 | 1.03 (1.00 -1.05) |
| SES.mother5Self-employed | 0.97 (0.95 - 0.99) | 0.001 | 0.97 (0.95 - 0.99) | 0.97 (0.96 - 0.99) | 0.007 | 0.98 (0.96 -0.99) |
| SES.mother5Recipient benefit | 0.99 (0.97 - 1.01) | 0.229 | 0.99 (0.98 - 1.01) | 0.99 (0.97 - 1.01) | 0.178 | 0.99 (0.97 -1.01) |
| SES.mother5Pensioner | 0.98 (0.93 - 1.04) | 0.504 | 0.98 (0.93 - 1.04) | 0.99 (0.94 - 1.04) | 0.677 | 0.99 (0.94 -1.05) |
| SES.mother5Child/student | 0.98 (0.94 - 1.02) | 0.261 | 0.98 (0.94 - 1.02) | 0.98 (0.95 - 1.02) | 0.386 | 0.98 (0.95 -1.02) |

| | | | | | | |
|---|---|---|---|---|---|---|
| SES.mother5Other | 0.95 (0.93 - 0.97) | <0.001 | 0.95 (0.94 - 0.97) | 0.95 (0.93 - 0.96) | <0.001 | 0.95 (0.93 -0.97) |
| SES.mother5Unknown | NA | NA | 0.87 (0.79 - 0.94) | NA | NA | 0.81 (0.72 -0.89) |
| UrbanisationExtremely urbanised | 1.01 (0.99 - 1.02) | 0.469 | 1.01 (0.99 - 1.02) | 1.00 (0.99 - 1.02) | 0.806 | 1.00 (0.99 -1.02) |
| UrbanisationHardly urbanised | 1.01 (0.99 - 1.03) | 0.199 | 1.01 (0.99 - 1.03) | 1.01 (0.99 - 1.03) | 0.266 | 1.01 (0.99 -1.02) |
| UrbanisationModerately urbanised | 1.01 (1.00 - 1.03) | 0.151 | 1.01 (1.00 - 1.03) | 1.01 (1.00 - 1.03) | 0.125 | 1.01 (1.00 -1.03) |
| UrbanisationStrongly urbanised | 1.01 (1.00 - 1.03) | 0.068 | 1.01 (1.00 - 1.03) | 1.02 (1.00 - 1.03) | 0.040 | 1.02 (1.00 -1.03) |
| UrbanisationUnknown | NA | NA | 0.74 (0.32 - 1.16) | NA | NA | 0.61 (0.15 -1.07) |
| family_size4 or more children | 0.91 (0.89 - 0.93) | <0.001 | 0.91 (0.89 - 0.93) | 0.91 (0.90 - 0.93) | <0.001 | 0.91 (0.89 -0.94) |
| family_sizeInstitutional | 0.90 (0.75 - 1.06) | 0.206 | 0.86 (0.72 - 1.00) | 0.86 (0.72 - 1.03) | 0.097 | 0.83 (0.68 -0.97) |
| Family_sizeUnknown | NA | NA | 0.99 (0.58 - 1.41) | NA | NA | 1.10 (0.64 -1.55) |
| Daycare2No daycare | 0.98 (0.97 - 0.99) | <0.001 | 0.97 (0.96 - 0.98) | 0.98 (0.97 - 0.99) | <0.001 | 0.97 (0.96 -0.98) |
| Birth_year2009:High_education_motherNo | 1.00 (0.98 - 1.02) | 0.997 | 1.00 (0.98 - 1.02) | 1.00 (0.98 - 1.02) | 0.965 | 1.00 (0.98 -1.02) |
| Birth_year2010:High_education_motherNo | 1.00 (0.99 - 1.02) | 0.781 | 1.00 (0.99 - 1.02) | 1.00 (0.99 - 1.02) | 0.698 | 1.00 (0.99 -1.02) |
| Birth_year2011:High_education_motherNo | 1.00 (0.99 - 1.02) | 0.660 | 1.00 (0.99 - 1.02) | 1.00 (0.99 - 1.02) | 0.630 | 1.00 (0.99 -1.02) |
| Birth_year2012:High_education_motherNo | 1.00 (0.99 - 1.02) | 0.754 | 1.00 (0.99 - 1.02) | 1.00 (0.99 - 1.02) | 0.702 | 1.00 (0.99 -1.02) |
| Birth_year2013:High_education_motherNo | 1.01 (0.99 - 1.02) | 0.520 | 1.01 (0.99 - 1.02) | 1.01 (0.99 - 1.02) | 0.432 | 1.01 (0.99 -1.02) |
| Birth_year2014:High_education_motherNo | 1.01 (0.99 - 1.02) | 0.404 | 1.01 (0.99 - 1.02) | 1.01 (0.99 - 1.03) | 0.240 | 1.01 (0.99 -1.03) |
| Birth_year2015:High_education_motherNo | 1.01 (0.99 - 1.02) | 0.407 | 1.01 (0.99 - 1.02) | 1.01 (0.99 - 1.03) | 0.271 | 1.01 (0.99 -1.02) |
| Birth_year2016:High_education_motherNo | 1.00 (0.99 - 1.02) | 0.736 | 1.00 (0.99 - 1.02) | 1.01 (0.99 - 1.02) | 0.352 | 1.01 (0.99 -1.02) |
| Birth_year2017:High_education_motherNo | 1.00 (0.98 - 1.02) | 0.948 | 1.00 (0.98 - 1.02) | 1.00 (0.99 - 1.02) | 0.621 | 1.00 (0.99 -1.02) |
| Birth_year2018:High_education_motherNo | 1.00 (0.98 - 1.01) | 0.896 | 1.00 (0.98 - 1.01) | 1.01 (0.99 - 1.02) | 0.508 | 1.01 (0.99 -1.02) |
| Birth_year2019:High_education_motherNo | 1.00 (0.98 - 1.01) | 0.793 | 1.00 (0.98 - 1.01) | 1.00 (0.99 - 1.02) | 0.646 | 1.00 (0.99 -1.02) |
| Birth_year2020:High_education_motherNo | 0.99 (0.97 - 1.00) | 0.148 | 0.99 (0.97 - 1.01) | 1.00 (0.98 - 1.01) | 0.624 | 1.00 (0.98 -1.01) |
| Birth_year2009:SDIhh_quartileFirst quartile | 1.00 (0.97 - 1.02) | 0.791 | 1.00 (0.97 - 1.02) | 1.00 (0.97 - 1.02) | 0.701 | 1.00 (0.97 -1.02) |
| Birth_year2010:SDIhh_quartileFirst quartile | 1.00 (0.97 - 1.02) | 0.724 | 1.00 (0.97 - 1.02) | 1.00 (0.97 - 1.02) | 0.681 | 1.00 (0.97 -1.02) |
| Birth_year2011:SDIhh_quartileFirst quartile | 0.99 (0.97 - 1.02) | 0.622 | 0.99 (0.97 - 1.02) | 1.00 (0.97 - 1.02) | 0.758 | 1.00 (0.97 -1.02) |
| Birth_year2012:SDIhh_quartileFirst quartile | 1.00 (0.97 - 1.02) | 0.768 | 1.00 (0.97 - 1.02) | 1.00 (0.97 - 1.02) | 0.858 | 1.00 (0.97 -1.02) |
| Birth_year2013:SDIhh_quartileFirst quartile | 0.99 (0.97 - 1.02) | 0.490 | 0.99 (0.97 - 1.02) | 0.99 (0.97 - 1.02) | 0.497 | 0.99 (0.97 -1.02) |
| Birth_year2014:SDIhh_quartileFirst quartile | 0.99 (0.97 - 1.02) | 0.444 | 0.99 (0.97 - 1.01) | 0.99 (0.97 - 1.02) | 0.505 | 0.99 (0.97 -1.02) |
| Birth_year2015:SDIhh_quartileFirst quartile | 0.98 (0.96 - 1.01) | 0.156 | 0.98 (0.96 - 1.01) | 0.98 (0.96 - 1.01) | 0.212 | 0.99 (0.96 -1.01) |
| Birth_year2016:SDIhh_quartileFirst quartile | 0.97 (0.95 - 1.00) | 0.037 | 0.97 (0.95 - 1.00) | 0.98 (0.95 - 1.00) | 0.076 | 0.98 (0.95 -1.00) |
| Birth_year2017:SDIhh_quartileFirst quartile | 0.98 (0.95 - 1.00) | 0.072 | 0.98 (0.95 - 1.00) | 0.98 (0.95 - 1.00) | 0.053 | 0.98 (0.95 -1.00) |
| Birth_year2018:SDIhh_quartileFirst quartile | 0.98 (0.96 - 1.01) | 0.134 | 0.98 (0.96 - 1.01) | 0.98 (0.96 - 1.01) | 0.128 | 0.98 (0.96 -1.01) |
| Birth_year2019:SDIhh_quartileFirst quartile | 0.96 (0.94 - 0.99) | 0.007 | 0.97 (0.94 - 0.99) | 0.97 (0.95 - 1.00) | 0.027 | 0.97 (0.95 -1.00) |
| Birth_year2020:SDIhh_quartileFirst quartile | 0.96 (0.93 - 0.98) | 0.001 | 0.96 (0.93 - 0.98) | 0.96 (0.94 - 0.99) | 0.003 | 0.96 (0.93 -0.99) |
| Birth_year2009:SDIhh_quartileSecond quartile | 1.00 (0.98 - 1.02) | 0.962 | 1.00 (0.98 - 1.02) | 1.00 (0.98 - 1.02) | 0.946 | 1.00 (0.98 -1.02) |

| | | | | | | |
|---|---|---|---|---|---|---|
| Birth_year2010:SDIhh_quartileSecond quartile | 1.00 (0.98 - 1.01) | 0.643 | 1.00 (0.98 - 1.01) | 0.99 (0.97 - 1.01) | 0.500 | 0.99 (0.97 -1.01) |
| Birth_year2011:SDIhh_quartileSecond quartile | 1.00 (0.98 - 1.02) | 0.668 | 1.00 (0.98 - 1.02) | 0.99 (0.97 - 1.01) | 0.546 | 0.99 (0.97 -1.01) |
| Birth_year2012:SDIhh_quartileSecond quartile | 1.00 (0.98 - 1.02) | 0.652 | 1.00 (0.98 - 1.01) | 0.99 (0.97 - 1.01) | 0.505 | 0.99 (0.97 -1.01) |
| Birth_year2013:SDIhh_quartileSecond quartile | 0.99 (0.97 - 1.01) | 0.311 | 0.99 (0.97 - 1.01) | 0.99 (0.97 - 1.01) | 0.277 | 0.99 (0.97 -1.01) |
| Birth_year2014:SDIhh_quartileSecond quartile | 0.99 (0.97 - 1.01) | 0.184 | 0.99 (0.97 - 1.01) | 0.99 (0.97 - 1.01) | 0.185 | 0.99 (0.97 -1.01) |
| Birth_year2015:SDIhh_quartileSecond quartile | 0.98 (0.96 - 1.00) | 0.062 | 0.98 (0.96 - 1.00) | 0.98 (0.96 - 1.00) | 0.067 | 0.98 (0.96 -1.00) |
| Birth_year2016:SDIhh_quartileSecond quartile | 0.98 (0.96 - 1.00) | 0.016 | 0.98 (0.96 - 1.00) | 0.98 (0.96 - 1.00) | 0.014 | 0.98 (0.96 -1.00) |
| Birth_year2017:SDIhh_quartileSecond quartile | 0.98 (0.96 - 1.00) | 0.035 | 0.98 (0.96 - 1.00) | 0.98 (0.96 - 1.00) | 0.029 | 0.98 (0.96 -1.00) |
| Birth_year2018:SDIhh_quartileSecond quartile | 0.98 (0.96 - 1.00) | 0.036 | 0.98 (0.96 - 1.00) | 0.97 (0.95 - 0.99) | 0.011 | 0.97 (0.95 -0.99) |
| Birth_year2019:SDIhh_quartileSecond quartile | 0.97 (0.95 - 0.99) | 0.007 | 0.97 (0.95 - 0.99) | 0.97 (0.95 - 0.99) | 0.008 | 0.97 (0.95 -0.99) |
| Birth_year2020:SDIhh_quartileSecond quartile | 0.97 (0.95 - 0.99) | 0.001 | 0.97 (0.94 - 0.99) | 0.97 (0.95 - 0.99) | 0.002 | 0.97 (0.95 -0.99) |
| Birth_year2009:SDIhh_quartileThird quartile | 1.00 (0.99 - 1.02) | 0.692 | 1.00 (0.99 - 1.02) | 1.00 (0.98 - 1.02) | 0.911 | 1.00 (0.98 -1.02) |
| Birth_year2010:SDIhh_quartileThird quartile | 1.00 (0.98 - 1.02) | 0.921 | 1.00 (0.98 - 1.02) | 1.00 (0.98 - 1.02) | 0.886 | 1.00 (0.98 -1.02) |
| Birth_year2011:SDIhh_quartileThird quartile | 1.00 (0.98 - 1.02) | 0.784 | 1.00 (0.98 - 1.02) | 1.00 (0.98 - 1.02) | 0.974 | 1.00 (0.98 -1.02) |
| Birth_year2012:SDIhh_quartileThird quartile | 1.00 (0.98 - 1.02) | 0.840 | 1.00 (0.98 - 1.02) | 1.00 (0.98 - 1.02) | 0.877 | 1.00 (0.98 -1.02) |
| Birth_year2013:SDIhh_quartileThird quartile | 1.00 (0.98 - 1.02) | 1.000 | 1.00 (0.98 - 1.02) | 1.00 (0.98 - 1.02) | 0.827 | 1.00 (0.98 -1.02) |
| Birth_year2014:SDIhh_quartileThird quartile | 1.00 (0.98 - 1.02) | 0.908 | 1.00 (0.98 - 1.02) | 1.00 (0.98 - 1.01) | 0.686 | 1.00 (0.98 -1.02) |
| Birth_year2015:SDIhh_quartileThird quartile | 0.99 (0.97 - 1.01) | 0.460 | 0.99 (0.97 - 1.01) | 0.99 (0.97 - 1.01) | 0.353 | 0.99 (0.97 -1.01) |
| Birth_year2016:SDIhh_quartileThird quartile | 0.99 (0.97 - 1.01) | 0.440 | 0.99 (0.97 - 1.01) | 0.99 (0.97 - 1.01) | 0.290 | 0.99 (0.97 -1.01) |
| Birth_year2017:SDIhh_quartileThird quartile | 0.99 (0.98 - 1.01) | 0.548 | 0.99 (0.98 - 1.01) | 0.99 (0.97 - 1.01) | 0.352 | 0.99 (0.97 -1.01) |
| Birth_year2018:SDIhh_quartileThird quartile | 1.00 (0.98 - 1.02) | 0.721 | 1.00 (0.98 - 1.02) | 0.99 (0.98 - 1.01) | 0.503 | 0.99 (0.98 -1.01) |
| Birth_year2019:SDIhh_quartileThird quartile | 0.99 (0.98 - 1.01) | 0.541 | 0.99 (0.98 - 1.01) | 0.99 (0.97 - 1.01) | 0.451 | 0.99 (0.97 -1.01) |
| Birth_year2020:SDIhh_quartileThird quartile | 0.99 (0.98 - 1.01) | 0.577 | 0.99 (0.98 - 1.01) | 0.99 (0.97 - 1.01) | 0.415 | 0.99 (0.97 -1.01) |
| Birth_year2009:SDIhh_quartileUnknown | NA | NA | 1.03 (0.94 - 1.11) | NA | NA | 1.03 (0.94 -1.12) |
| Birth_year2010:SDIhh_quartileUnknown | NA | NA | 1.04 (0.95 - 1.12) | NA | NA | 1.04 (0.95 -1.13) |
| Birth_year2011:SDIhh_quartileUnknown | NA | NA | 1.03 (0.95 - 1.12) | NA | NA | 1.03 (0.94 -1.12) |
| Birth_year2012:SDIhh_quartileUnknown | NA | NA | 1.06 (0.97 - 1.14) | NA | NA | 1.06 (0.97 -1.15) |
| Birth_year2013:SDIhh_quartileUnknown | NA | NA | 1.02 (0.93 - 1.11) | NA | NA | 1.06 (0.96 -1.15) |
| Birth_year2014:SDIhh_quartileUnknown | NA | NA | 1.03 (0.94 - 1.12) | NA | NA | 1.07 (0.98 -1.17) |
| Birth_year2015:SDIhh_quartileUnknown | NA | NA | 1.03 (0.94 - 1.12) | NA | NA | 1.06 (0.96 -1.15) |
| Birth_year2016:SDIhh_quartileUnknown | NA | NA | 1.00 (0.91 - 1.10) | NA | NA | 1.03 (0.94 -1.13) |
| Birth_year2017:SDIhh_quartileUnknown | NA | NA | 1.02 (0.92 - 1.11) | NA | NA | 1.08 (0.98 -1.17) |
| Birth_year2018:SDIhh_quartileUnknown | NA | NA | 1.04 (0.94 - 1.13) | NA | NA | 1.09 (0.99 -1.19) |
| Birth_year2019:SDIhh_quartileUnknown | NA | NA | 1.02 (0.93 - 1.12) | NA | NA | 1.09 (0.99 -1.18) |
| Birth_year2020:SDIhh_quartileUnknown | NA | NA | 0.93 (0.84 - 1.03) | NA | NA | 1.03 (0.93 -1.12) |

| | | | | | | |
|---|---|---|---|---|---|---|
| Birth_year2009:Country_origin_thirdgenEurope (excl NL) | 1.00 (0.98 - 1.03) | 0.893 | 1.00 (0.98 - 1.03) | 1.00 (0.98 - 1.02) | 0.944 | 1.00 (0.98 -1.03) |
| Birth_year2010:Country_origin_thirdgenEurope (excl NL) | 1.00 (0.98 - 1.03) | 0.873 | 1.00 (0.98 - 1.02) | 1.00 (0.98 - 1.03) | 0.678 | 1.01 (0.98 -1.03) |
| Birth_year2011:Country_origin_thirdgenEurope (excl NL) | 1.00 (0.98 - 1.03) | 0.758 | 1.00 (0.98 - 1.02) | 1.00 (0.98 - 1.03) | 0.689 | 1.01 (0.98 -1.03) |
| Birth_year2012:Country_origin_thirdgenEurope (excl NL) | 1.00 (0.98 - 1.03) | 0.758 | 1.00 (0.98 - 1.03) | 1.00 (0.98 - 1.03) | 0.774 | 1.01 (0.98 -1.03) |
| Birth_year2013:Country_origin_thirdgenEurope (excl NL) | 1.00 (0.98 - 1.03) | 0.884 | 1.00 (0.98 - 1.02) | 1.00 (0.98 - 1.03) | 0.755 | 1.01 (0.98 -1.03) |
| Birth_year2014:Country_origin_thirdgenEurope (excl NL) | 1.00 (0.98 - 1.03) | 0.816 | 1.00 (0.98 - 1.02) | 1.00 (0.98 - 1.03) | 0.719 | 1.01 (0.98 -1.03) |
| Birth_year2015:Country_origin_thirdgenEurope (excl NL) | 1.00 (0.98 - 1.02) | 0.983 | 1.00 (0.98 - 1.02) | 1.01 (0.98 - 1.03) | 0.646 | 1.01 (0.99 -1.03) |
| Birth_year2016:Country_origin_thirdgenEurope (excl NL) | 1.00 (0.97 - 1.02) | 0.780 | 1.00 (0.97 - 1.02) | 1.00 (0.98 - 1.03) | 0.745 | 1.01 (0.99 -1.03) |
| Birth_year2017:Country_origin_thirdgenEurope (excl NL) | 1.00 (0.98 - 1.02) | 0.935 | 1.00 (0.98 - 1.02) | 1.00 (0.98 - 1.03) | 0.831 | 1.01 (0.99 -1.03) |
| Birth_year2018:Country_origin_thirdgenEurope (excl NL) | 1.00 (0.98 - 1.02) | 0.999 | 1.00 (0.98 - 1.03) | 1.00 (0.98 - 1.03) | 0.872 | 1.01 (0.99 -1.04) |
| Birth_year2019:Country_origin_thirdgenEurope (excl NL) | 1.00 (0.97 - 1.02) | 0.805 | 1.00 (0.98 - 1.02) | 1.00 (0.98 - 1.03) | 0.816 | 1.02 (0.99 -1.04) |
| Birth_year2020:Country_origin_thirdgenEurope (excl NL) | 0.99 (0.97 - 1.02) | 0.503 | 1.00 (0.98 - 1.02) | 1.00 (0.98 - 1.02) | 0.954 | 1.01 (0.99 -1.04) |
| Birth_year2009:Country_origin_thirdgenIndonesia | 1.00 (0.97 - 1.04) | 0.915 | 1.00 (0.97 - 1.04) | 1.01 (0.97 - 1.04) | 0.704 | 1.01 (0.97 -1.04) |
| Birth_year2010:Country_origin_thirdgenIndonesia | 1.00 (0.97 - 1.04) | 0.894 | 1.00 (0.97 - 1.04) | 1.01 (0.97 - 1.04) | 0.685 | 1.01 (0.97 -1.04) |
| Birth_year2011:Country_origin_thirdgenIndonesia | 1.00 (0.97 - 1.03) | 0.971 | 1.00 (0.97 - 1.03) | 1.00 (0.97 - 1.04) | 0.994 | 1.00 (0.97 -1.03) |
| Birth_year2012:Country_origin_thirdgenIndonesia | 1.00 (0.97 - 1.04) | 0.834 | 1.00 (0.97 - 1.04) | 1.01 (0.97 - 1.04) | 0.683 | 1.01 (0.97 -1.04) |
| Birth_year2013:Country_origin_thirdgenIndonesia | 1.00 (0.97 - 1.04) | 0.877 | 1.00 (0.97 - 1.04) | 1.01 (0.97 - 1.04) | 0.771 | 1.00 (0.97 -1.04) |
| Birth_year2014:Country_origin_thirdgenIndonesia | 1.00 (0.97 - 1.04) | 0.917 | 1.00 (0.97 - 1.04) | 1.01 (0.97 - 1.04) | 0.737 | 1.00 (0.97 -1.04) |
| Birth_year2015:Country_origin_thirdgenIndonesia | 1.00 (0.97 - 1.04) | 0.965 | 1.00 (0.96 - 1.03) | 1.00 (0.97 - 1.04) | 0.941 | 1.00 (0.96 -1.03) |
| Birth_year2016:Country_origin_thirdgenIndonesia | 1.00 (0.96 - 1.04) | 0.956 | 1.00 (0.96 - 1.04) | 1.00 (0.97 - 1.04) | 0.882 | 1.00 (0.97 -1.04) |
| Birth_year2017:Country_origin_thirdgenIndonesia | 1.00 (0.97 - 1.04) | 0.896 | 1.00 (0.97 - 1.04) | 1.01 (0.97 - 1.04) | 0.787 | 1.01 (0.97 -1.04) |
| Birth_year2018:Country_origin_thirdgenIndonesia | 1.00 (0.96 - 1.04) | 0.970 | 1.00 (0.96 - 1.04) | 1.00 (0.96 - 1.04) | 0.929 | 1.00 (0.97 -1.04) |
| Birth_year2019:Country_origin_thirdgenIndonesia | 0.99 (0.95 - 1.03) | 0.576 | 0.99 (0.95 - 1.03) | 1.00 (0.96 - 1.04) | 0.973 | 1.00 (0.96 -1.04) |
| Birth_year2020:Country_origin_thirdgenIndonesia | 0.99 (0.95 - 1.03) | 0.563 | 0.99 (0.95 - 1.03) | 0.99 (0.95 - 1.03) | 0.582 | 0.99 (0.95 -1.03) |
| Birth_year2009:Country_origin_thirdgenMorocco | 1.00 (0.96 - 1.03) | 0.800 | 1.00 (0.96 - 1.03) | 1.00 (0.96 - 1.03) | 0.889 | 1.00 (0.97 -1.03) |

| | | | | | | |
|---|---|---|---|---|---|---|
| Birth_year2010:Country_origin_thirdgenMorocco | 1.00 (0.97 - 1.04) | 0.853 | 1.00 (0.97 - 1.04) | 1.01 (0.97 - 1.04) | 0.728 | 1.01 (0.97 -1.04) |
| Birth_year2011:Country_origin_thirdgenMorocco | 1.00 (0.96 - 1.03) | 0.855 | 1.00 (0.97 - 1.03) | 1.00 (0.97 - 1.04) | 0.877 | 1.00 (0.97 -1.04) |
| Birth_year2012:Country_origin_thirdgenMorocco | 1.00 (0.96 - 1.03) | 0.829 | 1.00 (0.96 - 1.03) | 1.00 (0.97 - 1.04) | 0.840 | 1.01 (0.97 -1.04) |
| Birth_year2013:Country_origin_thirdgenMorocco | 0.99 (0.95 - 1.02) | 0.403 | 0.99 (0.95 - 1.02) | 0.99 (0.96 - 1.03) | 0.726 | 1.00 (0.96 -1.03) |
| Birth_year2014:Country_origin_thirdgenMorocco | 0.97 (0.94 - 1.00) | 0.071 | 0.97 (0.93 - 1.00) | 0.98 (0.95 - 1.02) | 0.348 | 0.99 (0.95 -1.02) |
| Birth_year2015:Country_origin_thirdgenMorocco | 0.93 (0.90 - 0.96) | <0.001 | 0.93 (0.90 - 0.97) | 0.95 (0.92 - 0.98) | 0.003 | 0.95 (0.92 -0.98) |
| Birth_year2016:Country_origin_thirdgenMorocco | 0.91 (0.87 - 0.94) | <0.001 | 0.90 (0.87 - 0.94) | 0.92 (0.89 - 0.95) | <0.001 | 0.92 (0.89 -0.96) |
| Birth_year2017:Country_origin_thirdgenMorocco | 0.90 (0.87 - 0.93) | <0.001 | 0.90 (0.87 - 0.94) | 0.91 (0.88 - 0.94) | <0.001 | 0.91 (0.88 -0.95) |
| Birth_year2018:Country_origin_thirdgenMorocco | 0.87 (0.84 - 0.90) | <0.001 | 0.87 (0.84 - 0.91) | 0.88 (0.85 - 0.92) | <0.001 | 0.89 (0.85 -0.92) |
| Birth_year2019:Country_origin_thirdgenMorocco | 0.79 (0.76 - 0.82) | <0.001 | 0.79 (0.76 - 0.83) | 0.83 (0.80 - 0.86) | <0.001 | 0.83 (0.79 -0.86) |
| Birth_year2020:Country_origin_thirdgenMorocco | 0.73 (0.70 - 0.75) | <0.001 | 0.73 (0.69 - 0.77) | 0.75 (0.72 - 0.78) | <0.001 | 0.75 (0.71 -0.79) |
| Birth_year2009:Country_origin_thirdgenOther, Africa | 1.01 (0.97 - 1.05) | 0.684 | 1.01 (0.97 - 1.05) | 1.01 (0.97 - 1.06) | 0.586 | 1.01 (0.96 -1.05) |
| Birth_year2010:Country_origin_thirdgenOther, Africa | 1.02 (0.98 - 1.06) | 0.373 | 1.03 (0.99 - 1.07) | 1.03 (0.99 - 1.07) | 0.202 | 1.03 (0.99 -1.07) |
| Birth_year2011:Country_origin_thirdgenOther, Africa | 1.03 (0.99 - 1.08) | 0.142 | 1.04 (1.00 - 1.08) | 1.05 (1.00 - 1.09) | 0.032 | 1.05 (1.01 -1.10) |
| Birth_year2012:Country_origin_thirdgenOther, Africa | 1.03 (0.98 - 1.07) | 0.222 | 1.04 (1.00 - 1.08) | 1.04 (1.00 - 1.09) | 0.057 | 1.05 (1.01 -1.09) |
| Birth_year2013:Country_origin_thirdgenOther, Africa | 1.04 (0.99 - 1.08) | 0.093 | 1.05 (1.01 - 1.09) | 1.04 (1.00 - 1.09) | 0.042 | 1.05 (1.01 -1.09) |
| Birth_year2014:Country_origin_thirdgenOther, Africa | 1.02 (0.98 - 1.07) | 0.306 | 1.04 (1.00 - 1.08) | 1.04 (1.00 - 1.09) | 0.054 | 1.05 (1.01 -1.09) |
| Birth_year2015:Country_origin_thirdgenOther, Africa | 1.03 (0.99 - 1.08) | 0.134 | 1.04 (1.00 - 1.08) | 1.05 (1.01 - 1.10) | 0.022 | 1.05 (1.01 -1.09) |
| Birth_year2016:Country_origin_thirdgenOther, Africa | 1.03 (0.99 - 1.08) | 0.146 | 1.04 (1.00 - 1.08) | 1.05 (1.01 - 1.09) | 0.025 | 1.05 (1.01 -1.09) |
| Birth_year2017:Country_origin_thirdgenOther, Africa | 1.03 (0.99 - 1.07) | 0.183 | 1.04 (1.00 - 1.08) | 1.05 (1.01 - 1.09) | 0.021 | 1.05 (1.01 -1.09) |
| Birth_year2018:Country_origin_thirdgenOther, Africa | 1.03 (0.99 - 1.07) | 0.126 | 1.04 (1.00 - 1.08) | 1.05 (1.01 - 1.09) | 0.020 | 1.05 (1.01 -1.09) |
| Birth_year2019:Country_origin_thirdgenOther, Africa | 1.02 (0.98 - 1.06) | 0.325 | 1.03 (0.99 - 1.07) | 1.04 (1.00 - 1.08) | 0.050 | 1.04 (1.00 -1.08) |
| Birth_year2020:Country_origin_thirdgenOther, Africa | 1.01 (0.97 - 1.05) | 0.707 | 1.02 (0.98 - 1.06) | 1.04 (1.00 - 1.08) | 0.071 | 1.03 (0.99 -1.07) |
| Birth_year2009:Country_origin_thirdgenOther, America/Oceania | 1.00 (0.95 - 1.05) | 0.907 | 1.00 (0.95 - 1.04) | 0.99 (0.94 - 1.04) | 0.694 | 1.00 (0.95 -1.05) |
| Birth_year2010:Country_origin_thirdgenOther, America/Oceania | 1.00 (0.96 - 1.05) | 0.892 | 1.00 (0.95 - 1.04) | 1.00 (0.95 - 1.05) | 0.944 | 1.00 (0.96 -1.05) |
| Birth_year2011:Country_origin_thirdgenOther, America/Oceania | 1.00 (0.96 - 1.05) | 0.920 | 1.00 (0.96 - 1.05) | 1.00 (0.96 - 1.05) | 0.882 | 1.01 (0.97 -1.06) |
| Birth_year2012:Country_origin_thirdgenOther, America/Oceania | 1.00 (0.95 - 1.05) | 0.988 | 1.00 (0.95 - 1.04) | 0.99 (0.95 - 1.04) | 0.794 | 1.00 (0.95 -1.04) |
| Birth_year2013:Country_origin_thirdgenOther, America/Oceania | 1.00 (0.96 - 1.05) | 0.839 | 1.00 (0.95 - 1.04) | 1.00 (0.96 - 1.05) | 0.918 | 1.01 (0.96 -1.06) |
| Birth_year2014:Country_origin_thirdgenOther, America/Oceania | 1.00 (0.96 - 1.05) | 0.912 | 1.00 (0.96 - 1.04) | 1.00 (0.96 - 1.05) | 0.855 | 1.02 (0.97 -1.06) |
| Birth_year2015:Country_origin_thirdgenOther, America/Oceania | 1.00 (0.96 - 1.05) | 0.938 | 1.00 (0.95 - 1.04) | 1.01 (0.96 - 1.05) | 0.828 | 1.02 (0.97 -1.06) |

| | | | | | | |
|---|---|---|---|---|---|---|
| Birth_year2016:Country_origin_thirdgenOther, America/Oceania | 1.01 (0.96 - 1.06) | 0.782 | 1.00 (0.96 - 1.04) | 1.01 (0.96 - 1.06) | 0.654 | 1.03 (0.99 -1.08) |
| Birth_year2017:Country_origin_thirdgenOther, America/Oceania | 1.01 (0.96 - 1.06) | 0.711 | 1.00 (0.96 - 1.05) | 1.01 (0.96 - 1.05) | 0.806 | 1.03 (0.98 -1.07) |
| Birth_year2018:Country_origin_thirdgenOther, America/Oceania | 1.01 (0.96 - 1.06) | 0.717 | 1.00 (0.96 - 1.04) | 1.01 (0.97 - 1.06) | 0.578 | 1.04 (0.99 -1.08) |
| Birth_year2019:Country_origin_thirdgenOther, America/Oceania | 1.00 (0.96 - 1.05) | 0.921 | 0.99 (0.95 - 1.04) | 1.01 (0.96 - 1.05) | 0.777 | 1.03 (0.99 -1.07) |
| Birth_year2020:Country_origin_thirdgenOther, America/Oceania | 1.00 (0.95 - 1.05) | 0.997 | 0.99 (0.95 - 1.04) | 1.00 (0.96 - 1.05) | 0.874 | 1.03 (0.99 -1.07) |
| Birth_year2009:Country_origin_thirdgenOther, Asia | 1.00 (0.97 - 1.04) | 0.887 | 1.00 (0.97 - 1.04) | 1.01 (0.97 - 1.05) | 0.623 | 1.00 (0.97 -1.04) |
| Birth_year2010:Country_origin_thirdgenOther, Asia | 1.01 (0.98 - 1.05) | 0.443 | 1.02 (0.98 - 1.05) | 1.02 (0.98 - 1.06) | 0.285 | 1.02 (0.99 -1.06) |
| Birth_year2011:Country_origin_thirdgenOther, Asia | 1.01 (0.98 - 1.05) | 0.445 | 1.02 (0.98 - 1.05) | 1.02 (0.99 - 1.06) | 0.180 | 1.02 (0.99 -1.06) |
| Birth_year2012:Country_origin_thirdgenOther, Asia | 1.01 (0.98 - 1.05) | 0.487 | 1.02 (0.98 - 1.05) | 1.03 (0.99 - 1.06) | 0.150 | 1.03 (1.00 -1.06) |
| Birth_year2013:Country_origin_thirdgenOther, Asia | 1.01 (0.98 - 1.05) | 0.434 | 1.02 (0.99 - 1.06) | 1.03 (0.99 - 1.06) | 0.137 | 1.03 (1.00 -1.07) |
| Birth_year2014:Country_origin_thirdgenOther, Asia | 1.01 (0.97 - 1.04) | 0.611 | 1.02 (0.98 - 1.05) | 1.02 (0.99 - 1.06) | 0.188 | 1.03 (1.00 -1.06) |
| Birth_year2015:Country_origin_thirdgenOther, Asia | 0.99 (0.96 - 1.03) | 0.726 | 1.01 (0.97 - 1.04) | 1.01 (0.98 - 1.05) | 0.495 | 1.02 (0.99 -1.06) |
| Birth_year2016:Country_origin_thirdgenOther, Asia | 1.01 (0.98 - 1.05) | 0.499 | 1.02 (0.99 - 1.05) | 1.03 (0.99 - 1.06) | 0.105 | 1.03 (1.00 -1.07) |
| Birth_year2017:Country_origin_thirdgenOther, Asia | 1.02 (0.99 - 1.06) | 0.186 | 1.03 (1.00 - 1.07) | 1.04 (1.01 - 1.08) | 0.020 | 1.05 (1.02 -1.08) |
| Birth_year2018:Country_origin_thirdgenOther, Asia | 1.02 (0.99 - 1.06) | 0.169 | 1.03 (1.00 - 1.07) | 1.04 (1.01 - 1.08) | 0.017 | 1.05 (1.02 -1.08) |
| Birth_year2019:Country_origin_thirdgenOther, Asia | 1.03 (0.99 - 1.06) | 0.134 | 1.04 (1.01 - 1.07) | 1.04 (1.01 - 1.08) | 0.014 | 1.05 (1.02 -1.08) |
| Birth_year2020:Country_origin_thirdgenOther, Asia | 1.03 (0.99 - 1.06) | 0.123 | 1.04 (1.01 - 1.07) | 1.04 (1.01 - 1.08) | 0.014 | 1.05 (1.02 -1.08) |
| Birth_year2009:Country_origin_thirdgenSuriname | 1.00 (0.96 - 1.04) | 0.992 | 1.00 (0.96 - 1.04) | 1.00 (0.97 - 1.04) | 0.863 | 1.01 (0.97 -1.04) |
| Birth_year2010:Country_origin_thirdgenSuriname | 1.00 (0.97 - 1.04) | 0.866 | 1.00 (0.97 - 1.04) | 1.01 (0.97 - 1.05) | 0.633 | 1.01 (0.97 -1.05) |
| Birth_year2011:Country_origin_thirdgenSuriname | 1.00 (0.96 - 1.04) | 0.887 | 1.00 (0.97 - 1.04) | 1.01 (0.97 - 1.05) | 0.546 | 1.02 (0.98 -1.05) |
| Birth_year2012:Country_origin_thirdgenSuriname | 1.01 (0.97 - 1.05) | 0.754 | 1.01 (0.97 - 1.04) | 1.01 (0.98 - 1.06) | 0.458 | 1.02 (0.98 -1.06) |
| Birth_year2013:Country_origin_thirdgenSuriname | 1.00 (0.97 - 1.04) | 0.835 | 1.00 (0.96 - 1.04) | 1.01 (0.97 - 1.05) | 0.514 | 1.02 (0.98 -1.05) |
| Birth_year2014:Country_origin_thirdgenSuriname | 1.01 (0.97 - 1.05) | 0.665 | 1.01 (0.97 - 1.05) | 1.02 (0.98 - 1.06) | 0.420 | 1.02 (0.98 -1.06) |
| Birth_year2015:Country_origin_thirdgenSuriname | 1.00 (0.96 - 1.04) | 0.826 | 1.00 (0.96 - 1.03) | 1.01 (0.97 - 1.05) | 0.570 | 1.01 (0.97 -1.05) |
| Birth_year2016:Country_origin_thirdgenSuriname | 0.98 (0.94 - 1.02) | 0.313 | 0.98 (0.94 - 1.02) | 0.99 (0.96 - 1.03) | 0.739 | 0.99 (0.95 -1.03) |
| Birth_year2017:Country_origin_thirdgenSuriname | 0.99 (0.95 - 1.03) | 0.517 | 0.99 (0.95 - 1.03) | 1.00 (0.96 - 1.04) | 0.912 | 1.00 (0.96 -1.04) |
| Birth_year2018:Country_origin_thirdgenSuriname | 0.97 (0.94 - 1.01) | 0.177 | 0.97 (0.93 - 1.01) | 0.99 (0.95 - 1.03) | 0.515 | 0.99 (0.95 -1.03) |
| Birth_year2019:Country_origin_thirdgenSuriname | 0.95 (0.92 - 0.99) | 0.020 | 0.95 (0.91 - 0.99) | 0.98 (0.94 - 1.01) | 0.216 | 0.97 (0.93 -1.01) |
| Birth_year2020:Country_origin_thirdgenSuriname | 0.93 (0.90 - 0.97) | 0.001 | 0.93 (0.89 - 0.97) | 0.95 (0.92 - 0.99) | 0.024 | 0.95 (0.91 -0.99) |
| Birth_year2009:Country_origin_thirdgenThe Dutch Caribbean | 1.01 (0.96 - 1.06) | 0.772 | 1.01 (0.96 - 1.06) | 1.01 (0.96 - 1.07) | 0.596 | 1.02 (0.97 -1.07) |

| | | | | | | |
|---|---|---|---|---|---|---|
| Birth_year2010:Country_origin_thirdgenThe Dutch Caribbean | 1.00 (0.95 - 1.06) | 0.852 | 1.01 (0.96 - 1.06) | 1.01 (0.96 - 1.06) | 0.704 | 1.01 (0.96 -1.06) |
| Birth_year2011:Country_origin_thirdgenThe Dutch Caribbean | 1.01 (0.96 - 1.06) | 0.727 | 1.01 (0.96 - 1.06) | 1.01 (0.96 - 1.07) | 0.595 | 1.02 (0.97 -1.07) |
| Birth_year2012:Country_origin_thirdgenThe Dutch Caribbean | 1.01 (0.96 - 1.06) | 0.817 | 1.01 (0.96 - 1.06) | 1.01 (0.96 - 1.07) | 0.642 | 1.01 (0.96 -1.06) |
| Birth_year2013:Country_origin_thirdgenThe Dutch Caribbean | 1.01 (0.96 - 1.06) | 0.733 | 1.01 (0.96 - 1.06) | 1.02 (0.97 - 1.08) | 0.431 | 1.02 (0.97 -1.07) |
| Birth_year2014:Country_origin_thirdgenThe Dutch Caribbean | 1.01 (0.96 - 1.06) | 0.708 | 1.01 (0.96 - 1.06) | 1.02 (0.97 - 1.08) | 0.387 | 1.01 (0.96 -1.06) |
| Birth_year2015:Country_origin_thirdgenThe Dutch Caribbean | 1.00 (0.95 - 1.06) | 0.892 | 1.00 (0.95 - 1.05) | 1.02 (0.97 - 1.07) | 0.499 | 1.01 (0.96 -1.06) |
| Birth_year2016:Country_origin_thirdgenThe Dutch Caribbean | 0.98 (0.93 - 1.03) | 0.506 | 0.98 (0.93 - 1.03) | 1.00 (0.95 - 1.05) | 0.937 | 0.99 (0.94 -1.04) |
| Birth_year2017:Country_origin_thirdgenThe Dutch Caribbean | 0.99 (0.94 - 1.04) | 0.673 | 0.99 (0.94 - 1.04) | 1.00 (0.95 - 1.05) | 0.980 | 0.99 (0.94 -1.04) |
| Birth_year2018:Country_origin_thirdgenThe Dutch Caribbean | 0.97 (0.92 - 1.02) | 0.273 | 0.97 (0.92 - 1.02) | 0.98 (0.94 - 1.04) | 0.555 | 0.97 (0.92 -1.02) |
| Birth_year2019:Country_origin_thirdgenThe Dutch Caribbean | 0.95 (0.90 - 1.00) | 0.062 | 0.95 (0.90 - 1.00) | 0.97 (0.92 - 1.02) | 0.268 | 0.96 (0.91 -1.01) |
| Birth_year2020:Country_origin_thirdgenThe Dutch Caribbean | 0.93 (0.88 - 0.98) | 0.004 | 0.93 (0.88 - 0.98) | 0.95 (0.91 - 1.00) | 0.071 | 0.94 (0.89 -0.99) |
| Birth_year2009:Country_origin_thirdgenTurkey | 1.01 (0.97 - 1.04) | 0.787 | 1.00 (0.97 - 1.04) | 1.01 (0.97 - 1.05) | 0.617 | 1.01 (0.97 -1.05) |
| Birth_year2010:Country_origin_thirdgenTurkey | 1.01 (0.98 - 1.05) | 0.532 | 1.01 (0.98 - 1.05) | 1.02 (0.98 - 1.06) | 0.385 | 1.02 (0.98 -1.05) |
| Birth_year2011:Country_origin_thirdgenTurkey | 1.01 (0.97 - 1.05) | 0.585 | 1.01 (0.98 - 1.05) | 1.02 (0.98 - 1.05) | 0.394 | 1.02 (0.98 -1.06) |
| Birth_year2012:Country_origin_thirdgenTurkey | 1.01 (0.98 - 1.05) | 0.509 | 1.02 (0.98 - 1.05) | 1.02 (0.98 - 1.06) | 0.290 | 1.03 (0.99 -1.07) |
| Birth_year2013:Country_origin_thirdgenTurkey | 1.02 (0.98 - 1.05) | 0.431 | 1.02 (0.98 - 1.05) | 1.02 (0.98 - 1.06) | 0.304 | 1.02 (0.99 -1.06) |
| Birth_year2014:Country_origin_thirdgenTurkey | 1.01 (0.98 - 1.05) | 0.510 | 1.01 (0.98 - 1.05) | 1.02 (0.98 - 1.06) | 0.345 | 1.02 (0.98 -1.06) |
| Birth_year2015:Country_origin_thirdgenTurkey | 1.00 (0.96 - 1.03) | 0.844 | 1.00 (0.96 - 1.03) | 1.01 (0.97 - 1.05) | 0.611 | 1.01 (0.98 -1.05) |
| Birth_year2016:Country_origin_thirdgenTurkey | 0.98 (0.94 - 1.01) | 0.182 | 0.98 (0.94 - 1.01) | 0.99 (0.96 - 1.03) | 0.656 | 0.99 (0.96 -1.03) |
| Birth_year2017:Country_origin_thirdgenTurkey | 0.97 (0.93 - 1.00) | 0.067 | 0.97 (0.93 - 1.00) | 0.98 (0.95 - 1.02) | 0.336 | 0.98 (0.94 -1.02) |
| Birth_year2018:Country_origin_thirdgenTurkey | 0.96 (0.92 - 0.99) | 0.019 | 0.96 (0.92 - 0.99) | 0.97 (0.94 - 1.01) | 0.173 | 0.97 (0.94 -1.01) |
| Birth_year2019:Country_origin_thirdgenTurkey | 0.90 (0.87 - 0.94) | <0.001 | 0.90 (0.87 - 0.94) | 0.94 (0.90 - 0.97) | 0.001 | 0.94 (0.90 -0.97) |
| Birth_year2020:Country_origin_thirdgenTurkey | 0.87 (0.84 - 0.91) | <0.001 | 0.87 (0.83 - 0.91) | 0.90 (0.86 - 0.93) | <0.001 | 0.90 (0.86 -0.94) |
| Birth_year2009:SES.mother5Self-employed | 0.99 (0.97 - 1.02) | 0.667 | 0.99 (0.97 - 1.02) | 0.99 (0.97 - 1.02) | 0.564 | 0.99 (0.97 -1.02) |
| Birth_year2010:SES.mother5Self-employed | 0.99 (0.97 - 1.02) | 0.607 | 0.99 (0.97 - 1.02) | 0.99 (0.97 - 1.02) | 0.511 | 0.99 (0.96 -1.02) |
| Birth_year2011:SES.mother5Self-employed | 0.99 (0.97 - 1.02) | 0.688 | 0.99 (0.97 - 1.02) | 0.99 (0.96 - 1.02) | 0.478 | 0.99 (0.96 -1.02) |

| | | | | | | |
|---|---|---|---|---|---|---|
| Birth_year2012:SES.mother5Self-employed | 0.99 (0.97 - 1.02) | 0.515 | 0.99 (0.96 - 1.02) | 0.99 (0.96 - 1.01) | 0.286 | 0.99 (0.96 -1.01) |
| Birth_year2013:SES.mother5Self-employed | 0.99 (0.96 - 1.01) | 0.269 | 0.98 (0.96 - 1.01) | 0.98 (0.95 - 1.01) | 0.140 | 0.98 (0.95 -1.01) |
| Birth_year2014:SES.mother5Self-employed | 0.98 (0.95 - 1.00) | 0.097 | 0.98 (0.95 - 1.00) | 0.97 (0.95 - 1.00) | 0.059 | 0.97 (0.95 -1.00) |
| Birth_year2015:SES.mother5Self-employed | 0.97 (0.95 - 1.00) | 0.055 | 0.97 (0.95 - 1.00) | 0.97 (0.95 - 1.00) | 0.034 | 0.97 (0.95 -1.00) |
| Birth_year2016:SES.mother5Self-employed | 0.97 (0.95 - 1.00) | 0.033 | 0.97 (0.95 - 1.00) | 0.97 (0.94 - 0.99) | 0.012 | 0.97 (0.94 -0.99) |
| Birth_year2017:SES.mother5Self-employed | 0.99 (0.96 - 1.01) | 0.312 | 0.99 (0.96 - 1.01) | 0.98 (0.95 - 1.00) | 0.097 | 0.98 (0.95 -1.00) |
| Birth_year2018:SES.mother5Self-employed | 0.98 (0.96 - 1.01) | 0.193 | 0.98 (0.96 - 1.01) | 0.97 (0.95 - 1.00) | 0.057 | 0.97 (0.95 -1.00) |
| Birth_year2019:SES.mother5Self-employed | 0.98 (0.95 - 1.01) | 0.118 | 0.98 (0.95 - 1.01) | 0.97 (0.95 - 1.00) | 0.046 | 0.97 (0.95 -1.00) |
| Birth_year2020:SES.mother5Self-employed | 0.97 (0.94 - 1.00) | 0.021 | 0.97 (0.94 - 1.00) | 0.96 (0.94 - 0.99) | 0.006 | 0.96 (0.94 -0.99) |
| Birth_year2009:SES.mother5Recipient benefit | 1.01 (0.98 - 1.03) | 0.645 | 1.01 (0.98 - 1.03) | 1.00 (0.98 - 1.03) | 0.770 | 1.00 (0.98 -1.03) |
| Birth_year2010:SES.mother5Recipient benefit | 1.01 (0.99 - 1.04) | 0.291 | 1.01 (0.99 - 1.03) | 1.01 (0.99 - 1.03) | 0.382 | 1.01 (0.99 -1.03) |
| Birth_year2011:SES.mother5Recipient benefit | 1.01 (0.99 - 1.03) | 0.388 | 1.01 (0.99 - 1.03) | 1.01 (0.98 - 1.03) | 0.512 | 1.01 (0.98 -1.03) |
| Birth_year2012:SES.mother5Recipient benefit | 1.01 (0.99 - 1.04) | 0.265 | 1.01 (0.99 - 1.04) | 1.01 (0.99 - 1.04) | 0.258 | 1.01 (0.99 -1.04) |
| Birth_year2013:SES.mother5Recipient benefit | 1.01 (0.99 - 1.03) | 0.390 | 1.01 (0.99 - 1.03) | 1.01 (0.99 - 1.04) | 0.312 | 1.01 (0.99 -1.03) |
| Birth_year2014:SES.mother5Recipient benefit | 1.00 (0.98 - 1.02) | 0.988 | 1.00 (0.98 - 1.02) | 1.01 (0.98 - 1.03) | 0.508 | 1.01 (0.98 -1.03) |
| Birth_year2015:SES.mother5Recipient benefit | 0.99 (0.96 - 1.01) | 0.323 | 0.99 (0.96 - 1.01) | 1.00 (0.97 - 1.02) | 0.897 | 1.00 (0.97 -1.02) |
| Birth_year2016:SES.mother5Recipient benefit | 1.00 (0.98 - 1.03) | 0.950 | 1.00 (0.98 - 1.03) | 1.01 (0.98 - 1.03) | 0.537 | 1.01 (0.98 -1.03) |
| Birth_year2017:SES.mother5Recipient benefit | 1.01 (0.99 - 1.04) | 0.404 | 1.01 (0.99 - 1.03) | 1.01 (0.99 - 1.04) | 0.383 | 1.01 (0.99 -1.04) |
| Birth_year2018:SES.mother5Recipient benefit | 1.01 (0.99 - 1.04) | 0.234 | 1.02 (0.99 - 1.04) | 1.02 (0.99 - 1.04) | 0.136 | 1.02 (0.99 -1.04) |
| Birth_year2019:SES.mother5Recipient benefit | 1.01 (0.99 - 1.04) | 0.315 | 1.01 (0.99 - 1.04) | 1.02 (0.99 - 1.05) | 0.120 | 1.02 (1.00 -1.05) |
| Birth_year2020:SES.mother5Recipient benefit | 1.01 (0.98 - 1.03) | 0.548 | 1.01 (0.98 - 1.03) | 1.01 (0.99 - 1.04) | 0.342 | 1.01 (0.99 -1.04) |
| Birth_year2009:SES.mother5Pensioner | 1.01 (0.94 - 1.09) | 0.820 | 1.01 (0.93 - 1.08) | 1.01 (0.94 - 1.09) | 0.719 | 1.01 (0.94 -1.09) |
| Birth_year2010:SES.mother5Pensioner | 1.01 (0.93 - 1.09) | 0.807 | 1.01 (0.93 - 1.09) | 1.01 (0.93 - 1.09) | 0.872 | 1.01 (0.93 -1.08) |
| Birth_year2011:SES.mother5Pensioner | 1.00 (0.92 - 1.09) | 0.907 | 1.00 (0.92 - 1.09) | 0.99 (0.92 - 1.08) | 0.906 | 1.00 (0.91 -1.08) |
| Birth_year2012:SES.mother5Pensioner | 1.00 (0.92 - 1.10) | 0.966 | 1.00 (0.91 - 1.09) | 0.99 (0.90 - 1.08) | 0.820 | 0.99 (0.90 -1.08) |
| Birth_year2013:SES.mother5Pensioner | 0.98 (0.90 - 1.07) | 0.718 | 0.98 (0.90 - 1.07) | 0.99 (0.91 - 1.08) | 0.844 | 0.99 (0.90 -1.08) |
| Birth_year2014:SES.mother5Pensioner | 0.99 (0.91 - 1.08) | 0.864 | 0.99 (0.90 - 1.08) | 0.99 (0.90 - 1.08) | 0.783 | 0.99 (0.90 -1.08) |
| Birth_year2015:SES.mother5Pensioner | 0.97 (0.88 - 1.07) | 0.555 | 0.97 (0.87 - 1.07) | 0.97 (0.88 - 1.08) | 0.610 | 0.97 (0.87 -1.07) |
| Birth_year2016:SES.mother5Pensioner | 1.00 (0.90 - 1.11) | 0.994 | 1.00 (0.89 - 1.10) | 0.99 (0.89 - 1.10) | 0.791 | 0.98 (0.88 -1.09) |
| Birth_year2017:SES.mother5Pensioner | 1.00 (0.89 - 1.11) | 0.941 | 0.99 (0.88 - 1.10) | 0.99 (0.89 - 1.10) | 0.844 | 0.98 (0.87 -1.09) |
| Birth_year2018:SES.mother5Pensioner | 1.01 (0.90 - 1.12) | 0.913 | 1.01 (0.90 - 1.12) | 1.00 (0.89 - 1.11) | 0.935 | 1.00 (0.88 -1.11) |
| Birth_year2019:SES.mother5Pensioner | 1.03 (0.90 - 1.17) | 0.697 | 1.03 (0.90 - 1.16) | 1.02 (0.89 - 1.16) | 0.801 | 1.02 (0.89 -1.15) |
| Birth_year2020:SES.mother5Pensioner | 0.98 (0.85 - 1.13) | 0.786 | 0.98 (0.83 - 1.12) | 0.97 (0.84 - 1.12) | 0.639 | 0.97 (0.82 -1.11) |
| Birth_year2009:SES.mother5Child/student | 1.00 (0.95 - 1.05) | 0.977 | 1.00 (0.95 - 1.05) | 0.99 (0.94 - 1.04) | 0.735 | 0.99 (0.94 -1.04) |
| Birth_year2010:SES.mother5Child/student | 1.01 (0.96 - 1.06) | 0.810 | 1.00 (0.95 - 1.06) | 1.00 (0.95 - 1.05) | 0.982 | 1.00 (0.95 -1.05) |
| Birth_year2011:SES.mother5Child/student | 1.01 (0.95 - 1.06) | 0.846 | 1.00 (0.95 - 1.06) | 1.00 (0.95 - 1.05) | 0.945 | 1.00 (0.94 -1.05) |

| | | | | | | |
|---|---|---|---|---|---|---|
| Birth_year2012:SES.mother5Child/student | 1.00 (0.95 - 1.06) | 0.910 | 1.00 (0.95 - 1.05) | 0.99 (0.94 - 1.05) | 0.803 | 0.99 (0.94 -1.05) |
| Birth_year2013:SES.mother5Child/student | 1.01 (0.95 - 1.06) | 0.824 | 1.00 (0.95 - 1.06) | 1.01 (0.95 - 1.06) | 0.791 | 1.01 (0.95 -1.06) |
| Birth_year2014:SES.mother5Child/student | 0.99 (0.94 - 1.05) | 0.731 | 0.99 (0.93 - 1.04) | 0.99 (0.94 - 1.05) | 0.795 | 0.99 (0.94 -1.05) |
| Birth_year2015:SES.mother5Child/student | 0.97 (0.91 - 1.02) | 0.227 | 0.96 (0.91 - 1.02) | 0.97 (0.92 - 1.03) | 0.318 | 0.97 (0.91 -1.03) |
| Birth_year2016:SES.mother5Child/student | 0.97 (0.92 - 1.03) | 0.333 | 0.97 (0.91 - 1.03) | 0.97 (0.92 - 1.03) | 0.378 | 0.97 (0.92 -1.03) |
| Birth_year2017:SES.mother5Child/student | 0.99 (0.94 - 1.05) | 0.737 | 0.99 (0.93 - 1.05) | 0.99 (0.94 - 1.05) | 0.792 | 0.99 (0.94 -1.05) |
| Birth_year2018:SES.mother5Child/student | 0.99 (0.94 - 1.05) | 0.744 | 0.99 (0.93 - 1.05) | 0.98 (0.93 - 1.04) | 0.598 | 0.99 (0.93 -1.04) |
| Birth_year2019:SES.mother5Child/student | 0.98 (0.93 - 1.04) | 0.531 | 0.98 (0.92 - 1.04) | 0.98 (0.93 - 1.04) | 0.548 | 0.98 (0.93 -1.04) |
| Birth_year2020:SES.mother5Child/student | 0.96 (0.90 - 1.02) | 0.157 | 0.96 (0.90 - 1.02) | 0.97 (0.91 - 1.03) | 0.274 | 0.97 (0.91 -1.03) |
| Birth_year2009:SES.mother5Other | 1.00 (0.97 - 1.03) | 0.973 | 1.00 (0.97 - 1.03) | 1.00 (0.97 - 1.03) | 0.941 | 1.00 (0.97 -1.02) |
| Birth_year2010:SES.mother5Other | 1.00 (0.98 - 1.03) | 0.926 | 1.00 (0.98 - 1.03) | 1.00 (0.98 - 1.03) | 0.935 | 1.00 (0.98 -1.03) |
| Birth_year2011:SES.mother5Other | 1.00 (0.98 - 1.03) | 0.739 | 1.00 (0.98 - 1.03) | 1.01 (0.98 - 1.03) | 0.607 | 1.01 (0.98 -1.03) |
| Birth_year2012:SES.mother5Other | 1.00 (0.98 - 1.03) | 0.824 | 1.00 (0.98 - 1.03) | 1.00 (0.98 - 1.03) | 0.841 | 1.00 (0.98 -1.03) |
| Birth_year2013:SES.mother5Other | 1.00 (0.98 - 1.03) | 0.757 | 1.00 (0.98 - 1.03) | 1.01 (0.98 - 1.03) | 0.639 | 1.00 (0.98 -1.03) |
| Birth_year2014:SES.mother5Other | 1.00 (0.98 - 1.03) | 0.848 | 1.00 (0.97 - 1.03) | 1.00 (0.98 - 1.03) | 0.871 | 1.00 (0.97 -1.03) |
| Birth_year2015:SES.mother5Other | 1.01 (0.98 - 1.03) | 0.710 | 1.00 (0.98 - 1.03) | 1.00 (0.98 - 1.03) | 0.774 | 1.00 (0.98 -1.03) |
| Birth_year2016:SES.mother5Other | 1.02 (0.99 - 1.04) | 0.293 | 1.01 (0.99 - 1.04) | 1.01 (0.98 - 1.04) | 0.488 | 1.01 (0.98 -1.04) |
| Birth_year2017:SES.mother5Other | 1.01 (0.99 - 1.04) | 0.338 | 1.01 (0.98 - 1.04) | 1.01 (0.98 - 1.04) | 0.519 | 1.01 (0.98 -1.04) |
| Birth_year2018:SES.mother5Other | 1.01 (0.99 - 1.04) | 0.332 | 1.01 (0.98 - 1.04) | 1.02 (0.99 - 1.05) | 0.318 | 1.01 (0.98 -1.04) |
| Birth_year2019:SES.mother5Other | 1.03 (1.00 - 1.06) | 0.056 | 1.03 (1.00 - 1.06) | 1.03 (1.00 - 1.06) | 0.064 | 1.03 (1.00 -1.06) |
| Birth_year2020:SES.mother5Other | 1.05 (1.02 - 1.08) | 0.003 | 1.04 (1.01 - 1.08) | 1.05 (1.01 - 1.08) | 0.005 | 1.04 (1.01 -1.07) |
| Birth_year2009:SES.mother5Unknown | NA | NA | 0.99 (0.88 - 1.09) | NA | NA | 1.01 (0.89 -1.12) |
| Birth_year2010:SES.mother5Unknown | NA | NA | 1.02 (0.91 - 1.12) | NA | NA | 1.06 (0.95 -1.17) |
| Birth_year2011:SES.mother5Unknown | NA | NA | 1.02 (0.91 - 1.13) | NA | NA | 1.07 (0.95 -1.18) |
| Birth_year2012:SES.mother5Unknown | NA | NA | 1.02 (0.91 - 1.13) | NA | NA | 1.08 (0.96 -1.19) |
| Birth_year2013:SES.mother5Unknown | NA | NA | 1.06 (0.95 - 1.17) | NA | NA | 1.08 (0.96 -1.19) |
| Birth_year2014:SES.mother5Unknown | NA | NA | 1.06 (0.95 - 1.17) | NA | NA | 1.08 (0.96 -1.19) |
| Birth_year2015:SES.mother5Unknown | NA | NA | 0.98 (0.86 - 1.10) | NA | NA | 1.05 (0.93 -1.17) |
| Birth_year2016:SES.mother5Unknown | NA | NA | 1.00 (0.88 - 1.12) | NA | NA | 1.03 (0.90 -1.15) |
| Birth_year2017:SES.mother5Unknown | NA | NA | 1.01 (0.89 - 1.14) | NA | NA | 1.06 (0.93 -1.19) |
| Birth_year2018:SES.mother5Unknown | NA | NA | 1.02 (0.90 - 1.15) | NA | NA | 1.03 (0.90 -1.16) |
| Birth_year2019:SES.mother5Unknown | NA | NA | 1.03 (0.90 - 1.15) | NA | NA | 1.07 (0.94 -1.20) |
| Birth_year2020:SES.mother5Unknown | NA | NA | 0.95 (0.84 - 1.07) | NA | NA | 0.86 (0.74 -0.98) |
| Birth_year2009:UrbanisationExtremely urbanised | 1.00 (0.98 - 1.03) | 0.744 | 1.00 (0.98 - 1.03) | 1.00 (0.98 - 1.03) | 0.783 | 1.00 (0.98 -1.03) |
| Birth_year2010:UrbanisationExtremely urbanised | 1.00 (0.98 - 1.03) | 0.761 | 1.00 (0.98 - 1.03) | 1.01 (0.98 - 1.03) | 0.490 | 1.01 (0.98 -1.03) |
| Birth_year2011:UrbanisationExtremely urbanised | 1.01 (0.98 - 1.03) | 0.533 | 1.01 (0.98 - 1.03) | 1.01 (0.99 - 1.04) | 0.377 | 1.01 (0.99 -1.03) |

| | | | | | | |
|---|---|---|---|---|---|---|
| Birth_year2012:UrbanisationExtremely urbanised | 1.01 (0.99 - 1.03) | 0.445 | 1.01 (0.99 - 1.03) | 1.01 (0.99 - 1.04) | 0.305 | 1.01 (0.99 -1.04) |
| Birth_year2013:UrbanisationExtremely urbanised | 1.01 (0.99 - 1.04) | 0.296 | 1.01 (0.99 - 1.04) | 1.01 (0.99 - 1.04) | 0.261 | 1.01 (0.99 -1.04) |
| Birth_year2014:UrbanisationExtremely urbanised | 1.01 (0.99 - 1.04) | 0.307 | 1.01 (0.99 - 1.04) | 1.01 (0.99 - 1.04) | 0.270 | 1.01 (0.99 -1.04) |
| Birth_year2015:UrbanisationExtremely urbanised | 1.02 (0.99 - 1.04) | 0.148 | 1.02 (0.99 - 1.04) | 1.02 (0.99 - 1.04) | 0.144 | 1.02 (0.99 -1.04) |
| Birth_year2016:UrbanisationExtremely urbanised | 1.02 (0.99 - 1.04) | 0.121 | 1.02 (1.00 - 1.05) | 1.02 (1.00 - 1.05) | 0.084 | 1.02 (1.00 -1.05) |
| Birth_year2017:UrbanisationExtremely urbanised | 1.01 (0.99 - 1.04) | 0.236 | 1.02 (0.99 - 1.04) | 1.02 (0.99 - 1.04) | 0.172 | 1.02 (0.99 -1.04) |
| Birth_year2018:UrbanisationExtremely urbanised | 1.02 (1.00 - 1.04) | 0.114 | 1.02 (1.00 - 1.05) | 1.02 (1.00 - 1.05) | 0.070 | 1.02 (1.00 -1.05) |
| Birth_year2019:UrbanisationExtremely urbanised | 1.01 (0.99 - 1.04) | 0.309 | 1.01 (0.99 - 1.04) | 1.02 (1.00 - 1.04) | 0.115 | 1.02 (1.00 -1.05) |
| Birth_year2020:UrbanisationExtremely urbanised | 1.01 (0.98 - 1.03) | 0.517 | 1.01 (0.99 - 1.03) | 1.01 (0.99 - 1.04) | 0.367 | 1.01 (0.99 -1.04) |
| Birth_year2009:UrbanisationHardly urbanised | 1.00 (0.98 - 1.03) | 0.808 | 1.00 (0.98 - 1.03) | 1.00 (0.98 - 1.03) | 0.737 | 1.00 (0.98 -1.03) |
| Birth_year2010:UrbanisationHardly urbanised | 1.00 (0.98 - 1.03) | 0.856 | 1.00 (0.98 - 1.02) | 1.01 (0.98 - 1.03) | 0.649 | 1.01 (0.98 -1.03) |
| Birth_year2011:UrbanisationHardly urbanised | 1.00 (0.98 - 1.03) | 0.768 | 1.00 (0.98 - 1.03) | 1.01 (0.98 - 1.03) | 0.616 | 1.01 (0.98 -1.03) |
| Birth_year2012:UrbanisationHardly urbanised | 1.00 (0.98 - 1.03) | 0.783 | 1.00 (0.98 - 1.03) | 1.01 (0.98 - 1.03) | 0.600 | 1.01 (0.98 -1.03) |
| Birth_year2013:UrbanisationHardly urbanised | 1.00 (0.97 - 1.02) | 0.876 | 1.00 (0.97 - 1.02) | 1.00 (0.97 - 1.02) | 0.890 | 1.00 (0.97 -1.02) |
| Birth_year2014:UrbanisationHardly urbanised | 1.00 (0.98 - 1.02) | 0.992 | 1.00 (0.98 - 1.02) | 1.00 (0.98 - 1.02) | 0.991 | 1.00 (0.98 -1.02) |
| Birth_year2015:UrbanisationHardly urbanised | 1.00 (0.97 - 1.02) | 0.766 | 1.00 (0.97 - 1.02) | 1.00 (0.97 - 1.02) | 0.887 | 1.00 (0.98 -1.02) |
| Birth_year2016:UrbanisationHardly urbanised | 1.00 (0.98 - 1.02) | 0.998 | 1.00 (0.98 - 1.02) | 1.00 (0.98 - 1.03) | 0.874 | 1.00 (0.98 -1.03) |
| Birth_year2017:UrbanisationHardly urbanised | 1.00 (0.98 - 1.02) | 0.936 | 1.00 (0.98 - 1.02) | 1.00 (0.98 - 1.03) | 0.853 | 1.00 (0.98 -1.03) |
| Birth_year2018:UrbanisationHardly urbanised | 1.01 (0.99 - 1.03) | 0.446 | 1.01 (0.99 - 1.03) | 1.01 (0.99 - 1.03) | 0.405 | 1.01 (0.99 -1.03) |
| Birth_year2019:UrbanisationHardly urbanised | 1.01 (0.98 - 1.03) | 0.606 | 1.01 (0.98 - 1.03) | 1.01 (0.99 - 1.03) | 0.477 | 1.01 (0.99 -1.03) |
| Birth_year2020:UrbanisationHardly urbanised | 1.01 (0.98 - 1.03) | 0.655 | 1.01 (0.98 - 1.03) | 1.01 (0.98 - 1.03) | 0.546 | 1.01 (0.98 -1.03) |
| Birth_year2009:UrbanisationModerately urbanised | 1.00 (0.98 - 1.02) | 0.868 | 1.00 (0.98 - 1.02) | 1.00 (0.98 - 1.02) | 0.950 | 1.00 (0.98 -1.02) |
| Birth_year2010:UrbanisationModerately urbanised | 1.00 (0.98 - 1.03) | 0.718 | 1.00 (0.98 - 1.03) | 1.00 (0.98 - 1.03) | 0.786 | 1.00 (0.98 -1.03) |
| Birth_year2011:UrbanisationModerately urbanised | 1.01 (0.98 - 1.03) | 0.622 | 1.01 (0.98 - 1.03) | 1.01 (0.98 - 1.03) | 0.586 | 1.01 (0.98 -1.03) |
| Birth_year2012:UrbanisationModerately urbanised | 1.01 (0.99 - 1.03) | 0.492 | 1.01 (0.99 - 1.03) | 1.01 (0.98 - 1.03) | 0.533 | 1.01 (0.98 -1.03) |
| Birth_year2013:UrbanisationModerately urbanised | 1.01 (0.98 - 1.03) | 0.555 | 1.01 (0.98 - 1.03) | 1.01 (0.98 - 1.03) | 0.613 | 1.01 (0.98 -1.03) |
| Birth_year2014:UrbanisationModerately urbanised | 1.01 (0.99 - 1.03) | 0.442 | 1.01 (0.99 - 1.03) | 1.01 (0.98 - 1.03) | 0.516 | 1.01 (0.99 -1.03) |
| Birth_year2015:UrbanisationModerately urbanised | 1.01 (0.99 - 1.03) | 0.357 | 1.01 (0.99 - 1.03) | 1.01 (0.98 - 1.03) | 0.602 | 1.01 (0.98 -1.03) |
| Birth_year2016:UrbanisationModerately urbanised | 1.01 (0.99 - 1.04) | 0.264 | 1.01 (0.99 - 1.04) | 1.01 (0.99 - 1.04) | 0.311 | 1.01 (0.99 -1.04) |
| Birth_year2017:UrbanisationModerately urbanised | 1.01 (0.99 - 1.03) | 0.455 | 1.01 (0.99 - 1.03) | 1.01 (0.98 - 1.03) | 0.550 | 1.01 (0.98 -1.03) |
| Birth_year2018:UrbanisationModerately urbanised | 1.01 (0.99 - 1.04) | 0.329 | 1.01 (0.99 - 1.04) | 1.01 (0.99 - 1.04) | 0.343 | 1.01 (0.99 -1.04) |
| Birth_year2019:UrbanisationModerately urbanised | 1.01 (0.98 - 1.03) | 0.517 | 1.01 (0.98 - 1.03) | 1.01 (0.98 - 1.03) | 0.560 | 1.01 (0.98 -1.03) |
| Birth_year2020:UrbanisationModerately urbanised | 1.01 (0.99 - 1.03) | 0.386 | 1.01 (0.99 - 1.03) | 1.01 (0.99 - 1.03) | 0.390 | 1.01 (0.99 -1.04) |
| Birth_year2009:UrbanisationStrongly urbanised | 1.00 (0.98 - 1.02) | 0.972 | 1.00 (0.98 - 1.02) | 1.00 (0.98 - 1.02) | 0.832 | 1.00 (0.98 -1.02) |
| Birth_year2010:UrbanisationStrongly urbanised | 1.00 (0.98 - 1.02) | 0.885 | 1.00 (0.98 - 1.02) | 1.00 (0.98 - 1.02) | 0.924 | 1.00 (0.98 -1.02) |
| Birth_year2011:UrbanisationStrongly urbanised | 1.00 (0.98 - 1.03) | 0.709 | 1.00 (0.98 - 1.03) | 1.00 (0.98 - 1.03) | 0.770 | 1.00 (0.98 -1.03) |

| | | | | | | |
|---|---|---|---|---|---|---|
| Birth_year2012:UrbanisationStrongly urbanised | 1.01 (0.98 - 1.03) | 0.645 | 1.01 (0.98 - 1.03) | 1.00 (0.98 - 1.03) | 0.704 | 1.00 (0.98 -1.03) |
| Birth_year2013:UrbanisationStrongly urbanised | 1.00 (0.98 - 1.03) | 0.754 | 1.00 (0.98 - 1.03) | 1.00 (0.98 - 1.03) | 0.824 | 1.00 (0.98 -1.02) |
| Birth_year2014:UrbanisationStrongly urbanised | 1.01 (0.98 - 1.03) | 0.553 | 1.01 (0.99 - 1.03) | 1.00 (0.98 - 1.03) | 0.791 | 1.00 (0.98 -1.03) |
| Birth_year2015:UrbanisationStrongly urbanised | 1.01 (0.99 - 1.03) | 0.414 | 1.01 (0.99 - 1.03) | 1.00 (0.98 - 1.03) | 0.671 | 1.00 (0.98 -1.03) |
| Birth_year2016:UrbanisationStrongly urbanised | 1.01 (0.99 - 1.03) | 0.401 | 1.01 (0.99 - 1.03) | 1.01 (0.98 - 1.03) | 0.526 | 1.01 (0.99 -1.03) |
| Birth_year2017:UrbanisationStrongly urbanised | 1.01 (0.99 - 1.03) | 0.439 | 1.01 (0.99 - 1.03) | 1.01 (0.98 - 1.03) | 0.569 | 1.01 (0.98 -1.03) |
| Birth_year2018:UrbanisationStrongly urbanised | 1.02 (0.99 - 1.04) | 0.150 | 1.02 (0.99 - 1.04) | 1.01 (0.99 - 1.04) | 0.219 | 1.01 (0.99 -1.04) |
| Birth_year2019:UrbanisationStrongly urbanised | 1.01 (0.99 - 1.03) | 0.461 | 1.01 (0.99 - 1.03) | 1.01 (0.99 - 1.03) | 0.438 | 1.01 (0.99 -1.03) |
| Birth_year2020:UrbanisationStrongly urbanised | 1.01 (0.99 - 1.04) | 0.307 | 1.01 (0.99 - 1.03) | 1.01 (0.99 - 1.03) | 0.439 | 1.01 (0.99 -1.03) |
| Birth_year2009:UrbanisationUnknown | NA | NA | 1.15 (0.60 - 1.70) | NA | NA | 1.25 (0.65 -1.86) |
| Birth_year2010:UrbanisationUnknown | NA | NA | 1.10 (0.53 - 1.68) | NA | NA | 1.08 (0.42 -1.73) |
| Birth_year2011:UrbanisationUnknown | NA | NA | 1.32 (0.72 - 1.93) | NA | NA | 1.48 (0.76 -2.19) |
| Birth_year2012:UrbanisationUnknown | NA | NA | 1.24 (0.52 - 1.96) | NA | NA | 1.92 (1.00 -2.85) |
| Birth_year2013:UrbanisationUnknown | NA | NA | 1.46 (0.75 - 2.16) | NA | NA | 1.71 (0.25 -3.17) |
| Birth_year2014:UrbanisationUnknown | NA | NA | 1.33 (0.69 - 1.97) | NA | NA | 1.54 (0.47 -2.61) |
| Birth_year2015:UrbanisationUnknown | NA | NA | 1.18 (0.45 - 1.90) | NA | NA | 0.97 (-1.04 -2.98) |
| Birth_year2016:UrbanisationUnknown | NA | NA | 1.41 (0.87 - 1.95) | NA | NA | 1.63 (0.97 -2.29) |
| Birth_year2017:UrbanisationUnknown | NA | NA | 1.40 (0.79 - 2.02) | NA | NA | 1.60 (0.68 -2.53) |
| Birth_year2018:UrbanisationUnknown | NA | NA | 1.15 (0.53 - 1.77) | NA | NA | 1.32 (0.45 -2.19) |
| Birth_year2019:UrbanisationUnknown | NA | NA | 1.36 (0.85 - 1.87) | NA | NA | 1.70 (1.07 -2.33) |
| Birth_year2020:UrbanisationUnknown | NA | NA | 1.33 (0.87 - 1.79) | NA | NA | 1.40 (0.69 -2.11) |
| Birth_year2009:family_size4 or more children | 1.01 (0.98 - 1.04) | 0.708 | 1.01 (0.98 - 1.03) | 1.00 (0.97 - 1.03) | 0.875 | 1.00 (0.97 -1.03) |
| Birth_year2010:family_size4 or more children | 1.01 (0.98 - 1.04) | 0.740 | 1.00 (0.97 - 1.03) | 1.00 (0.97 - 1.04) | 0.775 | 1.00 (0.97 -1.03) |
| Birth_year2011:family_size4 or more children | 1.01 (0.98 - 1.04) | 0.500 | 1.01 (0.98 - 1.04) | 1.01 (0.98 - 1.04) | 0.510 | 1.01 (0.98 -1.04) |
| Birth_year2012:family_size4 or more children | 1.00 (0.97 - 1.03) | 0.840 | 1.00 (0.97 - 1.03) | 0.99 (0.96 - 1.02) | 0.702 | 0.99 (0.96 -1.02) |
| Birth_year2013:family_size4 or more children | 1.01 (0.98 - 1.04) | 0.453 | 1.01 (0.98 - 1.04) | 1.01 (0.98 - 1.04) | 0.641 | 1.01 (0.98 -1.04) |
| Birth_year2014:family_size4 or more children | 1.00 (0.97 - 1.03) | 0.940 | 1.00 (0.97 - 1.03) | 1.00 (0.97 - 1.03) | 0.999 | 1.00 (0.97 -1.03) |
| Birth_year2015:family_size4 or more children | 1.00 (0.97 - 1.03) | 0.977 | 1.00 (0.97 - 1.03) | 1.00 (0.97 - 1.03) | 0.986 | 1.00 (0.97 -1.03) |
| Birth_year2016:family_size4 or more children | 1.01 (0.98 - 1.04) | 0.596 | 1.01 (0.98 - 1.04) | 1.01 (0.98 - 1.04) | 0.706 | 1.01 (0.98 -1.04) |
| Birth_year2017:family_size4 or more children | 1.02 (0.99 - 1.05) | 0.295 | 1.02 (0.99 - 1.05) | 1.01 (0.98 - 1.04) | 0.526 | 1.01 (0.98 -1.04) |
| Birth_year2018:family_size4 or more children | 1.01 (0.98 - 1.04) | 0.722 | 1.00 (0.97 - 1.03) | 1.00 (0.97 - 1.03) | 0.927 | 1.00 (0.97 -1.03) |
| Birth_year2019:family_size4 or more children | 1.00 (0.97 - 1.04) | 0.759 | 1.00 (0.97 - 1.03) | 1.00 (0.97 - 1.03) | 0.876 | 1.00 (0.97 -1.03) |
| Birth_year2020:family_size4 or more children | 0.99 (0.96 - 1.02) | 0.375 | 0.99 (0.95 - 1.02) | 0.98 (0.95 - 1.01) | 0.287 | 0.98 (0.95 -1.02) |
| Birth_year2009:family_sizeInstitutional | 1.11 (0.87 - 1.42) | 0.393 | 1.17 (0.98 - 1.37) | 1.13 (0.88 - 1.45) | 0.334 | 1.22 (1.01 -1.42) |
| Birth_year2010:family_sizeInstitutional | 1.07 (0.85 - 1.35) | 0.548 | 1.13 (0.95 - 1.31) | 1.12 (0.88 - 1.42) | 0.343 | 1.24 (1.05 -1.43) |
| Birth_year2011:family_sizeInstitutional | 1.06 (0.83 - 1.36) | 0.641 | 1.10 (0.92 - 1.29) | 1.10 (0.85 - 1.41) | 0.472 | 1.20 (1.00 -1.39) |

| | | | | | | |
|---|---|---|---|---|---|---|
| Birth_year2012:family_sizeInstitutional | 1.10 (0.86 - 1.41) | 0.435 | 1.14 (0.95 - 1.33) | 1.11 (0.86 - 1.43) | 0.425 | 1.20 (1.00 -1.39) |
| Birth_year2013:family_sizeInstitutional | 1.09 (0.86 - 1.39) | 0.457 | 1.13 (0.94 - 1.32) | 1.13 (0.89 - 1.44) | 0.314 | 1.20 (1.01 -1.39) |
| Birth_year2014:family_sizeInstitutional | 1.04 (0.82 - 1.32) | 0.737 | 1.07 (0.89 - 1.26) | 1.05 (0.82 - 1.34) | 0.712 | 1.13 (0.94 -1.32) |
| Birth_year2015:family_sizeInstitutional | 1.05 (0.82 - 1.35) | 0.718 | 1.05 (0.86 - 1.24) | 1.12 (0.87 - 1.44) | 0.387 | 1.15 (0.95 -1.35) |
| Birth_year2016:family_sizeInstitutional | 1.05 (0.82 - 1.35) | 0.673 | 1.03 (0.84 - 1.22) | 1.11 (0.87 - 1.42) | 0.391 | 1.10 (0.91 -1.30) |
| Birth_year2017:family_sizeInstitutional | 1.06 (0.82 - 1.37) | 0.644 | 1.03 (0.83 - 1.22) | 1.12 (0.87 - 1.44) | 0.377 | 1.11 (0.91 -1.30) |
| Birth_year2018:family_sizeInstitutional | 1.09 (0.85 - 1.39) | 0.498 | 1.03 (0.84 - 1.21) | 1.11 (0.87 - 1.42) | 0.386 | 1.08 (0.89 -1.27) |
| Birth_year2019:family_sizeInstitutional | 1.02 (0.79 - 1.32) | 0.878 | 0.95 (0.76 - 1.14) | 1.05 (0.81 - 1.35) | 0.729 | 1.00 (0.80 -1.19) |
| Birth_year2020:family_sizeInstitutional | 1.09 (0.86 - 1.40) | 0.475 | 1.08 (0.89 - 1.27) | 1.16 (0.90 - 1.48) | 0.250 | 1.11 (0.91 -1.30) |
| Birth_year2009:family_sizeUnknown | NA | NA | 0.92 (0.36 - 1.47) | NA | NA | 0.82 (0.22 -1.43) |
| Birth_year2010:family_sizeUnknown | NA | NA | 0.92 (0.34 - 1.50) | NA | NA | 0.95 (0.29 -1.60) |
| Birth_year2011:family_sizeUnknown | NA | NA | 0.80 (0.19 - 1.41) | NA | NA | 0.71 (0.00 -1.43) |
| Birth_year2012:family_sizeUnknown | NA | NA | 0.83 (0.10 - 1.56) | NA | NA | 0.55 (-0.38 -1.47) |
| Birth_year2013:family_sizeUnknown | NA | NA | 0.71 (0.01 - 1.42) | NA | NA | 0.63 (-0.84 -2.09) |
| Birth_year2014:family_sizeUnknown | NA | NA | 0.78 (0.13 - 1.43) | NA | NA | 0.69 (-0.39 -1.77) |
| Birth_year2015:family_sizeUnknown | NA | NA | 0.96 (0.23 - 1.69) | NA | NA | 1.18 (-0.83 -3.20) |
| Birth_year2016:family_sizeUnknown | NA | NA | 0.80 (0.25 - 1.35) | NA | NA | 0.75 (0.08 -1.41) |
| Birth_year2017:family_sizeUnknown | NA | NA | 0.80 (0.18 - 1.42) | NA | NA | 0.73 (-0.20 -1.66) |
| Birth_year2018:family_sizeUnknown | NA | NA | 0.99 (0.36 - 1.62) | NA | NA | 0.95 (0.08 -1.83) |
| Birth_year2019:family_sizeUnknown | NA | NA | 0.86 (0.34 - 1.37) | NA | NA | 0.76 (0.13 -1.40) |
| Birth_year2020:family_sizeUnknown | NA | NA | 0.86 (0.40 - 1.32) | NA | NA | 1.00 (0.29 -1.71) |
| Birth_year2009:Daycare2No daycare | 1.00 (0.98 - 1.01) | 0.792 | 1.00 (0.98 - 1.01) | 1.00 (0.98 - 1.01) | 0.783 | 1.00 (0.98 -1.01) |
| Birth_year2010:Daycare2No daycare | 1.00 (0.98 - 1.01) | 0.564 | 1.00 (0.98 - 1.01) | 0.99 (0.98 - 1.01) | 0.469 | 0.99 (0.98 -1.01) |
| Birth_year2011:Daycare2No daycare | 1.00 (0.98 - 1.01) | 0.642 | 1.00 (0.98 - 1.01) | 0.99 (0.98 - 1.01) | 0.499 | 1.00 (0.98 -1.01) |
| Birth_year2012:Daycare2No daycare | 0.99 (0.98 - 1.01) | 0.473 | 1.00 (0.98 - 1.01) | 0.99 (0.98 - 1.01) | 0.380 | 0.99 (0.98 -1.01) |
| Birth_year2013:Daycare2No daycare | 0.99 (0.97 - 1.01) | 0.227 | 0.99 (0.98 - 1.01) | 0.99 (0.98 - 1.01) | 0.262 | 0.99 (0.98 -1.01) |
| Birth_year2014:Daycare2No daycare | 0.98 (0.97 - 1.00) | 0.054 | 0.99 (0.97 - 1.00) | 0.98 (0.97 - 1.00) | 0.051 | 0.98 (0.97 -1.00) |
| Birth_year2015:Daycare2No daycare | 0.98 (0.96 - 1.00) | 0.017 | 0.98 (0.96 - 1.00) | 0.98 (0.96 - 1.00) | 0.015 | 0.98 (0.96 -1.00) |
| Birth_year2016:Daycare2No daycare | 0.97 (0.95 - 0.98) | <0.001 | 0.97 (0.95 - 0.98) | 0.97 (0.95 - 0.98) | <0.001 | 0.97 (0.95 -0.98) |
| Birth_year2017:Daycare2No daycare | 0.96 (0.94 - 0.98) | <0.001 | 0.96 (0.94 - 0.98) | 0.96 (0.94 - 0.98) | <0.001 | 0.96 (0.94 -0.98) |
| Birth_year2018:Daycare2No daycare | 0.95 (0.94 - 0.97) | <0.001 | 0.95 (0.93 - 0.97) | 0.95 (0.94 - 0.97) | <0.001 | 0.95 (0.94 -0.97) |
| Birth_year2019:Daycare2No daycare | 0.94 (0.92 - 0.96) | <0.001 | 0.94 (0.92 - 0.96) | 0.94 (0.92 - 0.96) | <0.001 | 0.94 (0.92 -0.96) |
| Birth_year2020:Daycare2No daycare | 0.92 (0.90 - 0.93) | <0.001 | 0.92 (0.90 - 0.94) | 0.93 (0.91 - 0.94) | <0.001 | 0.93 (0.91 -0.95) |

**Supplementary Table S5. Adjusted relative change in DTaP-IPV vaccination coverage**

| | Birth cohort | | | | | | | | | | | |
|---|---|---|---|---|---|---|---|---|---|---|---|---|
| | 2009 | 2010 | 2011 | 2012 | 2013 | 2014 | 2015 | 2016 | 2017 | 2018 | 2019 | 2020 |
| Education level mother (ref: high) | | | | | | | | | | | | |
| Not high | -0,05 | -0,05 | -0,21 | -0,50 | -0,11 | -0,27 | -0,65 | -0,95 | -0,87 | -0,88 | -0,76 | -1,7 |
| SDI quartiles (ref: fourth quartile) | | | | | | | | | | | | |
| First | -1,45 | -1,83 | -1,94 | -1,99 | -2,54 | -2,99 | -4,03 | -4,83 | -4,65 | -4,29 | -4,90 | -6,13 |
| Second | -0,42 | -1,36 | -1,53 | -1,80 | -2,17 | -2,85 | -3,67 | -4,44 | -3,78 | -4,27 | -4,09 | -4,67 |
| Third | 0,08 | -0,51 | -0,58 | -0,96 | -0,96 | -1,60 | -2,41 | -2,70 | -2,15 | -2,05 | -1,85 | -2,08 |
| Income source mother (ref: job in employment) | | | | | | | | | | | | |
| Self-employed | -3,35 | -3,79 | -4,09 | -4,75 | -5,21 | -6,17 | -6,76 | -7,43 | -5,94 | -6,39 | -6,23 | -7,39 |
| Benefit recipient | -0,83 | -0,50 | -1,00 | -0,63 | -0,70 | -1,57 | -2,84 | -2,1 | -1,36 | -0,73 | -0,31 | -1,21 |
| Pensioner | 0,19 | -0,90 | -2,25 | -2,99 | -2,76 | -3,58 | -5,19 | -4,21 | -3,49 | -3,01 | -0,63 | -5,74 |
| Student | -2,55 | -1,95 | -2,42 | -3,10 | -1,67 | -3,55 | -5,95 | -5,80 | -3,64 | -4,53 | -4,50 | -6,02 |
| Other | -5,40 | -5,54 | -5,23 | -5,81 | -5,40 | -6,23 | -6,37 | -5,97 | -5,60 | -5,22 | -3,70 | -2,31 |
| Country of origin (ref: The Netherlands) | | | | | | | | | | | | |
| Europe excl NL | -1,32 | -1,09 | -1,34 | -1,67 | -1,59 | -1,99 | -2,19 | -2,52 | -2,22 | -2,43 | -2,06 | -2,44 |
| Indonesia | 0,88 | 0,58 | -0,34 | 0,16 | 0,02 | -0,36 | -1,15 | -1,18 | -0,52 | -1,00 | -0,82 | -2,16 |
| Morocco | 3,85 | 4,36 | 3,77 | 3,64 | 2,70 | 1,17 | -2,81 | -5,74 | -6,26 | -9,18 | -15,03 | -22,83 |
| Other, Africa | -0,73 | 0,48 | 2,11 | 1,37 | 1,74 | 1,01 | 1,50 | 1,15 | 1,73 | 1,51 | 1,03 | 0,57 |
| Other, America/Oceania | -2,54 | -2,08 | -1,78 | -2,97 | -2,04 | -2,31 | -2,55 | -2,17 | -2,23 | -1,65 | -2,01 | -2,47 |
| Other, Asia | 0,52 | 1,22 | 1,48 | 1,43 | 1,58 | 0,76 | -0,71 | 0,71 | 2,39 | 2,27 | 2,69 | 2,55 |
| Suriname | 0,58 | 0,82 | 0,84 | 0,92 | 0,81 | 0,65 | -0,15 | -2,10 | -0,80 | -2,46 | -3,34 | -5,54 |
| The Dutch Caribbean | 0,83 | 0,08 | 0,25 | -0,14 | 0,78 | 0,48 | -0,32 | -2,44 | -1,75 | -3,46 | -4,50 | -6,39 |
| Turkey | 3,54 | 3,89 | 3,62 | 3,81 | 3,83 | 3,15 | 1,99 | -0,02 | -0,58 | -1,49 | -5,03 | -9,07 |
| Level of urbanization (ref: not urbanized) | | | | | | | | | | | | |
| Extremely | 0,52 | 0,68 | 0,68 | 0,66 | 0,84 | 0,34 | 0,47 | 0,63 | 0,62 | 1,03 | 1,02 | 0,01 |
| Strongly | 1,35 | 1,34 | 1,33 | 1,22 | 1,10 | 0,68 | 0,54 | 0,61 | 0,97 | 1,59 | 1,35 | 1,18 |
| Moderately | 1,13 | 1,16 | 1,25 | 1,14 | 1,07 | 0,77 | 0,29 | 0,71 | 0,65 | 0,92 | 0,77 | 0,93 |

| | | | | | | | | | | | | |
|---|---|---|---|---|---|---|---|---|---|---|---|---|
| Hardly | 1,26 | 1,05 | 0,87 | 0,70 | -0,03 | -0,35 | -0,83 | -0,64 | -0,17 | 0,46 | 0,61 | 0,30 |
| Family size (ref: 1-3 children) | | | | | | | | | | | | |
| ≥ 4 children | -8,35 | -8,49 | -8,18 | -9,84 | -8,58 | -9,66 | -9,98 | -9,58 | -8,83 | -9,72 | -9,37 | -11,26 |
| Institutional | -2,65 | -3,86 | -6,27 | -5,44 | -3,34 | -11,01 | -5,28 | -5,95 | -4,86 | -5,51 | -11,0 | -1,90 |
| Day-care attendance (ref: day-care yes) | | | | | | | | | | | | |
| Day-care no | -2,32 | -3,01 | -3,20 | -3,56 | -3,71 | -4,81 | -5,55 | -6,84 | -7,27 | -7,92 | -8,87 | -10,39 |

**Supplementary Table S6. Distribution of sociodemographic groups and number of unvaccinated children in birth cohort 2020**

| Category | | Population (N) | Total population (%) | Unvaccinated N | Unvaccinated population (%) |
|---|---|---|---|---|---|
| Total | | 169500 | 100% | 17990 | 100% |
| Education level mother | | | | | |
| | High | 73790 | 43,5% | 5000 | 27,8% |
| | Not high | 95710 | 56,5% | 12990 | 72,2% |
| Income household | | | | | |
| | First quartile | 18500 | 10,9% | 3420 | 19,0% |
| | Second quartile | 44860 | 26,5% | 6100 | 33,9% |
| | Third quartile | 60590 | 35,7% | 4790 | 26,6% |
| | Fourth quartile | 43400 | 25,6% | 2750 | 15,3% |
| Income source mother | | | | | |
| | Job in employment | 121260 | 71,5% | 8910 | 49,5% |
| | Self-employed | 13660 | 8,1% | 2090 | 11,6% |
| | Benefit recipient | 18510 | 10,9% | 3380 | 18,8% |
| | Pensioner | 270 | 0,2% | 50 | 0,3% |
| | Student | 2450 | 1,4% | 390 | 2,2% |
| | Other | 11750 | 6,9% | 2420 | 13,5% |
| Country of origin | | | | | |
| | The Netherlands | 97260 | 57,4% | 7230 | 40,2% |
| | Europe (excl. The Netherlands) | 20650 | 12,2% | 2540 | 14,1% |
| | Morocco | 8020 | 4,7% | 2770 | 15,4% |
| | Turkey | 6530 | 3,9% | 1350 | 7,5% |
| | Suriname | 5500 | 3,2% | 790 | 4,4% |
| | The Dutch Caribbean | 3720 | 2,2% | 610 | 3,4% |
| | Indonesia | 4270 | 2,5% | 340 | 1,9% |

| | | | | |
|---|---|---|---|---|
| Other, America/Oceania | 5090 | 3,0% | 500 | 2,8% |
| Other, Africa | 6790 | 4,0% | 880 | 4,9% |
| Other, Asia | 11670 | 6,9% | 980 | 5,4% |
| Level of urbanisation | | | | |
| Extremely urbanised | 38100 | 22,5% | 4990 | 27,7% |
| Strongly urbanised | 42400 | 25,0% | 4110 | 22,8% |
| Moderately urbanised | 31940 | 18,8% | 2810 | 15,6% |
| Hardly urbanised | 28430 | 16,8% | 2400 | 13,3% |
| Not urbanised | 25940 | 15,3% | 2650 | 14,7% |
| Family-size | | | | |
| 1-3 children | 155820 | 91,9% | 14230 | 79,1% |
| ≥ 4 children | 10740 | 6,3% | 2630 | 14,6% |
| Institutional household | 380 | 0,2% | 120 | 0,7% |
| Day care attendance | | | | |
| Yes | 123520 | 72,9% | 8000 | 44,5% |
| No | 45980 | 27,1% | 9990 | 55,5% |
